# Categorizing the Status of COVID-19 Outbreaks Around the World

**DOI:** 10.1101/2021.03.08.21252586

**Authors:** J. Paul Callan, Carlijn J.A. Nouwen, Axel S. Lexmond, Othmane Fourtassi

## Abstract

Although the SARS-CoV-19 virus spread rapidly around in world in early 2020, disease epidemics in different places evolved differently as the year progressed – and the state of the COVID-19 pandemic now varies significantly across different countries and territories. We have created a taxonomy of possible categories of disease dynamics, and used the evolution of reported COVID-19 cases relative to changes in disease control measures, together with total reported cases and deaths, to allocate most countries and territories among the possible categories. As of 31 January 2021, we find that the disease was (1) kept out or suppressed quickly through quarantines and testing & tracing in 39 countries with 29 million people, (2) suppressed on one or more occasions through control measures in 74 countries with 2.49 billion people, (3) spread slowly but not suppressed, with cases still increasing or just past a peak, in 31 countries with 1.45 billion people, (4) spread through the population, but slowed a result of control measures, leading to a “flattened curve” and fewer infections than if the epidemic were unmitigated, in 32 countries with 2.24 billion people, and (5) spread through the population with some but limited mitigation in 5 countries with 168 million people. In addition, several countries have experienced increases in cases after disease appeared to have finished spreading due to declining numbers of susceptible people. For some of these countries – for example Kenya, Pakistan and Afghanistan – the resurgences can be explained by the relaxation of control measures (and may have been enhanced by disease spread in population segments that experienced lower infection levels during the first waves). For other countries, the resurgences point to the effects of new virus variants with higher transmissibility or immunity resistance – including most countries in Southern Africa (where the B.1.351 variant has been identified) and several countries in West Africa (potentially due to the B.1.1.7 or other variants). These findings are consistent with mounting evidence of high infection rates in several low- and middle-income countries, both from seroprevalence studies and estimates of actual deaths from COVID-19 combined with estimates of expected mortality rates. We estimate that 1.3–3.0 billion people, or 17–39% of the global population, have been infected by SARS-CoV-2 to date, and that at least 4.5 million people have died from COVID-19 – much higher than reported cases and deaths. Disease control policies and vaccination strategies should be designed based on the state of the COVID-19 epidemic in the population – and consequently may need to be different in different countries.

**Key Points:**

✥ **The state of the COVID-19 pandemic varies significantly in different countries and territories around the world – and policies for disease control and vaccination will need to be tailored accordingly**.
✥ **In any epidemic, there are several possibilities for how the disease will spread over time – and our analysis finds that, in fact, as of 31 January 2021, there were many countries and territories in each of the main categories of COVID-19 epidemic dynamics that might have been expected:**
  - Kept out or suppressed quickly through quarantines and testing & tracing – in 39 countries with 29 million people (0.4% of the global population), mostly small island states and a few countries in Southeast Asia. *[Category H in the following map and table]*
  - Suppressed through control measures (social distancing, hygiene and testing & tracing) –in 74 countries with 2.49 billion people (31.9% of global population), mostly in Europe, East Asia and the Pacific. *[Categories F and G]*
  - Spread slowly but not suppressed, with cases still increasing or just past a peak – in 31 countries with 1.45 billion people (18.6% of global population), including many countries in Latin America, Eastern Europe and the Middle East, as well as the United States and Russia. *[Categories D and E]*
  - Spread through the population, but slowed as a result of control measures, leading to a “flattened curve” and fewer infections than if the epidemic were unmitigated – in 32 countries with 2.24 billion people (28.8% of global population), mostly in South and Southeast Asia (including India) and Africa. *[Category B]*
  - Spread through the population with some but limited mitigation or “flattening the curve” –in 5 countries with 168 million people (2.2% of global population). *[Category A]*
  - Experienced increases in cases after disease appeared to have finished spreading, which in some countries might have been solely due to relaxation of control measures (especially in wealthier population segments which experienced low infection levels during the first wave) – for example in Kenya and in Pakistan and some Central Asian countries – but which in some countries is likely to be due to new virus variants with higher transmissibility or immunity resistance – for example in most countries in Southern Africa and several in West Africa, and possibly also in parts of South and Central America. *[Category J and many countries in Category K]*

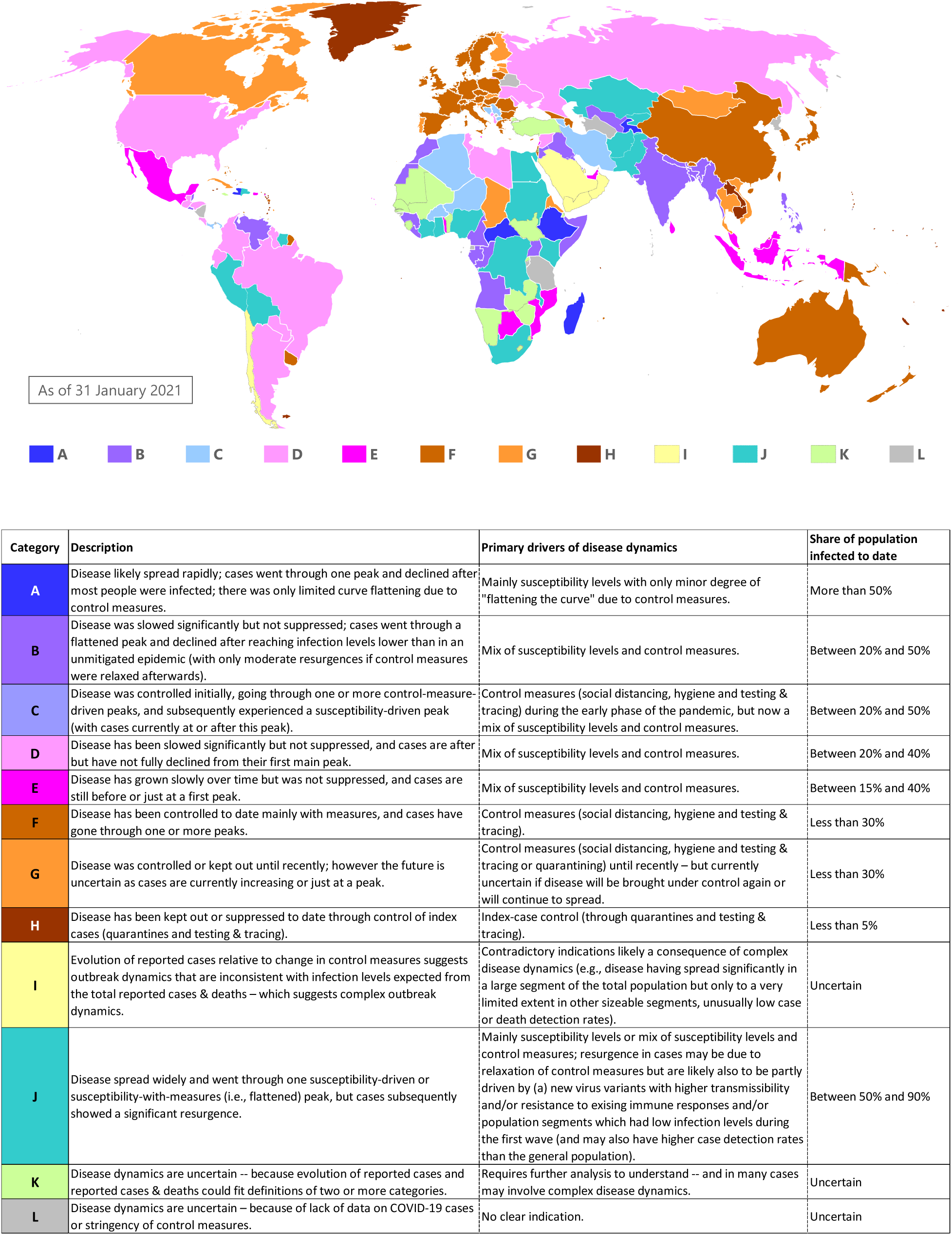

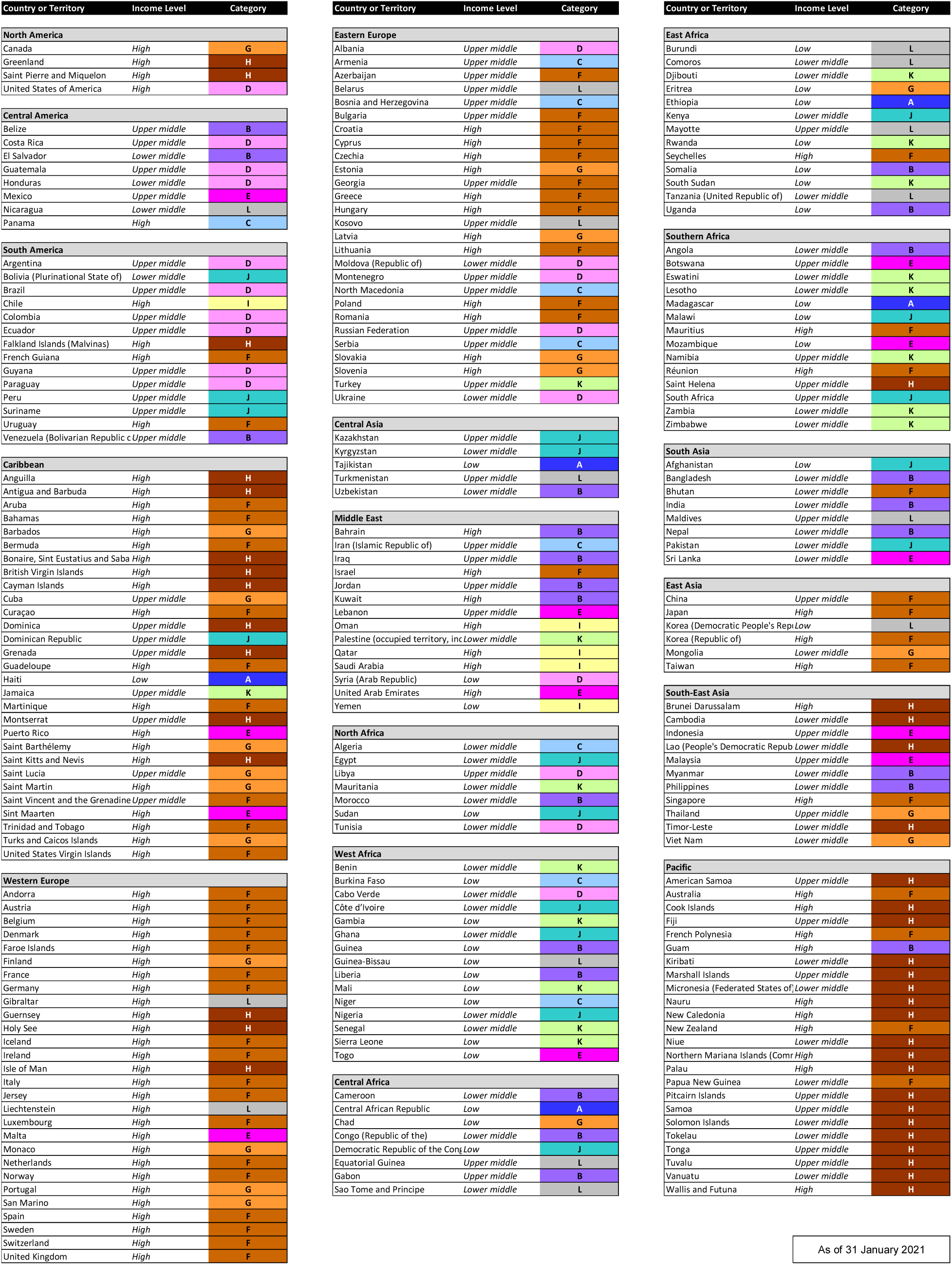
✥ **These findings are backed up by mounting evidence of high infection rates in several low- and middle-income countries**. Seroprevalence studies in Kenya, Nigeria, Pakistan and South Africa have reported finding antibodies for SARS-CoV-2 in large percentages of the studied populations – and suggest that current infection levels are likely above 50% in each country. Studies of actual deaths due to COVID-19, combined with estimates of expected mortality rates, similarly suggest that SARS-CoV-2 has, by now, infected more than half of the populations in Bolivia, Ecuador, Peru, Mexico, South Africa, Sudan, Syria, Yemen and Zambia.
✥ **Countries of all income levels, and from all regions, appear in each of the main disease dynamics categories; however, there are clear income and geographical patterns in states of COVID-19 epidemics around the world**. Most high-income countries have controlled the spread of SARS-CoV-2 through measures. Middle-income countries are spread across all categories, and account for 45 of the 63 countries which have slowed the disease significantly but not fully suppressed it. Some low-income have experienced largely unmitigated susceptibility-driven dynamics, while others have “flattened the curve” to varying degrees.
✥ **We estimate that between one and two out of every five people globally has been infected by SARS-CoV-2 to date, and that at least 4.5 million people have died from COVID-19**. Our estimate of total infections – 1.3–3.0 billion people, or 17–39% of the global population – is between 13 and 30 times the number of confirmed cases, and twice to four times as much as previous estimates of total infection numbers. We estimate that 4.6–10.0 million people have died from COVID-19, between 2.1 and 4.5 times the number of deaths attributed to COVID-19.
✥ **An estimated 8.9-12.5 million lives remained at risk from COVID-19 as of the end of January 2021, prior to vaccination efforts –** mainly in high-income countries (2.4–2.9 million), China (2.1 million) and India (1.7–2.9 million). Vaccinations, of course, have already started to reduce these numbers substantially.
✥ **Our analytical approach is simple but useful – providing insight into the epidemic status even in many low-income countries with limited disease monitoring, and with potential to provide early warnings of significant new variants**. We compare the evolution of reported cases with changes in stringency of disease control measures, and check that infection levels are plausible given total reported cases and deaths and the income level of the country. Anomalies in which changes in the evolution of reported cases cannot be explained by changes in the stringency index provide indications of possible significant variants of the virus. Up to the end of January, the data provide indications of the presence of significant new variants in:
  - Most countries in Southern Africa (where the B.1.351 variant, with higher transmissibility and some resistance to immunity, was first identified in South Africa)
  - Several countries in West Africa (likely with higher transmissibility and resistance to immunity, possibly the B.1.1.7 variant, which was first identified in the UK and has been found in Ghana and Nigeria, or possibly a different variant).
  They also suggest, with less certainty, that the disease dynamics may be affected by new variants in several countries in South and Central America (perhaps the P.1 variant descended from the B.1.1.28 variant which was first identified as coming from the Brazilian Amazon).
✥ **Different countries should adopt different disease control policies, according to the state of the COVID-19 epidemic in the population**.
  - For countries that have kept the disease out or suppressed outbreaks through control measures, their measures need to be kept in place – and potentially strengthened especially in the face of higher-transmissibility variants – until vaccines have been widely administered.
  - For countries in which the disease is spreading slowly, full control measures should be maintained at least until new case numbers fully decline from the peak; later, it may be possible to relax some measures, but if measures are relaxed too soon or too much after cases peak, then significant further outbreaks can be expected (as has already happened in several such countries).
  - For countries in which cases have declined following a flattened curve, there may be room to relax control measures that have the greatest negative health, economic and social consequences – but the most effective control measures will need to be maintained (even when cases remain low for extended periods), and measures may need to be strengthened to tackle variants which higher transmissibility or ability to evade immune responses.
  - For countries in which the disease spread was largely unmitigated, many control measures could be relaxed for most people – although there may be risks if sizeable population segments have much lower infection levels than the general population or from variants with a high degree of immunity resistance.
✥ **These findings may have implications for the optimal distribution of early batches of vaccines within countries**.
  - For countries that have kept the disease out or suppressed outbreaks through control measures, vaccinations should be given first to frontline healthcare and essential workers and to elderly and vulnerable groups (starting with the oldest and most vulnerable).
  - For countries in which the disease is spreading slowly, detailed modelling should be done to determine whether the optimal strategy is to vaccinate at-risk groups first or to vaccinate key transmitters to halt the outbreak and “crush the curve” while waiting for further vaccine supplies to arrive. For any country choosing the key transmitter strategy – as Indonesia has done and has been suggested for the United States – it will be essential to maintain control measures, and to keep higher transmissibility variants out, or otherwise the benefits of a key transmitter vaccination strategy could be lost.
  - For countries in which the cases declined following a flattened curve, vaccinations should probably be given first to elderly and vulnerable groups, but the optimal strategy may switch to vaccinating key transmitters if there are resurgences in cases due to higher-transmissibility or immunity-resistant variants.
  - For countries in which the disease spread was largely unmitigated, vaccination should concentrate on elderly and vulnerable people, because population-level immunity already exists, and the greatest danger lies in vulnerable people becoming infected due to endemic SARS-CoV-2 from variants that will likely circulate over many years.
✥ **These findings may also have implications for the optimal distribution of the first vaccines across countries**. For most countries, the optimal allocation of vaccines doses is likely still to be according to population size – as current recommendations suggest. However, the global optimal allocation strategy might include providing somewhat greater supplies, during the next few months, to countries where using the vaccine to halt spread of the disease might be possible (provided that disease control measures are maintained in those countries).
✥ **Vaccination strategies will need to account for current and potential future virus variants as well as the likelihood that immunity from vaccination will wane over time**. Higher-transmissibility variants of the SARS-CoV-2 virus increase the urgency of distributing vaccines in countries which have controlled the disease to date, and may alter the optimal strategy for countries deciding between vaccination first of elderly and vulnerable people or of key transmitters. Immunity-resistant variants of the virus may reduce the effectiveness of current vaccines, but are not likely to negate fully the protection they offer. Immunity acquired through vaccination is likely to wane over time – like immunity acquired through infection. In many, perhaps most, countries, the time to vaccinate the whole population will exceed the timeframe in which immunity from vaccination wanes or new immunity-resistant variants emerge. Looking to the longer term, therefore, new virus variants and waning immunity are likely to necessitate re-vaccination (with vaccines tailored to the latest variants) on a regular basis – and the optimal long-term strategies for ongoing vaccination will vary widely across countries and will depend on many factors.

**Summary:** **The state of the COVID-19 pandemic varies significantly in different countries and territories around the world – and policies for disease control and vaccination will need to be tailored accordingly**. Although the SARS-CoV-19 virus spread rapidly around in world in early 2020, the state of disease epidemics in different countries diverged rapidly as the year progressed. Many high-income countries have had second or third waves; other countries have seen cases continue to increase gradually; still others have experienced declines in cases to low levels after peaks in mid-2020. Facing different situations, different countries might need to adopt different policies in the coming months, including different disease control measures and vaccination strategies.

**Each country needs to know its COVID-19 status**. There are several possible courses that a disease epidemic can take in a population. The disease can spread rapidly until its runs out of people remaining to infect; the disease can be slowed with control measures but still spread until large numbers of people are infected and immune; the disease can be suppressed or “crushed” by control measures; or the disease can be kept out completely. More complex disease dynamics will occur when a virus mutates, if new variants evade immune responses in people already infected or spread faster than before, or if the disease spreads differently in different segments of a population. Our research suggests that different countries have experienced outbreaks in each of the main possible categories:

- **Susceptibility-Driven Dynamics with Limited Mitigation *[Category A]*** – in which the disease spread until it infect most people and declined due to low susceptibility levels (i.e., low share of the population still able to be infected).
- **Susceptibility-Driven Dynamics Mitigated by Measures *[Categories B and C]*** – in which the disease curve was “flattened” by control measures, but the disease still spread and declined after infecting large numbers of people (but fewer than if there were no mitigation).
- **Susceptibility-plus-Measures-Driven Dynamics *[Categories D and E]*** – in which the disease was slowed significantly but not suppressed, i.e., the curve was “flattened” but not “crushed”, and the disease is still spreading in the population.
- **Measures-Driven Dynamics *[Categories F and G]*** – in which the disease has been constrained to date mainly through control measures (social distancing, hygiene and testing & tracing), but, of course, could spread again if measures are relaxed because only a minority of the population has been infected.
- **Index-Case-Control Dynamics *[Category H]*** – in which the disease has been kept out or suppressed to date through strict control measures (especially quarantines and testing & tracing).
- **Complex Disease Dynamics due to Differences Across Population Segments and/or New Variants *[Categories I and J, and many countries in Category K]*** – in which the disease experienced an apparently susceptibility-driven curve but with low overall infection levels (i.e., share of population infected) because some segments of the population have not had many infections, or in which the disease later shows an unexpected resurgence, due to spreading within the previously less-affected population segments or due to the emergence of immunity-evading strains of the virus.

**From reported data on COVID-19 cases and disease control measures, we can categorize, for most countries and territories, the dynamics of the disease to date**. First, we compare the timing of increases and/or decreases in reported new cases with the timing of changes in the “Stringency Index” of control measures compiled by the Oxford COVID-19 Government Response Tracker – and select the appropriate disease dynamics category. Second, we check if the infection levels expected for the category or categories indicated in the first step, are consistent with predicted ranges from the total numbers of reported cases and deaths using plausible ranges for the case detection rate, death detection rate and infection fatality ratio (IFR) given the country’s income level. This approach yields definitive categories for most countries and territories. COVID-19 disease dynamics are complicated in many countries, due to changes in control measures, seasonal patterns, geographical differences within countries, variability in case testing over time and emergence of new variants; such effects can be seen in the reported cases and deaths for many countries, but they do not obscure the basic drivers of disease dynamics – in other words, which of the categories applies – for most countries.

**The results suggest that there is a wide variation in the state of the COVID-19 epidemic around the world – as of 31 January 2021 – as illustrated in the map and the table below**. COVID-19 has been suppressed through control measures – Categories F, G and H – in 113 countries and territories with 2.52 billion people or about 32.3% of the global population. However, the rest of the world are in different situations. A total of 31 countries, with populations of 1.45 billion people (18.6% of global population), fall into Categories D and E, meaning that the disease spread has been slowed but not suppressed and cases are currently still increasing or just past their peak. In the 23 countries of Category B, home to 2.05 billion people (26.3%), the disease was slowed but not suppressed, and cases have declined fully from the peak. In a further 9 countries with 0.19 billion people (2.5%), the disease spread through the population after initial waves were suppressed. COVID-19 outbreaks in 5 countries with 0.17 billion people (2.2%) were only somewhat mitigated by control measures and the virus has likely infected most of the population, falling into Category A.

**For several countries, which have apparent anomalies and fall outside the “basic” categories, the methodology provides important insights into epidemic status – pointing to situations where significant differences may exist across population segments or providing early warnings of new variants with higher transmissibility or resistance to immunity**. For 5 Arabian Peninsula countries (3 of which are in Category I) and Singapore, it is likely that the virus has spread widely among migrant worker communities but has been controlled in the rest of the population. Categories J and K include 28 countries in which reported cases have surged after first waves which were likely or possibly susceptibility-driven, with curves flattened to various extents as a result of control measures which mitigated the epidemics. For some countries – including (1) Kenya, (2) Pakistan, Afghanistan, Kyrgyzstan and Kazakhstan, and (3) Egypt and Sudan – the second peaks are likely due to relaxation of measures but larger than might be expected due to disproportionate effects of the second waves on population segments (likely more affluent groups) which had lower infection levels during the first waves. For other countries – including (1) most countries in Southern Africa and (2) many countries in West Africa – the data suggest the presence of new virus variants with higher transmissibility and possible resistance to immunity, because resurgences or accelerations in cases happened in several neighbouring countries around the same time, and often without changes in control measures, and the second surges in cases usually involved faster increases than the first waves. The B.1.351 variant, with higher transmissibility and some resistance to immunity, was first identified in South Africa and is known to have caused most cases in the country’s second wave; the B.1.1.7 variant, which has higher transmissibility, has been found in Ghana and Nigeria. Several countries in South and Central America have experienced second waves or surges in cases: Suriname’s might be due to a higher-transmissibility variant (perhaps the P.1 variant that was first identified as coming from the Brazilian Amazon); Bolivia’s was large but could be explained by a significant decline in control measures; increases in Brazil and several other countries across South and Central America might simply be due to relaxation of social distancing behaviours over the Christmas and New Year holiday season although a role for virus variants cannot be discounted.

**Countries of all income levels appear in each of the main disease dynamics categories; however there are clear correlations between income groups and COVID-19 status categories**. Most high-income countries have controlled the spread of SARS-CoV-2 through measures (and thus fall in Categories F, G and H). Middle-income countries are spread across all categories, and account for 45 of the 63 countries which have slowed the disease significantly but not fully suppressed it (Categories B, C, D and E). Some low-income countries have experienced largely unmitigated susceptibility-driven dynamics (Category A), while others have “flattened the curve” to varying degrees (Categories B, C, D and E). A mix of low- and middle-income countries are among the 34 countries in Categories J and K.

**Clear geographical patterns have emerged in the states of COVID-19 epidemics**. There was more diversity in the state of the epidemic within regions earlier in the pandemic, but regional patterns had become clear by the end of January 2021.

- In the Americas, the disease has spread slowly but has not been suppressed (Categories D and E) in most countries, including those with the largest populations, while many (but not all) of the Caribbean islands have kept SARS-CoV-2 out or under control (Category H).
- Western and Northern European countries have, for the most part, controlled the disease through social distancing and hygiene measures, through two or three waves, and fall in Categories F and G.
- Across Eastern Europe, the Levant, the Caucuses and Iran, all countries have constrained growth of the disease significantly, but infection levels in most have grown to moderate levels: different countries in these regions are included in Categories C, D/E and F/G, although their infection levels may all be in the moderate range.
- In South and Central Asia, the virus has spread widely in most countries and cases have declined. In India, Bangladesh, Nepal and Uzbekistan, the case curve was flattened considerably, and current infection levels are likely moderate (Category B). In Pakistan, Afghanistan, Kyrgyzstan, and Kazakhstan (all in Category J), there have been two peaks in cases. Bhutan has contained the outbreaks of the virus to date (Category F).
- Many countries in East and South-East Asia have largely kept the disease under control or kept it out (Categories F, G and H). However, Malaysia, Mongolia and Myanmar experienced widespread outbreaks in the second half of 2020, the Philippines appears to be past the peak of its epidemic (Category B), and Indonesia has had a continuous but very slow rise in cases since the start of the pandemic (Category E).
- In Australia, New Zealand and most Pacific Island States, SARS-CoV-2 has been excluded through quarantines, together with testing and tracing and lockdowns when the virus has spread beyond quarantined individuals (Categories F and H).
- African countries appear to have differed greatly in how the disease has spread. Many countries appear to have experienced widespread epidemics followed by declines in case numbers, with varying degrees of “curve flattening” due to control measures (Categories A and B). In some countries – Tunisia, Libya, Togo, Botswana and Mozambique – cases spread very slowly (Categories D and E). A few countries appear to have kept the disease out, and a few others appear to have experienced full outbreaks after having previously kept the virus largely out. As described earlier, most countries in Southern Africa and many in West Africa experienced rapid growth in case numbers in December and January (putting many in Categories J and K) – suggestive of the presence of one or more new variants with higher transmissibility and possible resistance to immunity.

**We estimate that 1**.**3–3**.**0 billion people have been infected by SARS-CoV-2 to date, or about 17–39% of the global population**. This estimate is between 13 and 30 times the number of confirmed cases, and perhaps twice to four times as much as previous estimates of total infection numbers. We estimate that 4.6–10.0 million people have died from COVID-19, between 2.1 and 4.5 times the number of deaths attributed to COVID-19.

**An estimated 8**.**9-12**.**5 million lives remain at risk from COVID-19, which can be saved through appropriate disease control measures and effective deployment of vaccines**. Of these estimated extra deaths, if 90% of the population were to contract SARS-CoV-2, high-income countries account for about 2.4–2.9 million, China for about 2.1 million, and India for about 1.7– 2.9 million. Vaccinations, of course, have already started to reduce these numbers substantially.

**The findings of this report are backed up by mounting evidence of high infection rates in several low- and middle-income countries**. Immunity testing provides direct evidence of the current state of the COVID-19 epidemic. Serological studies in several cities and regions in Brazil, India, Kenya, Pakistan, Qatar and South Africa have already reported finding antibodies for SARS-CoV-2 in large percentages of the studied populations. Note, however, that serological testing will underestimate the number of people who have been infected, due to waning of SARS-CoV-2 antibodies which affects significant numbers of people at about 4-6 months after infection. Consequently, serological testing might understate the actual degree of immunity in a population, because some people may have antibodies at levels below the detection threshold of the serology tests or may have memory B cell or T cell responses, either or both of which will likely reduce the severity of their illness if reinfected, and may reduce their vulnerability to reinfection and their likelihood to pass on the virus to other people if reinfected. In some places, reliable estimates of actual deaths due to COVID-19 may be a substitute for immunity testing to determine the share of population infected to date, at least approximately. Estimates, using a variety of methodologies, in Bolivia, Ecuador, Mexico, Peru, South Africa, Sudan, Syria, Yemen and Zambia all indicate that moderate to high shares of their populations have already been infected.

**Different countries should adopt different disease control policies, according to the state of the COVID-19 epidemic in the population**. The following recommendations for countries in different categories take into account their current infection levels and the potential for additional infections if measures are relaxed or if new variants become common in a country.

➢ ***Category A:*** Control measures should be relaxed for most people; such relaxation is not likely to lead to many more cases and deaths. In some low- and middle-income countries, wealthier population segments may have implemented greater degrees of social distancing during the epidemic to date, and have much lower infection levels than in the overall population; these segments should maintain social distancing, until vaccines arrive, because otherwise they could experience substantial outbreaks (which may have generated “second waves” in some countries). If and when new virus strains with higher transmissibility and/or resistance to immunity arrive, control measures should be strengthened again to avoid new outbreaks, if the new variants cause high mortality levels and if it seems likely that control measures will be more effective at controlling the new outbreaks than they were during the initial outbreaks.
➢ ***Categories B and C:*** Control measures currently in place that have the greatest negative health, economic and social consequences could be relaxed. However, many control measures, especially the most effective in limiting virus spread, will need to be maintained, even though current case numbers are low; otherwise, significant resurgences can take place (as has happened, for instance, in Kenya and Bolivia). Population segments that may have maintained lower infection levels during the outbreak to date will need to maintain social distancing. If and when new virus strains with higher transmissibility and/or resistance to immunity arrive, control measures will likely have to be strengthened again to avoid new outbreaks.
➢ ***Categories D and E:*** Control measures should be maintained at least until new case numbers fully decline from the peak; if measures are relaxed too soon after cases peak, then significant further outbreaks can be expected (as has happened, for instance, in Brazil, Colombia and Paraguay). Once cases fully decline from the peak – through further infections or as a result of vaccination programmes – then, and only then, some of the disease control measures with the greatest negative health, economic and social consequences could be relaxed. For some countries in Categories D and E, it may be possible to push *R*_*0_e*_ below 1 and hence “crush the curve” by introducing some additional control measures or improving compliance with existing measures. New virus strains, especially with higher transmissibility, can generate resurgences or accelerations in growth of cases (as seen, for example, in Mozambique and Togo).
➢ ***Categories F and G:*** COVID-19 control measures, put in place by governments and implemented by citizens, have saved perhaps 13.1–14.2 million lives. To continue to protect these lives, control measures need to be maintained until vaccines become widely available – and strengthened, if necessary, to compensate for new virus variants with higher transmissibility.
➢ ***Category H:*** Measures to keep the disease out – mainly strict quarantines for new arrivals and testing & tracing of suspected cases – should be maintained until vaccines become widely available.

**The findings of this report may have implications for the optimal distribution of early batches of vaccines within countries**. Current policies in several countries call for deployment of vaccines first to healthcare workers and then by age cohort, starting with the oldest. These plans are aligned with the results of modelling (by Imperial College London and others) which suggest that, when the supply of vaccines is limited, the optimal strategy is to target the elderly and other high-risk groups. However, the models indicate that, if the supply is sufficient to stop transmission of the virus, the optimal strategy switches to targeting key transmitters (e.g., working age people and potentially children) to indirectly protect the elderly and vulnerable. Consequently, the optimal strategy may vary according to the disease status category for each country:

➢ ***Category A:*** Vaccination should concentrate on elderly and vulnerable people, starting with the oldest and most vulnerable. There is no alternative strategy to consider because population-level immunity already exists, and the greatest danger lies in vulnerable people becoming infected due to endemic SARS-CoV-2.
➢ ***Categories B and C:*** Vaccinations should probably be given first to elderly and vulnerable groups, and to frontline healthcare and other essential workers. However, if there are resurgences in cases across the population due to higher-transmissibility or immunity-resistant variants, then the optimal strategy may switch to targeting key transmitters, similar to some countries in Categories D and E.
➢ ***Categories D and E:*** In some of these countries, the optimal strategy may to be vaccinate key transmitters – *while maintaining current disease control measures* – because it may be possible to halt the outbreak and “crush the curve”, while waiting for further vaccine supplies to arrive (after which disease control measures could be released). This strategy is being pursued by Indonesia and was suggested for the United States of America in a recent paper. However, careful modelling and planning would be necessary, for any country considering such an approach, to determine if a key transmitter strategy would in fact be optimal and if it would be feasible to implement. Further, for such a strategy to work, it will be necessary to keep control measures in place and to keep high-transmissibility variants of the virus out, until enough people have been vaccinated.
➢ ***Categories F, G and H:*** Vaccinations should be given first to frontline healthcare and other essential workers and to elderly and vulnerable groups (starting with the oldest and most vulnerable).

**These findings may also have implications for the optimal distribution of the first vaccines across countries**. Modelling by the Imperial College London COVID-19 Response Team suggests that the optimal allocation of vaccine doses among countries “is sensitive to many assumptions and will vary depending both on the vaccine characteristics and the stage of the epidemic in each country at vaccine introduction,” and concluded that, “[g]iven this uncertainty, allocating vaccine doses according to population size appears to be the next most efficient approach.” Our findings reinforce the uncertainty strongly: it is very likely that that stage of the epidemic varies greatly across countries. For most countries, the optimal allocation of vaccines doses is likely still to be according to population size – and then for those countries to give doses first to elderly and vulnerable people. However, the global optimal allocation strategy might include providing somewhat greater supplies, during the next few months, to Category D and E countries where using the vaccine to halt spread of the disease might be possible (provided that disease control measures are maintained in those countries). It is clear, in any case, that further modelling of vaccine allocation strategies is essential, taking into account the actual vaccine efficacies and projected available doses by month, as well as allowing for disease stage categories in different countries.

**Vaccination strategies will need to account for current and potential future virus variants as well as the likelihood that immunity from vaccination will wane over time**. Higher-transmissibility variants of the SARS-CoV-2 virus increase the urgency of distributing vaccines, especially in Category F and G countries which may struggle to keep the disease suppressed, and might cause vaccination of key transmitters to be a less effective strategy for Category D and E countries if higher-transmissibility variants mean that they can’t suppress the disease fully with limited vaccinations. Immunity-resistant variants of the virus may reduce the effectiveness of current vaccines, but are not likely to negate fully the protection offered by existing vaccines. Immunity acquired through vaccination is likely to wane over time – like immunity acquired through infection. Looking to the longer term, new virus variants and waning immunity are likely to necessitate re-vaccination (with vaccines effective against the latest variants) on a regular basis. In many, perhaps most, countries, the time to vaccinate the whole population will exceed the timeframe in which immunity from vaccination wanes or new immunity-resistant variants emerge. In making long-term plans, therefore, countries may face a wide range of options for who to vaccinate (elderly and vulnerable populations, key transmitters or entire populations) and for frequency of vaccination (every 6 months, annual, or once if residual benefits are sufficient). Optimal strategies for each country will be complicated to determine, as the choice will depend on many factors, including vaccine effectiveness in reducing mortality and in reducing transmission, how effectiveness wanes over time, mortality rates and transmissibility of new variants (in general and in previously infected or vaccinated people), and, once the risks to life and health from “endemic COVID” decrease to the point where COVID-19 is not an overriding issue, comparison with other health and budgetary priorities.

## 1 Introduction

COVID-19 upended the world in 2020 and, despite the arrival of vaccines, is likely to continue to impact lives for most of 2021 and beyond. As COVID-19 cases rose rapidly around the world in March 2020, governments and individuals imposed significant constraints on regular economic and social life in order to control the spread of the disease and save lives *[1]*. In recent months, COVID-19 cases have moved in different directions in different parts of the world – for example, experiencing second and sometimes third peaks in cases in many high-income countries (HICs) in Europe, declining to low levels in some low- and middle-income countries (L&MICs) but rising sharply in December and January in other L&MICs, and increasing gradually over time in a few countries *[2]*. Facing different situations, different countries might need to adopt different strategies during the year ahead, both for controlling the virus and for vaccination. Figuring out how to manage COVID-19 outbreaks first requires knowing the current status of the disease. The most basic questions are: how many people have really been infected to date, how many people have (at least temporary or partial) immunity, and how many people remain who could be infected in the future?

Several research studies have suggested that some L&MICs might have moderate to high levels of infection. Seroprevalence studies in Qatar *[3]*, Manaus (Brazil) *[4]*, Pakistan *[5]*, South Africa *[6]* and Nairobi (Kenya) *[7]* reported finding antibodies for SARS-CoV-2 in large percentages of the studied populations. Estimates of actual deaths due to COVID-19, combined with estimates of expected mortality rates, similarly suggest that SARS-CoV-2 has infected large shares of the populations in several L&MICs *[8]*, including Bolivia, Ecuador, Peru, Mexico and South Africa *[9]*, Sudan *[10]*, Syria *[11]*, Yemen *[12]* and Zambia *[13]*. In previous work, we showed that COVID-19 cases in some L&MICs could be described with the simplest of all epidemiological models, the Reed-Frost model, with disease outbreak parameters suggesting that the outbreaks had passed the point where cases were declining due to low susceptibility levels in the population, noting that control measures reduce the infection level at which cases start to decline *[14]*. (In this report, we use the terms infection level, immunity level and susceptibility level to refer to the percentages of the population that have been infected, are immune and are susceptible, respectively.)

It is evident from the recent increases in cases, hospitalizations and deaths that the cumulative infections and immunity levels are not above “herd immunity” levels in most HICs. Researchers at Imperial College London and others argued convincingly in June 2020 that European countries had not reached herd immunity at that time *[15]* – and the subsequent increases in cases in Europe proved that they were correct.

In rejecting the idea of “herd immunity” in Europe, Okell et al. relied on differences in per-capita mortality rates between countries that went into lockdown early or not and on a clear relationship between the prevalence of antibodies to SARS-CoV-2 and mortality from COVID-19 across countries *[15]*. In our previous work, suggesting that declines in cases and deaths in selected L&MICs in the third quarter of 2020 might be due to increasing shares of the populations that were infected and became immune, we relied on observing declines while control measures were constant or relaxing, on the shape of the reported cases curves (which are nearly symmetrical for the L&MICs studied and not for European comparator countries), and on the plausibility of very low case and death detection rates in L&MICs based on serological studies and excess deaths estimates in some L&MICs *[14]*. Both arguments illustrate how it is possible to infer whether an outbreak’s dynamics has been primarily susceptibility-driven, or primarily control-measure-driven, or driven by a combination of control measures and declining susceptibility levels, even in the absence of knowing the total infection level directly.

## 2 Taxonomy of Possible Trajectories for an Infectious Disease Epidemic

Disease outbreaks can evolve in a variety of different ways depending on the effects of disease control measures taken by governments and individuals.

Figure 1 illustrates how a disease like COVID-19 would be expected to evolve in different circumstances. Each panel shows expected case numbers following the introduction of the virus to a population under different scenarios for levels of control measures introduced after the onset of the epidemic.

**Figure 1:**
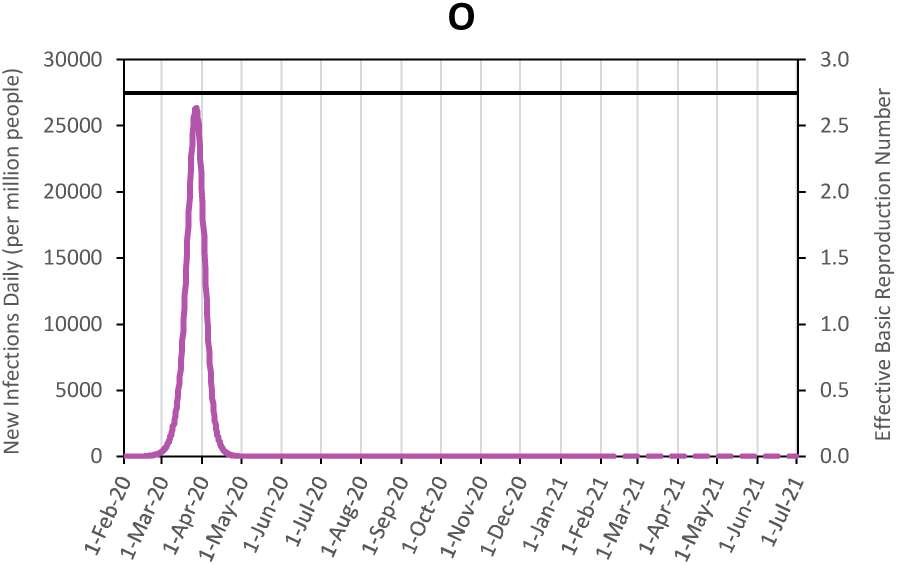
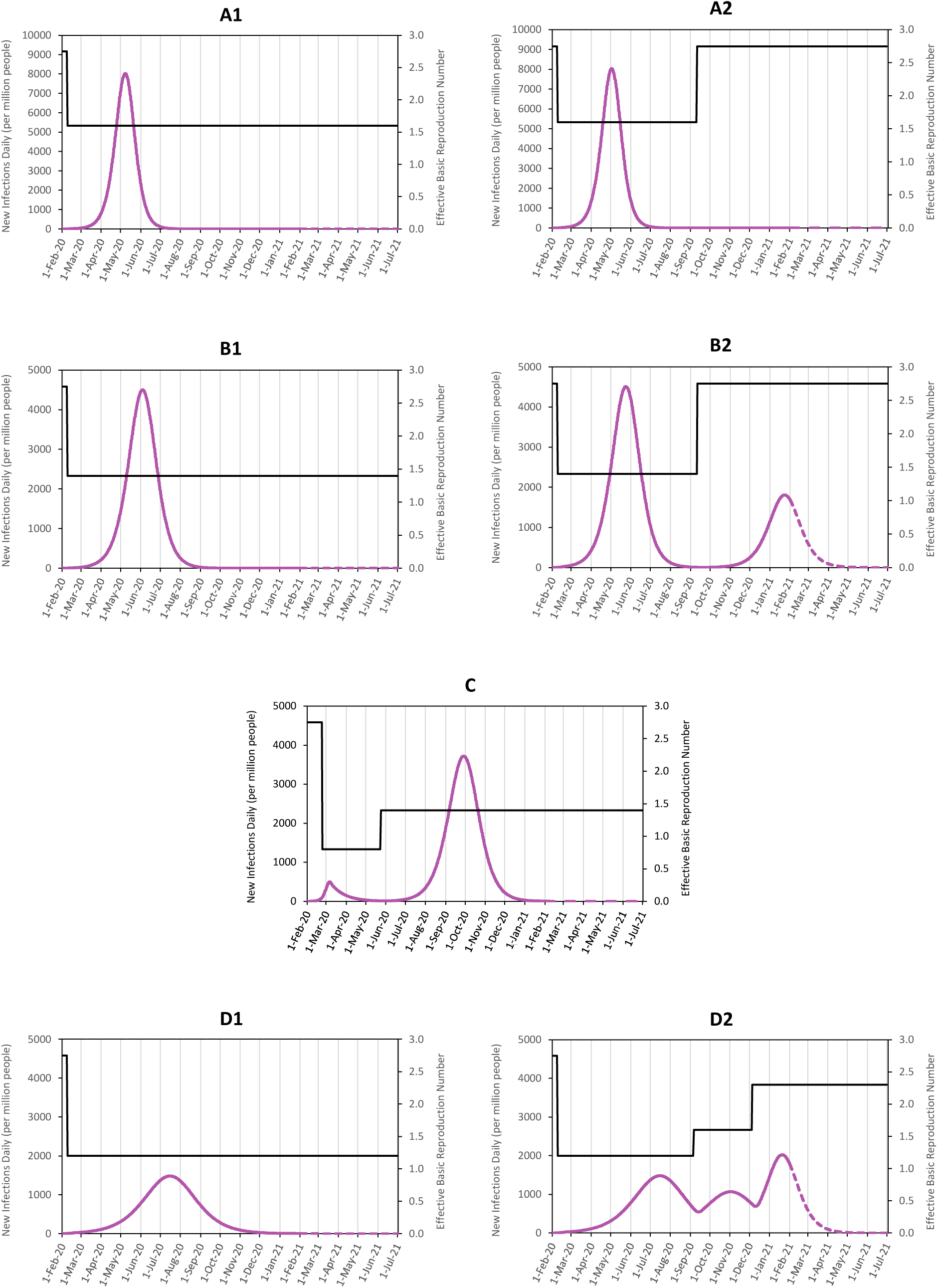
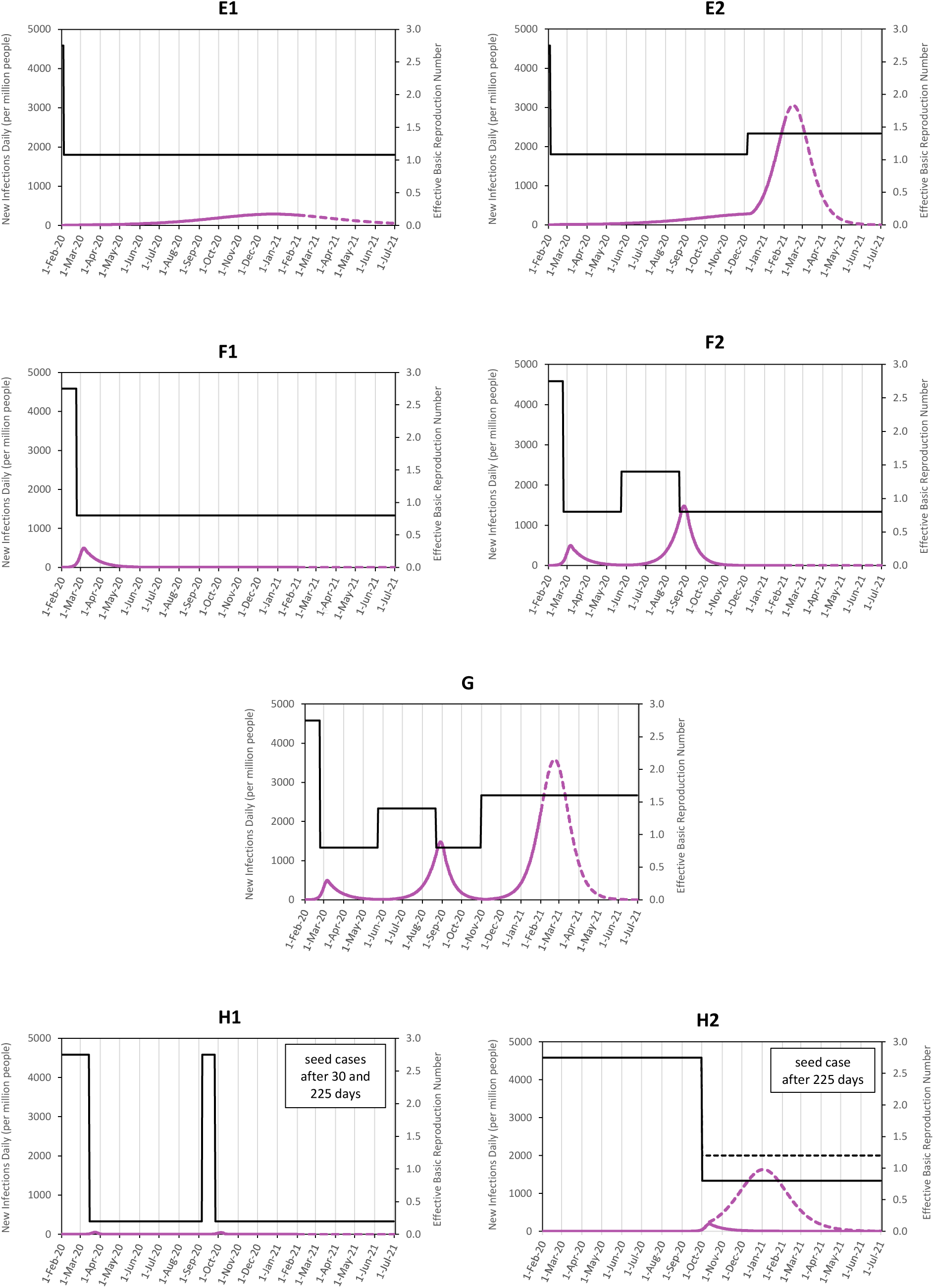
Evolution of numbers of disease cases, following a simple model in a homogeneous population, under different disease control circumstances. Calculated values for new daily cases per million people (purple lines), from a simple SIR (susceptible-infected-recovered) model, with basic reproduction number *R*_*0*_ = 2.75 and mean generation time *t*_*g*_ = 5 days, and assuming initial outbreak on 1 February (except for panels H1 and H2). Different panels use different assumptions regarding the introduction or relaxation of disease control measures, resulting in changes in effective basic reproduction number *R*_*0_e*_ (shown in black lines; stronger measures lower the value of *R*_*0_e*_), as follows: (**O**) No control measures and *R*_*0_e*_ = *R*_*0*_ = 2.75. (**A**) (1) Control measures introduced which reduce *R*_*0_e*_ to 1.6 at 15 days after initial outbreak, and (2) control measures relaxed to increase *R*_*0_e*_ to 2.75 after 230 days. (**B**) (1) *R*_*0_e*_ reduced to 1.4 after 15 days, and (2) increased to 2.75 after 230 days. (**C**) *R*_*0_e*_ reduced to 0.8 after 30 days and increased to 1.4 after 120 days. (**D**) (1) *R*_*0_e*_ reduced to 1.2 after 15 days, and (2) increased to 1.6 after 225 days, and increased to 2.3 after 315 days. (**E**) (1) *R*_*0_e*_ reduced to 1.08 after 10 days, and (2) increased to 1.4 after 315 days. (**F**) (1) *R*_*0_e*_ reduced to 0.8 after 30 days, and (2) increased to 1.4 after 120 days and decreased to 0.8 after 210 days. (**G**) *R*_*0_e*_ reduced to 0.8 after 30 days, increased to 1.4 after 120 days, decreased to 0.8 after 210 days, and increased to 1.6 after 280 days. (**H**) (1) Seed cases introduced after 30 days and 225 days, and *R*_*0_e*_ reduced to 0.2 at 20 days after each seed case to simulate the effect of intensive testing & tracing. (2) Seed case introduced after 225 days, and *R*_*0_e*_ reduced to 0.8 after 250 days (solid line) or reduced to 1.2 after 250 days (dashed line). Changes in *R*_*0_e*_ are implemented gradually through linear increases or decreases over 10 days from 5 days before to 5 days after the time specified. Note that the maximum value of the scale for new daily infections is greater for panels O and A than the other panels.

For an uncontrolled outbreak, in which people gain at least temporary immunity after infection, disease cases grow exponentially at first, reach a peak when the remaining susceptible population is not large enough to sustain further growth, and then decline exponentially (figure 1, panel O). The outbreak dynamics are determined in part by the *basic reproduction number, R*_*0*_, the average number of new infections caused by each current infected individual, at the outset of the disease outbreak when few people have been infected yet. The peak of the case curve is reached as the proportion of the population that has been infected and become immune increases beyond 1– 1/*R*_*0*_ – commonly referred to as the *herd immunity threshold*. For the initial variants of COVID-19, it has been estimated that *R*_*0*_ is between 2.3 and 3.5 *[16]*.

Disease control measures including social distancing, handwashing and wearing masks reduce *R*_*0*_ to an *effective basic reproduction number R*_*0_e*_. If *R*_*0_e*_ is greater than 1, the curve is similar in shape to an uncontrolled epidemic, but is “flattened”, with cases spread out more over time and fewer cases at the peak, with a greater degree of flattening when the value of *R*_*0_e*_ is closer to 1 (panels A1, B1, D1 and E1). The curve peaks when the number of people infected is 1–1/*R*_*0_e*_; this *effective herd immunity threshold* is lower than the “full” or “natural” herd immunity threshold, 1–1/*R*_*0*_, required to constrain the disease, through infection and/or vaccination, in the absence of control measures *[17]*.

If social distancing and hygiene measures push *R*_*0_e*_ to values less than 1, the case curve is “crushed” or “suppressed”; the initial exponential growth in case numbers is halted and cases decline (panel F1).

Strict quarantining of new arrivals and testing & tracing can keep the disease completely out of a country or territory or stop the disease from spreading beyond a few initial cases (panel H1). In such cases, control of the disease does not depend on the value of *R*_*0_e*_ because the disease is controlled by keeping out the “index cases” from which an outbreak can spread or quickly tracing and isolating the few contacts of any index cases that occur.

The remaining panels in figure 1 show what is expected to happen if control measures are relaxed after a period of time, as opposed to the assumption in panels A1-D1 and G1-H1 that the control measures remain in place throughout.

After an uncontrolled epidemic, there should be no further outbreaks (panel O) – unless immunity wanes and people can be infected and can transmit the virus again (the potential effects of which are discussed later). Vaccination of more than 1–1/*R*_*0*_ of the population (with a vaccine that has 100% efficacy) would also constrain future outbreaks.

If the case curve is flattened but the epidemic still continues until it is halted after reaching a reduced herd immunity threshold, additional smaller outbreaks can happen if control measures are subsequently relaxed. Such outbreaks will be limited if the curve was flattened to a moderate degree (panel B2) and could be completely absent if the final infection level exceeded the “natural” herd immunity threshold (panel A2). Later outbreaks can be more severe if the curve is flattened more and if the relaxation of measures takes place before cases have fully declined from the peak (panel D2). If the curve is flattened significantly due to getting *R*_*0_e*_ close to but still above 1, relaxing measures can lead to an almost full-scale outbreak, because a substantial proportion of the population is still uninfected (panel E2).

If the disease is suppressed initially, and control measures are subsequently relaxed, then the disease can spread again. Panel F2 illustrates the scenario where control measures are re-imposed and *R*_*0_e*_ is reduced to below 1 after each resurgence in cases. Panel C shows the expected evolution of cases for the scenario in which the disease spreads widely after *R*_*0_e*_ is allowed to increase above 1. Panel G shows the scenario where, at the present time, cases are increasing or just at a peak, but for which the future path may be uncertain. In these scenarios, if current or future control measures are sufficient to push *R*_*0_e*_ below 1, case numbers will decline (similar to the situation in panel F2); however, if *R*_*0_e*_ remains above 1, then case numbers will continue to increase until the epidemic is constrained by declining susceptibility levels (similar to the situation in panel C).

If the disease succeeds in taking hold in a country which had previously kept it out through strict quarantines and testing & tracing, the subsequent disease dynamics will depend on the value of *R*_*0_e*_. If measures are imposed that push *R*_*0_e*_ below 1, the disease will be suppressed, and the case curve will look like that in panel F1 but with the starting point of the epidemic shifted to later dates – as illustrated with the solid line in panel H2. If *R*_*0_e*_ remains above 1 after the virus enters and begins to spread in a country, then cases will follow the patterns in panels A1, B1, D1 or E1 (depending on the value of *R*_*0_e*_), with the starting point of the case curves shifted to later dates – as illustrated with the dashed line in panel H2 (which is similar to the case curve in panel E1).

In all these scenarios, people are assumed to become immune after infection and to retain immunity. In practice, for the SARS-CoV-2 virus, immunity appears to wane over time, and new variants of the virus are emerging some of which may have the ability to evade immune responses. In addition, new variants of SARS-CoV-2 have emerged which can be transmitted from person to person more readily than the original form of the virus; such variants will increase *R*_*0_e*_ even if there are no changes to the disease control measures implemented by governments and people. Waning immunity and new variants of the virus could lead to significant differences in the evolution of cases from any of the scenarios shown in figure 1 – as discussed later.

## 3 Methodology to Categorize COVID-19 Outbreak Dynamics for Different Countries

By expanding on the phenomenological reasoning employed both by Okell et al. *[15]* and in our previous work *[14]*, we can use reported data on COVID-19 cases and disease control measures, to elucidate and categorize, for most countries and territories (hereinafter called “countries” for simplicity), the dynamics of the disease to date in that country.

Figure 2 presents the possible categories and sub-categories of COVID-19 outbreak dynamics, together with key features and characteristics of each category.

**Figure 2:**
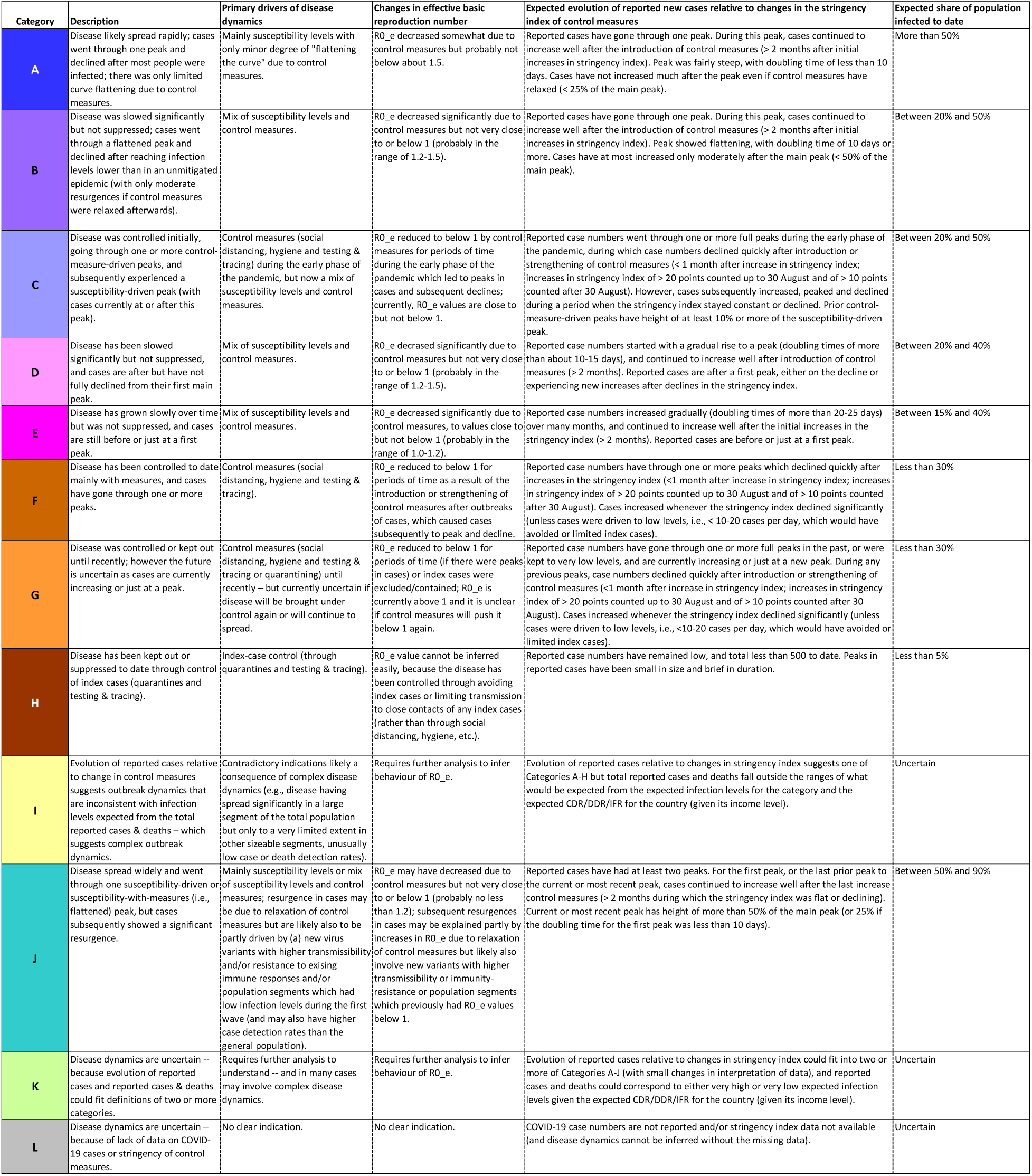
Characteristics expected for COVID-19 reported cases and deaths for different possible categories of outbreak dynamics. Table presenting possible categories of disease dynamics together with (a) expected evolution of reported new cases relative to changes in the stringency index of control measures and (b) expected range of possible infection levels (i.e., total share of population infected to date).

Two steps are used to identify the appropriate category for each country.

First, we study the evolution of reported daily new cases over time, and compare the timing of increases and/or decreases in reported new cases with changes in the “stringency index” of control measures compiled by the Oxford COVID-19 Government Response Tracker *[1]*. By matching the observations with the characteristics in evolution of reported cases expected for each possible category, from figure 2, we can identify the most appropriate category (or, in a few instances, possible categories) for each country.

Second, we take the infection levels expected for the category or categories indicated in the first step, and calculate predicted ranges for reported cases and deaths using plausible ranges for the case detection rate, death detection rate and infection fatality ratio (IFR), and compare these estimates with the actual reported cases and deaths. For most countries, the first step produces one category and the second step validates or rejects it. For a few countries, mostly ones for which there is no stringency index data, there are multiple possible categories from the first step, and only one category is consistent with the estimates calculated in the second step. Plausible ranges for the case detection rate, for countries of different income levels, come from comparing the results of serological surveys with reported cases from the same time periods *[18]*, and plausible ranges for the death detection rate come from comparing the estimates of excess deaths with reported deaths *[19]*. Overall IFRs for countries are estimated from age-specific IFRs *[20]* and the country’s population by age; these estimated IFR values are multiplied by 0.2–1.0 to create a range of possible IFR values, in light of the possibility that IFRs are likely to have declined substantially over time due to improvements in treatment of COVID-19 cases, especially in high- and middle-income countries, and that IFRs in some countries may be lower due to prior exposure to coronaviruses or other factors.

In figure 2, the Categories A to H correspond to the panels presented in figure 1. These categories and sub-categories fall into five main groupings, according to the dominant drivers of the disease dynamics.

- **Susceptibility-Driven Dynamics with Limited Mitigation – *Category A*:** Disease has probably infected large numbers of people and declined mainly due to low susceptibility levels (with little curve flattening due to control measures). Case numbers grew even after control measures were introduced (or strengthened), case numbers subsequently peaked and declined, with at most minor resurgences even if control measures were relaxed after cases declined. Total infection level is high, likely above 50%.
- **Susceptibility-Driven Dynamics Mitigated by Measures – *Categories B and C*:** Disease has probably infected large numbers of people and declined mainly due to low susceptibility levels, but with a curve that was flattened due to control measures. Case numbers grew even after control measures were introduced (or strengthened), case numbers subsequently peaked and declined, with at most moderate-scale resurgences if control measures were relaxed after cases declined. Total infection level is high but lower than if the outbreak were unmitigated, likely in the range of 20-50%.
- **Susceptibility-plus-Measures-Driven Dynamics – *Categories D and E*:** Disease has been slowed significantly but not suppressed; outbreak dynamics have been determined by a combination of susceptibility levels and control measures. Case numbers have grown for two months or more and are still growing or have reached a peak but not yet declined fully. Total infection level is moderate, likely in the range of 15-40%.
- **Measures-Driven Dynamics – *Categories F and G*:** Disease has been constrained to date mainly through control measures (social distancing, hygiene and testing & tracing). Case numbers declined very quickly after control measures were introduced (or strengthened) and/or increased again after measures were relaxed. Total infection level is low, likely well below 30%.
- **Index-Case-Control Dynamics – *Category H*:** Disease has been kept out or suppressed to date through strict control measures (especially quarantines and testing & tracing). Case numbers and infection levels were zero or very low, at least until recently.

There are four additional categories in figure 2. Categories I and J describe situations that are exceptions to the taxonomy of possibilities presented in figure 1 and discussed in the previous section, while Categories K and L capture situations where it is not possible to reach clear conclusions.

- **Inconsistencies between Evolution of Reported Cases and Estimates for Total Infection Levels – *Category I*:** Evolution of reported cases relative to timing of changes in the stringency index suggests one of Categories A-H but the total numbers of reported cases and/or deaths are inconsistent with the infection levels expected for that category – which would be an indication of more complex dynamics, i.e., large variations in *R*_*0_e*_ or case detection rates across regions, population segments or over time.
- **Resurgences after Susceptibility-Driven Peaks – *Category J*:** Case numbers experienced an apparently susceptibility-driven peak but subsequently showed a resurgence larger than what should be possible given the numbers likely to have been infected during the first peak, and hence would be an indication of more complex dynamics, such as low infection rates in large population segments during the first peak or emergence of immunity-evading strains of the virus.
- **Uncertain Dynamics Due to Ambiguities in Evolution of Reported Cases – *Category K*:** Findings about disease dynamics are uncertain because evolution of reported cases and total numbers of reported cases and deaths could fit the characteristics of two or more of Categories A-J.
- **Uncertain Due to Insufficient Data – *Category L:*** Disease dynamics cannot be ascertained because there is no data on cases and/or on the stringency index (and, the latter situation, the shape of the case curve and the total numbers of reported cases and deaths is insufficient to reach a conclusion about disease dynamics).

## 4 Each of the Main Categories of Possible COVID-19 Disease Dynamics Has Occurred in Practice

Figure 3 presents the findings on COVID-19 status categories for all countries and territories. Figure 4 shows the numbers of countries and territories in different categories, as well as the numbers of people living in countries and territories in each of the categories. Figures 5 and 6 provide illustrative examples of the reported case data and the diagnostic findings for a selection of countries. Charts showing reported cases and stringency index for all countries and territories are included in the Annex, grouped by COVID-19 status category.

**Figure 3:**
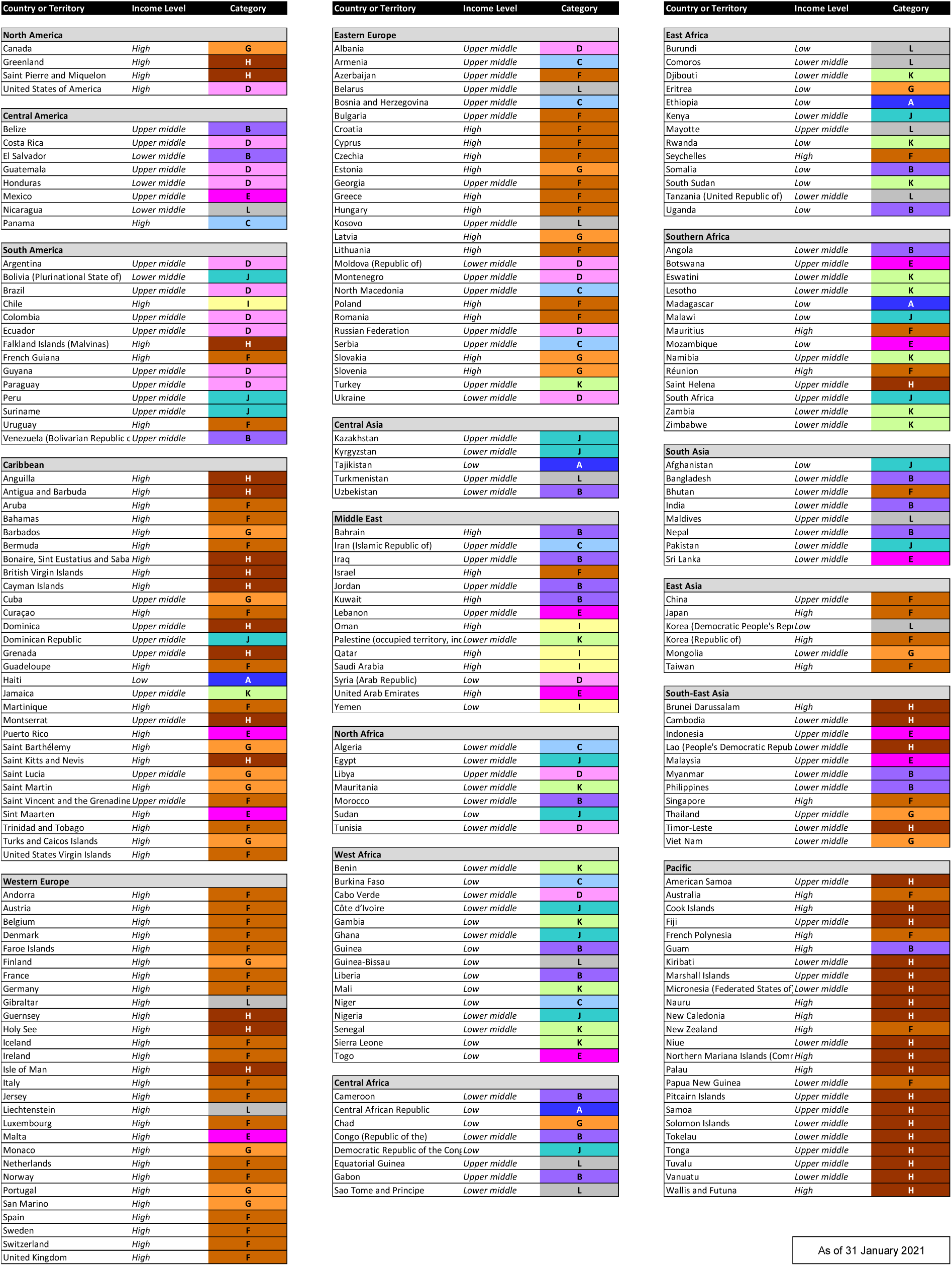
COVID-19 status category for each country and territory, as of 31 January 2021. The categories are defined and described in figure 2.

**Figure 4:**
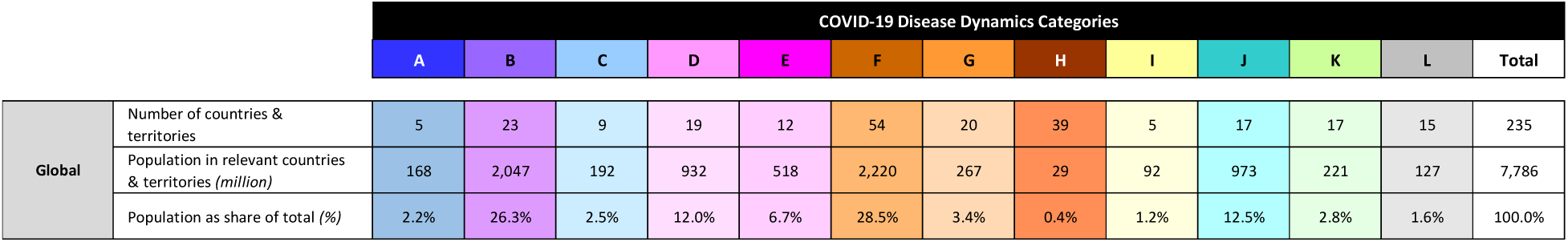
Numbers of countries and territories, and their populations, for each COVID-19 status category, as of 31 January 2021.

**Figure 5:**
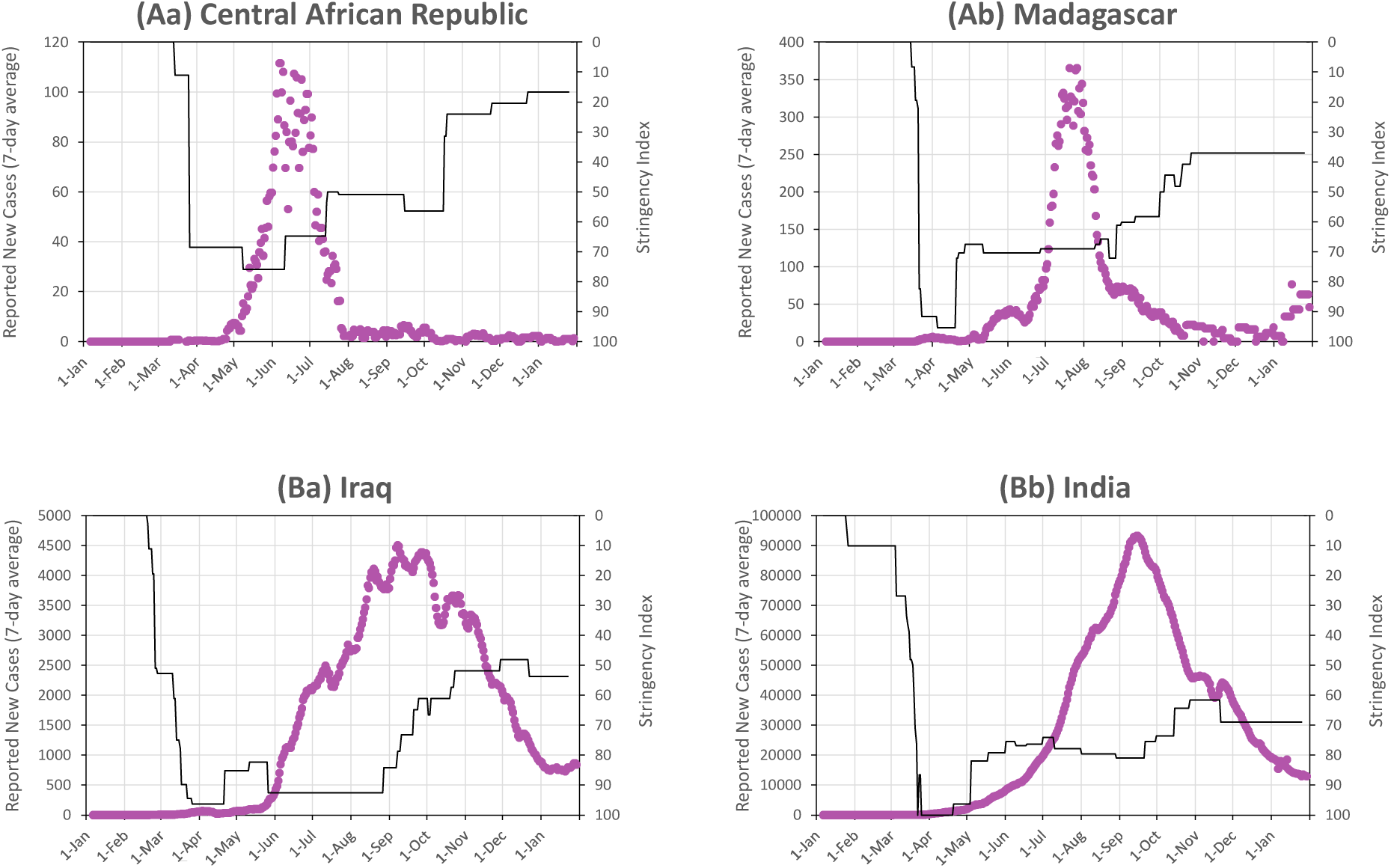
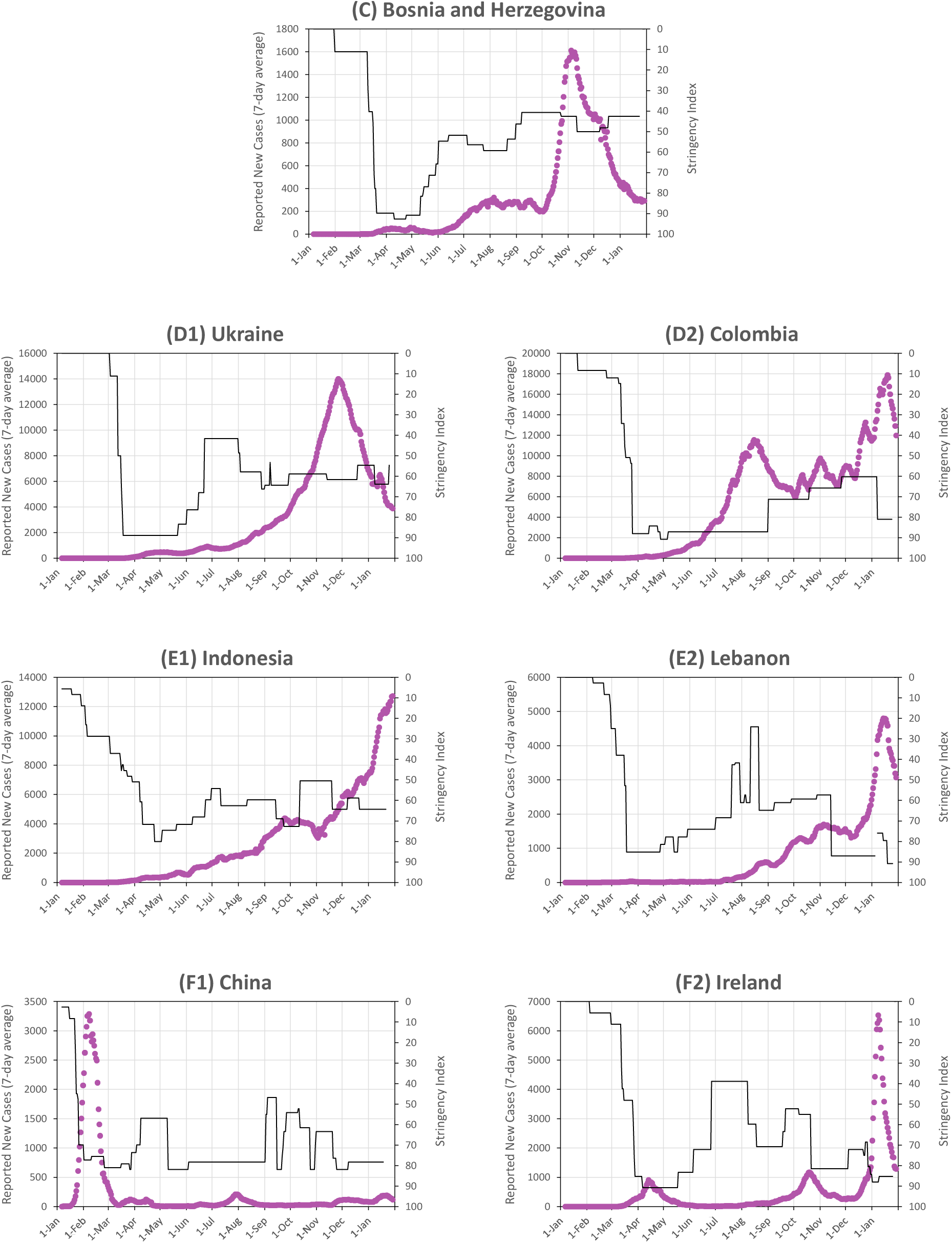
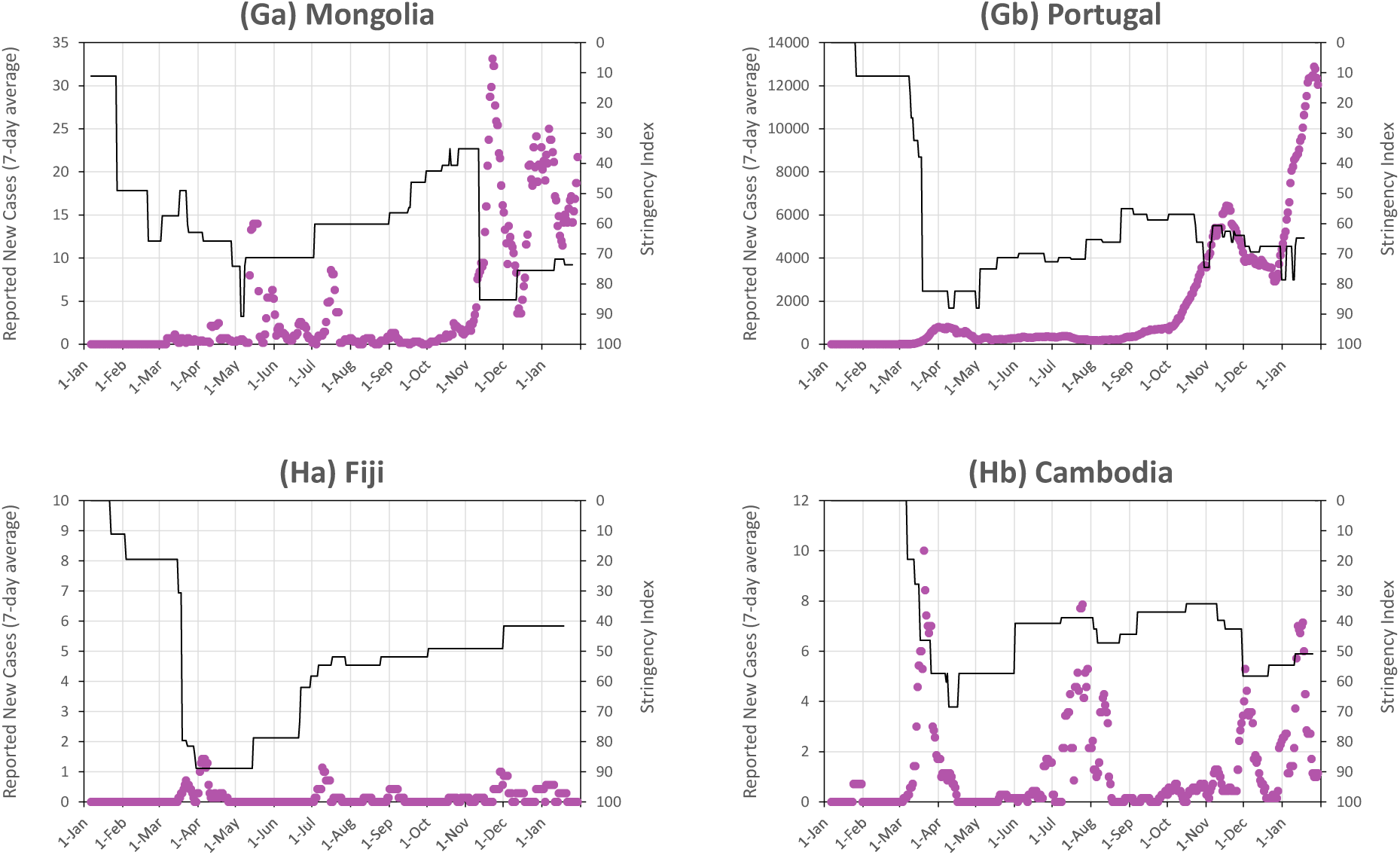
Reported cases and stringency index for selected countries, from each of the disease dynamics categories, up to 31 January 2021. (**A** to **H**) Selected examples of countries whose COVID-19 outbreak dynamics are in each of the categories described in the text and displayed in figure 1. The stringency index is shown on an inverted scale, for ease of comparison with figure 1 – although increases in stringency index will not necessarily produce proportional reductions in the effective basic reproduction number *R*_*0_e*_.

**Figure 6:**
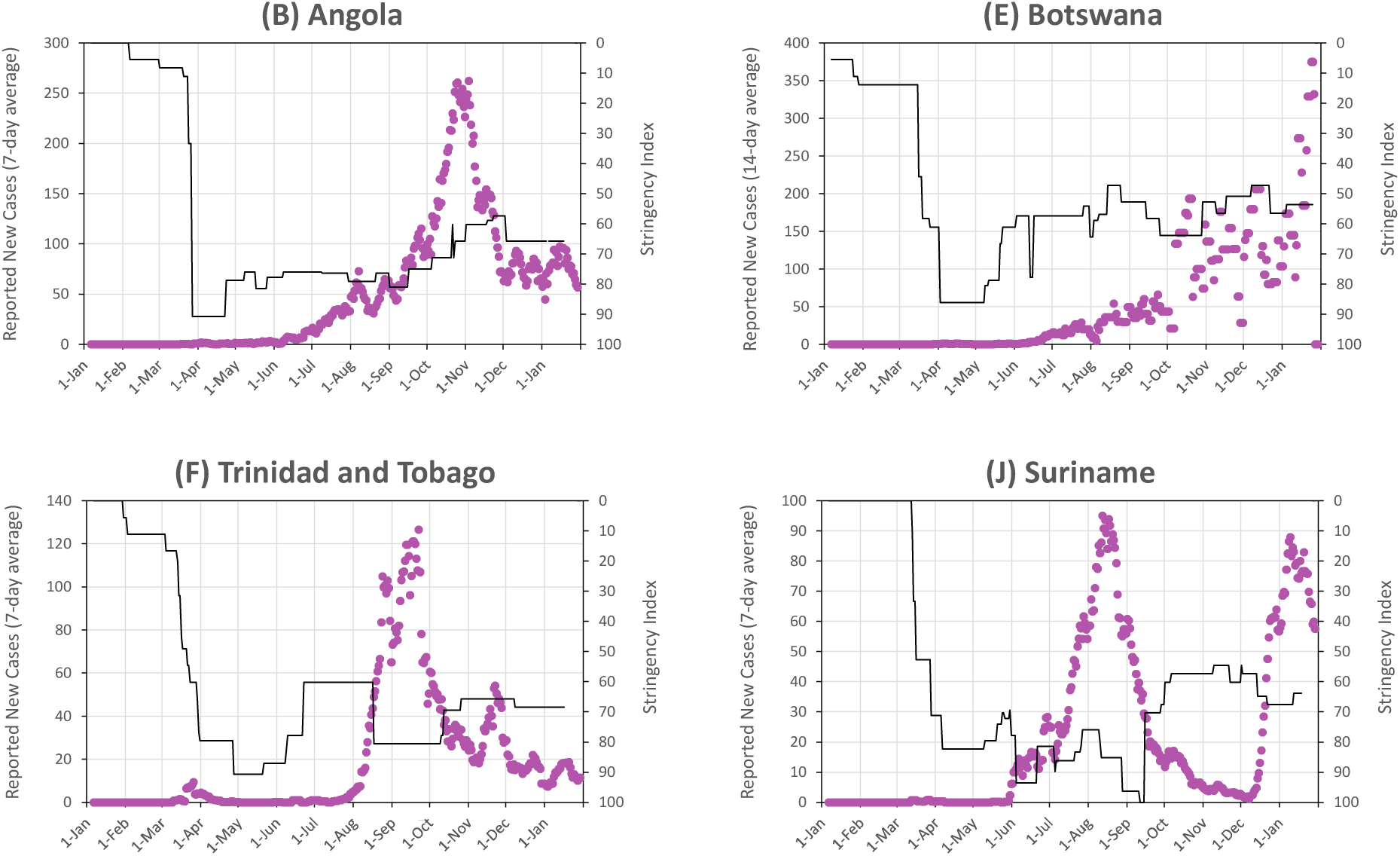
Reported cases and stringency index for selected countries in which COVID-19 was largely kept out up to 30 June 2020.

These figures show results and findings for 235 countries and territories *[21]* – often referred to as “countries” in this document for simplicity. They indicate that there is a wide variation in the state of the COVID-19 epidemic around the world. The data and the categorizations presented here show the state of the pandemic as of 31 January 2021; changes over time naturally cause countries to move between categories.

### Category A – Susceptibility-Driven Dynamics with Limited Mitigation

The evolution of reported COVID-19 cases shows characteristics of susceptibility-driven dynamics with only limited mitigation from control measures in 5 countries with about 168 million people (2.2% of the global population). For these countries in Category A, reported cases typically have narrow peaks with doubling times of less than 10 days, corresponding to values of *R*_*0_e*_ values of 1.4 or above (from the formula for the doubling time of *t*_*g*_ * *ln(2) / ln(R*_*0_e*_), where the mean generation time *t*_*g*_ is assumed to be 5 days *[22]*). Countries in this category include Central African Republic (for which data is presented in panel Aa of figure 5) and Madagascar (panel Ab).

### Categories B and C – Susceptibility-Driven Dynamics Mitigated by Measures

In a further 32 countries with about 2.24 billion people (28.8% of the global population), the COVID-19 epidemic also shows characteristics of susceptibility-driven dynamics, but the case curve was affected substantially due to social distancing and disease control measures.

Countries in Category B experienced substantial flattening of their case curves due to disease control measures, with doubling times of 10 days or more and *R*_*0_e*_ values which were typically below 1.4 but still above 1. There are 23 countries (with 2.05 billion people or 26.3% of the global population) in Category B, as of 31 January 2021, located mainly in Africa and Asia, and including Angola, Iraq (panel Ba), India (panel Bb), Morocco, and Venezuela, among others.

Category C includes countries which experienced susceptibility-driven peaks having previously controlled the virus through social distancing and other measures – such as Bosnia & Herzegovina (panel C). In all, this category included, as of 31 January 2021, 9 countries with 190 million people (2.5% of global population).

### Categories D and E – Susceptibility-plus-Measures-Driven Dynamics

In 31 countries, with 1.45 billion people (18.6% of global population), reported cases have increased slowly and have not yet either reached a peak or fully declined from their first peak, and estimated infection levels are moderate or high, indicating that control measures have had significant effects but not suppressed the virus.

Of these, the 19 countries in Category D have passed an apparent peak, mostly fairly recently, including, for example, Ukraine (panel D1) and Colombia (panel D2). A further 12 countries in Category E are before or just at a peak, including Indonesia (panel E1) and Lebanon (panel E2). For these countries, control measures significantly reduced *R*_*0_e*_, potentially to values falling in the range between about 1.3 and 1.05 (corresponding to case doubling times of between 13 days and 60 days if the mean generation time *t*_*g*_ = 5 days). As a result, the threshold levels beyond which cases have declined, or will decline in the future, was significantly reduced due to the control measures, and the numbers of people infected should be much lower than in an unmitigated epidemic.

Relaxation of control measures – or emergence of higher-transmissibility variants – in such circumstances, especially if it happens before cases have declined fully from the peak, can lead to substantial resurgences in cases. This pattern appears to have occurred already in several countries, including Brazil, Colombia and Paraguay (all in Category D) and Lebanon, Mexico, Mozambique, Togo and the United Arab Emirates (all in Category E).

Countries in Categories D and E will eventually transition to Category B or C once cases pass their peak and decline fully (unless changes are made which push *R*_*0_e*_ below 1 and “crush the curve”). Many of the countries in Categories B and C were previously in Categories D and E. India and Iraq, for example, would have been classified as Category E up to September and then as Category D until December, when they moved to Category B. Panama would have been classified as Category E until it recently moved to Category C. Ukraine, in Category D on 31 January 2021, will likely soon move into Category B.

### Categories F and G – Measures-Driven Dynamics

Disease control measures, including combinations of social distancing, hygiene, testing & tracing and quarantines in different combinations, have constrained the SARS-CoV-2 virus in 74 countries, with 2.49 billion people (31.9% of global population).

Category F includes 54 countries in which the disease has been suppressed through control measures to date and is currently under control. In 5 countries, outbreaks were suppressed in the first half of 2020 and the disease has since been kept largely out, including in China (panel F1) and New Zealand. In 11 countries, the disease was kept largely completely out at first, and outbreaks which happened in the second half of 2020 were suppressed – including, for example, Bhutan Georgia, Trinidad & Tobago and Uruguay. In 38 countries, control measures “crushed the curve” during two or more waves, including Ireland (panel F2) and many other European countries. This category covers a wide range of experiences, including countries with infection levels to date that are likely well below 10% as well as countries for which infection levels might be as much as 20% or more (for which the relatively high infection level can reduce the strength of control measures required to suppress further transmission of the virus).

Category G included – as of 31 January 2021 – 20 countries in which the disease was previously excluded – such as Mongolia (panel Ga) – or controlled – such as Portugal (panel Gb) – but in which reported cases are currently increasing. Many countries in Category G may bring their current outbreaks under control (and thus move to Category F, as has happened to several countries that were in Category G in December and to Portugal in the period since 31 January 2021), some may not (and thus move to Category C); the future evolution of the disease depends on whether *R*_*0_e*_ is or will be pushed below 1 by current or future control measures (which will require stronger measures in places with new variants of the virus that have higher “natural” basic reproduction numbers).

### Category H – Index-Case-Control Dynamics

COVID-19 appears to have been largely kept out, through strict quarantines and testing & tracing (together with lockdowns in a few cases), of all 39 countries that report fewer than 500 cases to date. These are almost all island states and territories (including Fiji shown in panel Ha of figure 5), except for Brunei, Cambodia (panel Hb of figure 5), Laos and Timor-Leste. The total population of countries and territories in Category H is 29 million or about 0.4% of the global population.

Some countries which largely kept SARS-CoV-2 virus out in the early months of the pandemic have subsequently experienced outbreaks, and some of these are shown in figure 6. In all, 82 countries and territories had fewer than 500 cases up to 30 June, of which 70 had fewer than 250 reported cases. Most of the countries which subsequently experienced substantial outbreaks managed to suppress the disease through control measures; 17 of them are in Category F (including Trinidad & Tobago shown in panel F of figure 6) and 9 in Category G (including Mongolia, previously shown in panel Ga of figure 5) as of 31 January 2020. However, 8 countries experienced outbreaks which have not been suppressed by control measures (and the situation in the other 9 countries is uncertain): as of 31 January 2021, 3 countries which kept the disease largely out up to 30 June 2020 were in Category B (Angola, shown in panel B of figure 6, Belize and Myanmar), 4 were in Categories D and E (Botswana, shown in panel E of figure 6, Guyana, Sint Maarten and Syria), and 1 was in Category J (Suriname, shown in panel J of figure 6).

### Category I – Inconsistencies between Evolution of Reported Cases and Estimates for Total Infection Levels

In 5 countries, the evolution of reported cases relative to timing of changes in the stringency index suggests one of Categories A-H but the total numbers of reported cases and/or deaths are inconsistent with the infection levels expected for that category. In all these countries, the evolution of reported cases suggests that the disease dynamics have been driven by susceptibility levels, but the total reported cases and/or deaths are lower than would be expected if most people had been infected, after accounting for plausible case detection and death detection rates.

For three of the countries – Oman, Qatar and Saudi Arabia – it is likely that large outbreaks have occurred within segments of the population – most likely segments of the large migrant worker populations in these countries – while lower infection levels prevailed in the rest of the population. Two other Gulf countries, Bahrain and Kuwait, are included in Category B2, but have reported cases that are close to the bottom of the plausible ranges if true infection levels are high; cases have started to increase again, likely due to the fact that large sections of the population remain uninfected to date.

Singapore, which is included in Category F, is another country in which the data suggests disease spread within a population segment – also migrant workers living in dormitories – but not much in the rest of the population. For Singapore, the case fatality ratio (CFR) is only 0.08 times the value of the infection fatality ratio (IFR) expected given the age-structure of Singapore’s population. The CFR may be reduced if many cases are undetected, but the very low value for Singapore suggests that deaths are also much lower than expected – a sign that the disease has likely spread mainly in a population segment with younger people than the general population.

Two other countries fall into Category I. For Yemen, it seems likely that the case detection rate is even lower than the rate of 1 in 2,000 assumed to be the lowest plausible level for LICs. For Chile, it is possible that fewer cases are being detected than expected for HICs, or that the virus is spreading mainly in a segment of the population, or that the evolution of reported cases is not due purely to susceptibility-driven dynamics.

### Category J – Resurgences after Susceptibility-Driven Peaks

In 17 countries, there were peaks in reported cases in mid-2020 characteristic of susceptibility-driven dynamics, but large resurgences of cases occurred after the end of the first peak. For some of these countries, such as Afghanistan and Pakistan, the first peaks appeared to be due to largely unmitigated outbreaks (similar to Category A), but resurgences have reached 25% or more of the maximum number of reported daily new cases during the first peak. For other countries, such as Egypt and Kenya, the first peaks were flattened to some degree (similar to Category B), but the resurgences have led to numbers of reported cases which exceeded 50% of the maximum case numbers during the first peak.

There are some clusters among the countries in Category J, both in geography and in the timing of the resurgences in cases. In South America, Bolivia, Peru and Suriname show steep surges in December-January. In South and Central Asia, Pakistan, Afghanistan, Kyrgyzstan and Kazakhstan show relatively slow-growing resurgences starting in September or October. In West Africa, Côte d’Ivoire, Ghana and Nigeria show steep surges starting in early or late December. In Southern Africa, Malawi and South Africa have resurgences starting in late December and late November, respectively. Egypt and Sudan experienced resurgences starting in October. Kenya, Democratic Republic of Congo and Dominican Republic experienced resurgences whose timing or shape don’t appear to match the evolution of reported cases in any of their neighbours.

For all these countries, the scale of the resurgences appears to be greater than would be expected given the share of population expected to have been infected during the first peak. Consequently, the resurgences are an indication of more complex dynamics – potentially including effects from new virus strains with higher transmissibility or ability to evade immune responses, from population segments having low infection levels after the first wave, or from waning of immunity acquired from infection in the first wave. These possibilities will be considered – to understand better the dynamics in Category J countries – at the end of section 5.

### Category K – Uncertain Dynamics Due to Ambiguities in Evolution of Reported Cases

There are 17 countries for which we classify the disease dynamics as uncertain because we cannot definitively allocate them to one of the earlier categories, since the evolution of reported cases could arguably fit with two or more of those categories and the total numbers of reported cases and deaths are consistent with each of the possible categories (considering the range of plausible values of the case detection rate and death detection rate for the country).

For 6 countries in West Africa (Benin, Gambia, Mali, Mauritania, Senegal and Sierra Leone) the first waves of cases continued growing for a month or more after the introduction of control measures before reaching a peak, and cases declined at the same time as the stringency index was declining or constant. On balance of probabilities, it seems likely that the peaks in cases were due to susceptibility-driven dynamics – with curve flattening due to measures, especially in Mali, Senegal and Sierra Leone. However, since the peaks in cases were reached within about 1-2 months after measures were introduced or the first cases appeared, there is a possibility that the peaks might have been due to control measures. In all 6 countries, cases have grown rapidly in the past 2-3 months, similar to the West African countries in Category J.

For 5 countries in Southern Africa (Eswatini, Lesotho, Namibia, Zambia and Zimbabwe), the first significant waves of cases reached peaks in July or August and cases declined to low levels by October. In most of the countries, the stringency index increased in July, but only by small amounts, and cases declined even as the stringency index was reduced in September. In all of these countries, there were surges in cases in December, and both the surges and the prior peaks around August are similar to those observed in South Africa and Malawi, which are in Category J.

For the countries in West Africa and Southern Africa in Category K, it is uncertain whether the second waves are due to spreading of the virus among uninfected people in the general population (similar to countries in Categories C and G) or due to resurgences generated by new variants or population segments with previously low infection levels (similar to countries in these regions that are in Category J). These possibilities will be considered at the end of section 5.

Two countries in the Eastern Mediterranean – Palestine and Turkey – appear to fall close to the boundary between Category C and Category F, and they have similar curve shapes to other countries in the Balkans and the Levant, some of which are included in Category C and some in Category F.

The remaining countries in Category K are Djibouti, Jamaica, Rwanda and South Sudan.

### Category L – Uncertain Due to Insufficient Data

Three countries – North Korea, Tanzania and Turkmenistan – do not track COVID-19 cases and deaths and do not provide reports to the World Health Organization.

Three countries – Belarus, Burundi and Nicaragua – had values of the stringency index which never exceeded 30 points, making it difficult to use the index to determine the strength of control measures.

In nine other countries and territories – the largest in population of which are Guinea-Bissau, Kosovo and Equatorial Guinea – there is no stringency index metric for the country, and, without this data, we could not categorize the disease dynamics reliably. (The Oxford COVID-19 Government Response Tracker does not have stringency index metrics for 55 countries, but 30 of these fall into Category H because they have fewer than 500 reported cases, and 13 more can be categorized based on the shape of the reported case curves and estimates of actual infection levels from total reported cases and deaths.)

## 5 Evolution of Reported Cases and Stringency Index Reveals COVID-19 Status for Most Countries – and Can Provide Early Warnings of New Variants

Our approach is simple. Compare the evolution of reported cases with changes in the stringency index, and check that infection levels are plausible given total reported cases and deaths and the income level of the country. Despite its simplicity, the approach is able to categorize the basic disease dynamics in most countries.

It is perhaps surprising that this simple approach should work. At first study, it would seem that many potential complicating factors should render national numbers of reported cases useless for understanding the disease dynamics. Reported cases represent only a small fraction of real cases, and this fraction will vary over time and across different regions or communities within a country. Variations in disease dynamics should happen over time and in different parts of a country, due to different degrees of social distancing, seasonal patterns, and so on. As described in this section, such patterns can be observed in the data – but they generate variations from the case curves expected if the disease dynamics were simple and populations were fully homogeneous, rather than obscuring the underlying dynamics in the observed data.

When complicating factors do cause reported case curves to deviate significantly from one of the simple forms in Categories A-H, the results may be particularly important for policymakers and public health leaders. For the countries in Categories I, J and K, the observations can indicate disease dynamics that are genuinely different in different population segments, or, perhaps most important, give early warnings of significant new variants – especially ones which can evade immune responses – in countries that do not have capacity to test for variants.

### Effects of variability of reproduction number and case detection on reported cases

Several factors can affect the evolution of reported cases, with the potential in theory to create patterns that might be misinterpreted as indicating one category of disease dynamics when in fact the disease is behaving very differently. First, the effective reproduction number, *R*_*0_e*_, can vary due to alteration of control measures, changes in compliance levels, or seasonal patterns. Second, there may be geographical differences in the onset date of the epidemic and wide variations in local values of *R*_*0_e*_. Third, case detection rates – the ratio of reported cases to real cases – can change as the availability of and regulations for testing change.

These complications do in fact affect the evolution of reported cases to some degree in all countries. They explain why no country has a smooth and simple reported cases curve like the simulation results shown in figure 1.

Figure 7 shows four countries where the evolution of reported cases likely implies deviations from simple assumptions of consistency in *R*_*0_e*_ and case detection rates across geography and time. Iran’s reported cases (shown in panel A) have increased, decreased and stayed roughly constant at various times, likely reflecting periods when the average *R*_*0_e*_ was above 1, below 1 and around 1.

**Figure 7:**
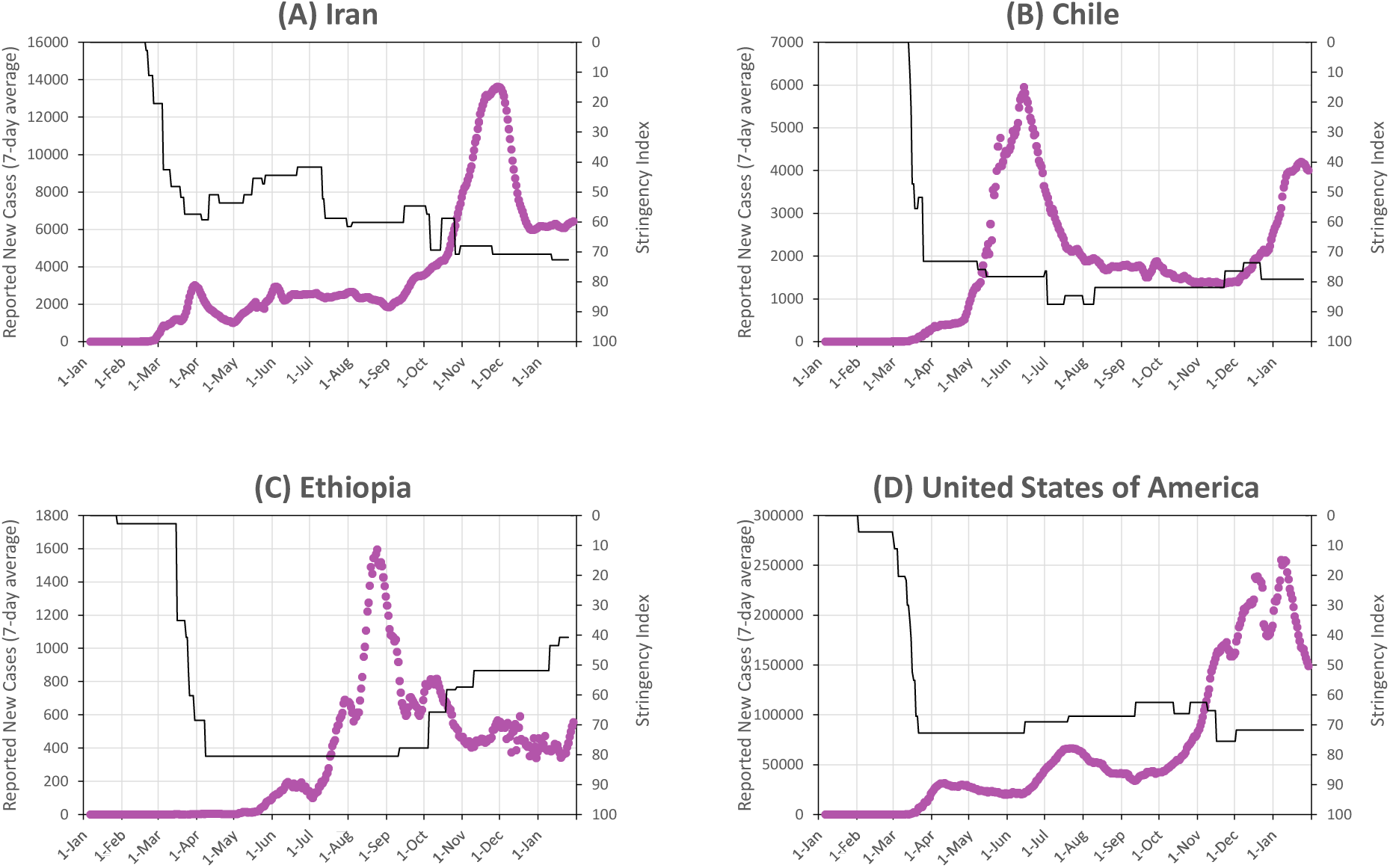
Reported case data and diagnostic findings for selected countries, which show features indicative of variations in reproduction number and/or case detection rates.

- Chile’s cases (panel B) show a single prominent peak in June, and the stringency index remained roughly constant before and after the peak – suggestive of susceptibility-driven dynamics. However, reported cases have experienced a ‘long tail’ and a resurgence since December. The Santiago capital region, which has 40% of the population, experienced a peak in cases around June. Cases peaked in northern regions mostly in July and August but experienced resurgences in December (perhaps related to the surge in cases across the border in Bolivia and Peru). Meanwhile, in most of southern Chile, which includes some of the least densely populated regions, cases continued to increase gradually throughout the year – suggesting that the disease arrived later in more remote regions and spread more slowly in less densely populated regions.
- Ethiopia’s cases (panel C) show a simple curve with a peak in late August, except for several undulations. These features are suggestive of changes in the case detection rate, and indeed some of the undulations correspond to notable dips in the number of tests reported in Ethiopia.
- Other countries, mainly large countries such as the United States of America (panel D), likely show the effects of variations, across geography and time, in both *R*_*0_e*_ and case detection rates.

In countries where *R*_*0_e*_ is usually close to 1, or with geographical regions or population segments that are strongly isolated from each other, it is likely to be important for policymakers to understand disease dynamics in detail to determine how *R*_*0_e*_ is changing over time and for different regions or population segments (in response to policy choices). However, our findings suggest that these complexities do not obscure the basic drivers of disease dynamics – in other words, which of our disease dynamics categories applies – for most countries.

The most significant assumption underlying the diagnostic tool is that the evolution of reported cases reflects roughly the evolution of actual cases, despite reported cases accounting for only a small proportion of all actual cases. Three considerations support this assumption. First, in most countries, the number of reported new cases has varied to a greater extent than the number of tests and the number of tests per confirmed case has varied substantially – even though one might expect the demand for tests to vary somewhat based on actual cases. Second, the numbers of reported deaths and reported cases follow a very similar pattern, with changes in reported deaths lagging changes in reported cases, even for countries with very low reported numbers of both – which suggests that changes in reported cases and deaths are genuinely reflecting the underlying disease dynamic. Third, for countries found to have high or medium infection levels (Categories A-E), the reported cases and deaths are a small fraction of the presumed actual numbers – but there is good evidence that case and death detection rates for such countries, mostly low- and middle-income countries, are in fact quite low *[3-13]*, and, indeed, our approach specifically checks to make sure that estimated infection levels (from reported cases and reported deaths) are not inconsistent with the disease dynamics suggested by the evolution of reported cases.

This is not to say that the case detection rate, the ratio of reported to actual cases, has not changed. It has. The number of reported deaths as a proportion of the number of reported cases – the case fatality ratio or CFR – declined in about three-quarters of countries in the second half of 2020 compared to the first half of the year, and by factors of 5-10 or more in many HICs. The reductions in CFR values are likely due to a combination of lower mortality rates, due to improved treatment of COVID-19 patients, and to an increase in the proportion of cases detected. These increases in case detection rate, however, were likely gradual and thus would not change any of the categorizations, which are principally based on changes in cases within a month or two of changes in the stringency index.

Another important assumption is that the evolution of reported cases is indicative of the evolution of actual cases in a country’s entire population. We have already found some countries where that assumption was not reasonable. For five Arabian Peninsula countries and Singapore, we found that reported cases might principally reflect the disease outbreak in one segment of the population. For some of the countries in Category J, it seems likely that resurgences in cases were due to disease spread, after relaxation of social distancing practices, in wealthier population segments which experienced lower infection levels than the rest of the population during the initial spread of the disease. For some countries, there are differences in disease dynamics in different regions (e.g., Chile and USA), and a few countries have unusually low case fatality rates which suggest that more younger people are being infected than older people (e.g., Singapore, Thailand, Martinique, Iceland and Réunion). However, aside from the countries just mentioned, there is little evidence to suggest that the disease evolution is markedly different for specific population segments (by geography or age or any other characteristic) to such a degree that the reported cases might not reflect fairly well the disease dynamics of the national population. Even in low-income countries, in some of which testing is disproportionately conducted in the capital cities or other large urban areas, it is likely that the reported cases do reflect the national picture: serology testing in India, Kenya and Nigeria *[18,23,24]*, for example, indicate that the disease spread quickly to rural areas after outbreaks started in urban centres.

### Effects of waning immunity

Evidence suggests that antibodies last for at least 3-4 months *[25]* but start to wane in some people for longer periods of time *[26,27]*, with the largest study, by the REACT Study Team, finding a decline in detectable IgG antibodies of 26.3% between 12 and 24 weeks after the peak in infections in England in April *[27]*. However, antibodies are only one component of the immune response to SARS-CoV-2, which also includes T cells (CD8^+^ and CD4^+^) and B cells. There is evidence that B cells *[28]* and T cells *[29,30]* can create strong immune responses to SARS-CoV-2 (with samples from about a third of people in studies in Germany *[29]* and Sweden *[30]* showing cross-reactive T cell responses to SARS-CoV-2 from prior exposure to other coronaviruses but not to SARS-CoV-2 itself). A recent study by J.M. Dan et al. *[31]* looked at five components of immune response (RBD IgG antibodies, RBD IgA antibodies, RBD memory B cells, total SARS-CoV-2-specific CD8^+^ T cells, and total SARS-CoV-2-specific CD4^+^ T cells) and found that “[b]y 5+ months after COVID-19, the proportion of individuals positive for all five of these immune memory compartments had dropped to 40%; nevertheless, 96% of individuals were still positive for at least three out of five SARS-CoV-2 immune memory responses”. They found wide variations in the magnitude of adaptive immune response across individuals, suggesting that a fraction of people may become susceptible to reinfection quickly after infection with SARS-CoV-2 (or vaccination).

These findings are consistent with observations of COVID-19 reinfections. In the most rigorous study to date, by the SIREN Study Group *[32]*, infection rates were tracked in a cohort of 6,614 UK public hospital staff with SARS-CoV-2 antibodies due to prior infection and compared to those in an antibody-negative cohort. The study found that “[a] prior history of SARS-CoV-2 infection was associated with an 83% lower risk of infection” – meaning that 17% of previously infected people were susceptible to reinfection – after a median observation time of 5 months following primary infection. The reduction in susceptibility to reinfection found by the SIREN Study is lower than the estimated decline in numbers of people with detectable IgG antibodies of 26.3% after 24 weeks. This finding is tentatively consistent with the idea that other components of the immune response to SARS-CoV-2, besides antibodies, can play a role in protecting people from reinfection.

The impact of immunity on disease dynamics, however, depends not on whether people become sick after reinfection with SARS-CoV-2, but on whether they contract the virus at all and whether, and to what extent, they transmit the virus to others. Only neutralizing antibodies provide sterilizing immunity, which stops a virus from replicating within the body and hence completely stops reinfection or risk of transmission to others. There is some evidence that T cell responses decrease viral shedding in other coronaviruses *[33]*, and hence it is plausible that such responses could decrease infectiousness of individuals. If future studies can provide estimates of the percentages of infected people who become susceptible to reinfection over time, and estimates of their likelihood to transmit the disease to others (compared to first infection), then models can simulate the likely impact on the dynamics of the disease in a population.

If immunity is lost over time, and people who were previously infected (or vaccinated) become able to contract and transmit the virus again, this could lead to future outbreaks. However, such outbreaks are likely to be significantly more modest in scale than the initial outbreaks when SARS-CoV-2 first spread during 2020, for several reasons. First, as discussed, people may retain partial immunity – with reduced susceptibility to reinfection compared to their first infection, and potentially a reduced likelihood of transmitting the disease to others. Second, of course, vaccines are likely to be available, which can continually replenish the overall level of immunity in the population.

### Effects of new virus strains with higher transmissibility or resistance to immunity

Many new strains of SARS-CoV-2 have emerged since the first variant of the virus started to circulate in human populations. Most of these have had similar characteristics to the original virus – causing similar disease symptoms and mortality, reproducing at similar rates, and generating and being controlled by similar immune system responses. However, late in 2020, at least three more concerning mutations have occurred. The B.1.1.7 variant, first identified in the United Kingdom and since found in more than 75 countries (as of 9 February 2021 *[34]*), has higher transmissibility, and is estimated to have increased the reproduction number in the UK by between 0.4 and 0.7 *[35]*. The B.1.351 variant, first identified in South Africa and since found in 32 countries (as of 9 February 2021 *[34]*), appears to have mutations which cause both higher transmissibility and decreased effectiveness of the immune system response in people who developed immunity to earlier variants (through infection or vaccination) *[36, 37]*. The P.1 variant, a descendant of the B.1.1.28 variant, first identified in Brazil and since found in 14 countries (as of 9 February 2021 *[34]*), has similar significant mutations to the B.1.351 variant, and thus appears likely to have higher transmissibility and resistance to existing immunity. It seems likely that more variants with immunity resistance and/or higher transmissibility will emerge in the coming months.

The emergence of new variants of SARS-CoV-2 with different characteristics would be expected to lead to deviations in the evolution of reported cases from the simple patterns expected in Categories A-H of figure 1. Consequently, keeping track of the trends in reported cases, compared to changes in the stringency of control measures, can help to identify such variants, especially in countries that do not regularly test for specific strains of SARS-CoV-2 in COVID-19 cases. The signatures of immunity-resistant and higher-transmissibility variants in reported case data should be different depending on the current status of the COVID-19 epidemic in a country.

#### New variants that evade immune responses

These variants allow people to be infected again even if they have immunity from past infection – all at the same time – although it is likely that the overall immune system response will still be able to mitigate the severity of symptoms from such new variants in most people.

- In countries with high to medium infection levels, i.e., those in Categories A-E, immunity-resistant strains can allow new outbreaks to occur among people who were previously immune, even after susceptibility-driven peaks.
- In countries with low existing infection levels, i.e., those in Categories F-H, immunity-resistant strains, if they have similar reproduction numbers to existing variants, will not have large effects on the case numbers.

Note that, although the effects of immunity-resistant variants on the potential for new cases may be low in countries in Categories F-H, immunity-resistant variants nevertheless have significant implications for all countries, because of their potential to reduce the effectiveness of current vaccines.

#### New variants that have higher transmissibility

Such variants have higher basic reproduction numbers *R*_*0*_ and thus results in higher effective basic reproduction numbers *R*_*0_e*_ even if disease control measures remain constant.

- For countries in Categories A, B and C, which have high or medium infection levels (above 20% at a minimum), higher-transmissibility strains may generate new outbreaks (if the increase in transmissibility is sufficiently great) by increasing the effective herd immunity threshold above the current infection level (as illustrated, for example, in figure 1, panels A2 and B2). New outbreaks should be more prominent for countries which have fewer people infected after their first peaks, i.e., those in Categories B and C. For countries in all of these categories, if there are distinct population segments in the countries with low infection rates from previous waves and low *R*_*0_e*_, new variants with higher transmissibility could push *R*_*0_e*_ above 1 and generate significant outbreaks within those segments.
- For countries in Categories D and E, which have medium infection levels (likely 15-40%), and where cases are before or just after the peak of a curve heavily flattened by control measures, new higher-transmissibility strains will increase *R*_*0_e*_ even if control measures are not relaxed, and generate new peaks (as illustrated, for example, in figure 1, panels D2 and E2).
- For countries in Categories F and G, which have suppressed the disease through social distancing and hygiene measures, higher-transmissibility strains are especially dangerous, because they can push *R*_*0_e*_ from below 1 to above 1, for a given level of control measures. This effect can be seen, for example, in the reported cases for the United Kingdom, Denmark, Ireland, Netherlands and Portugal, all of which increased sharply in December 2020 despite only small decreases in the stringency index (although it is likely also that social distancing practices during the holidays were relaxed to a greater degree than the stringency index might measure). Consequently, such strains require more stringent control measures to suppress the disease, compared to the original virus strains. In some countries, which previously were able to suppress the virus, it may prove impossible to suppress higher-transmissibility strains, in which cases the country would move from Category F to Category D or E (if they keep control measures in place that keep *R*_*0_e*_ close to 1) and eventually to Category B.
- For countries in Category H, which have largely kept the virus out, higher-transmissibility strains make this task more difficult, by increasing the chances that the virus make “break out” into the community following index cases. Such countries need to be even more vigilant in their quarantine and test & trace policies.

### Determinants of resurgences in cases observed in Category J and K countries (and indicators of new strains in some other countries)

Categories J and K include 28 countries in which reported cases have surged after first waves which were likely or possibly susceptibility-driven, with curves flattened to various extents as a result of control measures which mitigated the epidemics. In addition, there have been notable surges or accelerations in several other countries in South & Central America, in Northeast Africa, in West Africa and in Southern Africa besides the ones in Categories J and K. The complete list of countries to consider includes the following:

1. Kenya
2. Democratic Republic of Congo
3. Pakistan, Afghanistan, Kyrgyzstan and Kazakhstan
4. Egypt and Sudan
5. Southern Africa – Malawi and South Africa (in Category J); Eswatini, Lesotho, Namibia, Zambia and Zimbabwe (in Category K); Botswana and Mozambique (both in Category E but showing accelerations in reported new cases in January)
6. West Africa – Cote d’Ivoire, Ghana and Nigeria (in Category J); Benin, Gambia, Mali, Mauritania, Senegal and Sierra Leone (in Category K); Togo and Burkina Faso (in Categories E and C, respectively, but showing rapid growth in cases in December-January)
7. South and Central America – Bolivia, Peru and Suriname (in Category J); Brazil, Mexico, Argentina, Chile, Ecuador, El Salvador, Guatemala, Honduras (in other categories but showing substantial increases in cases in the November to January timeframe) Charts showing the reported cases and stringency index for selections from these countries are shown in figure 8.

**Figure 8:**
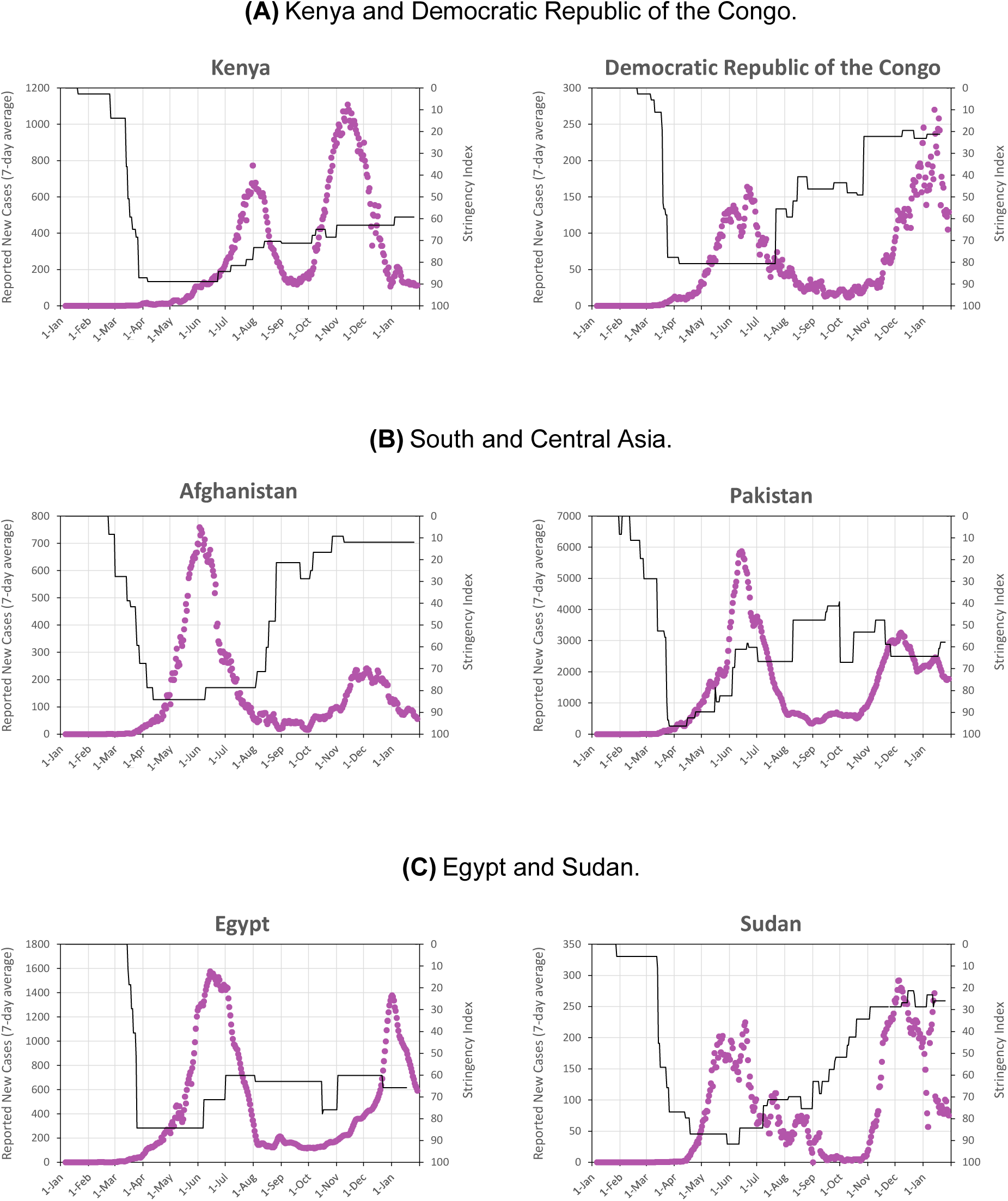
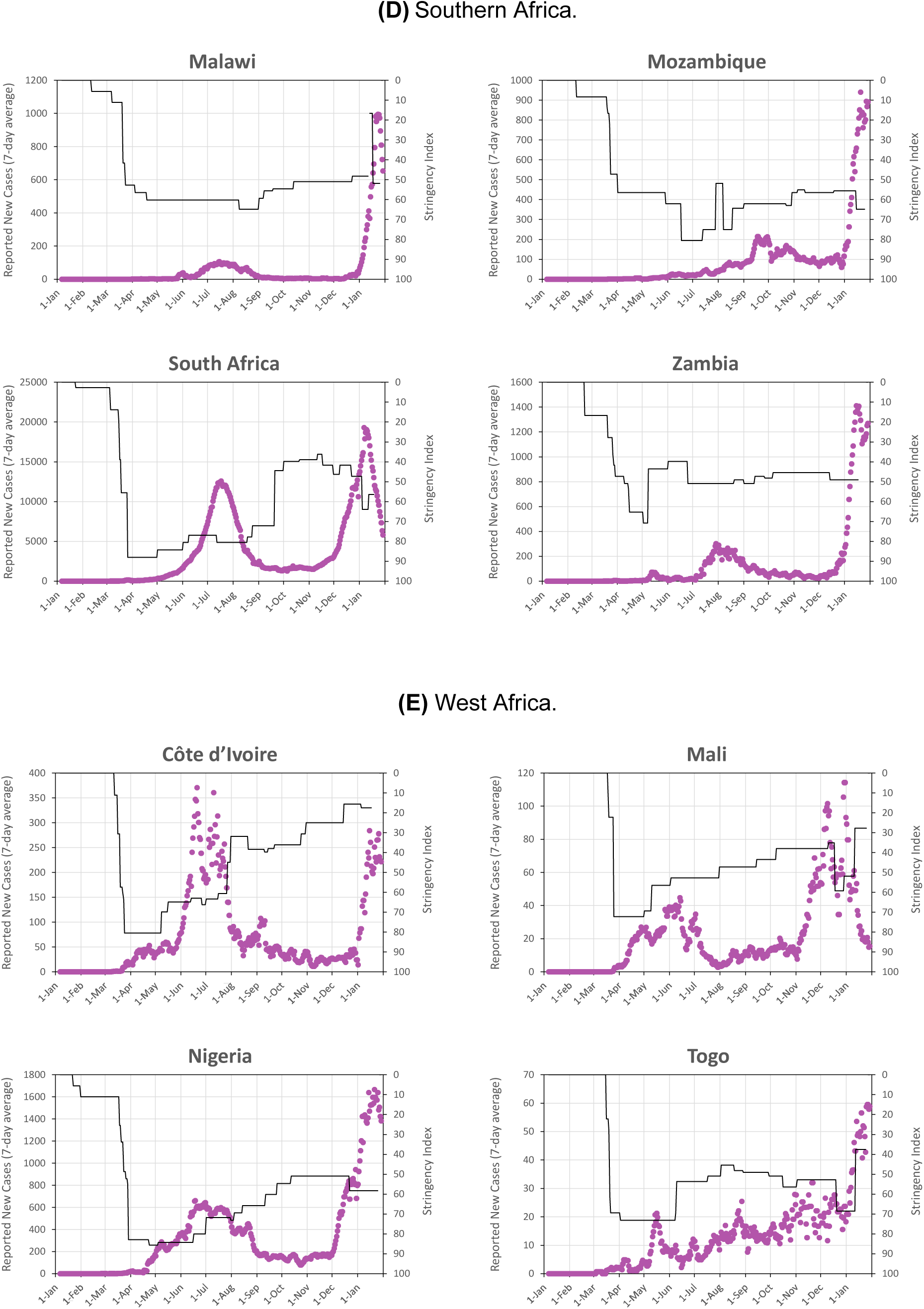
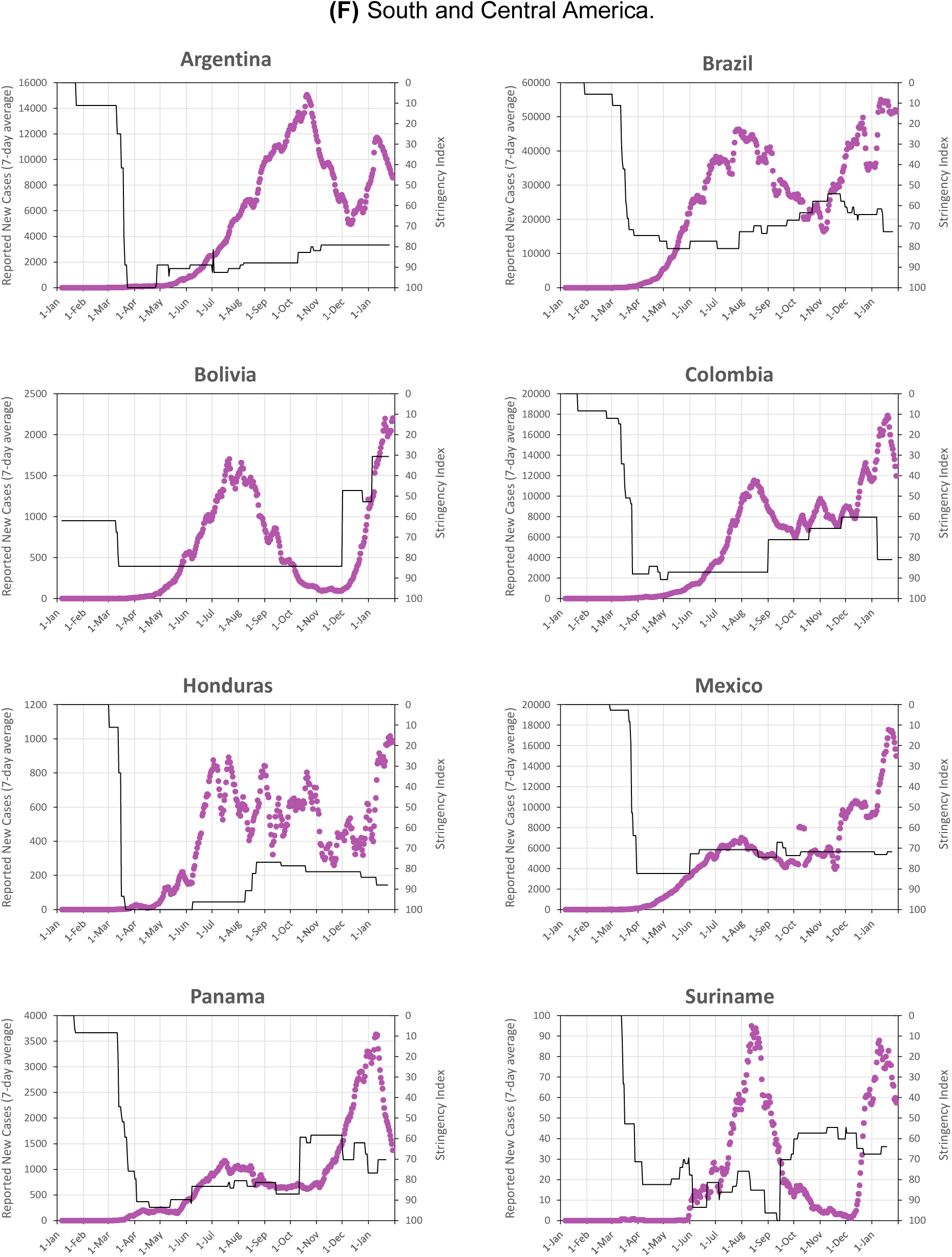
Reported cases and stringency index for selected countries which show features indicative of possible susceptibility-driven dynamics but with significant “second waves” after the first peak, or which show surges of accelerations in new reported cases. See text for discussion.

The resurgences and accelerations in new cases are potentially an indication of some or all of the more complex types of disease dynamics described in this section. To try to use the evolution of reported cases to diagnose the types of dynamics that are taking place, we consider five possible causes for resurgences after a susceptibility-driven peak that seem plausible:

a. Relaxation of control measures, leading to additional cases if the share of population infected at the end of the first peak is lower than the full herd immunity threshold when no measures are in place. (This scenario does not really constitute “complex dynamics”, but we consider it in order to understand what kinds of resurgences can be created purely by relaxation of measures after a susceptibility-plus-measures-driven peak.)
b. Waning of neutralizing immunity to the virus, which is expected to happen in some people after about 4-6 months.
c. Low infection levels in large population segments during the first peak, causing rapid spread of the disease after relaxation of measures within those segments. The effect on reported cases would be enhanced if the segments had higher-than-average case detection rates (as might be expected, for example, for upper- and middle-class communities in LMICs).
d. Emergence of new strains of the virus with higher transmissibility, if the effective herd immunity threshold with the new strain is increased to more than the share of population infected at the end of the first peak.
e. Emergence of new strains of the virus which are able to fully or partly evade immunity acquired from infection by earlier virus strains.

Figure 9 shows the evolution of reported cases that might be expected for each of these five possibilities, for scenarios in which *R*_*0_e*_ = 1.25 and 1.5 during the first peak (corresponding to doubling times of about 15.5 days and 8.5 days if *t*_*g*_ = 5 days), which would result in infection levels of about 38% and 59%, respectively, of the population after the first peak.

**Figure 9:**
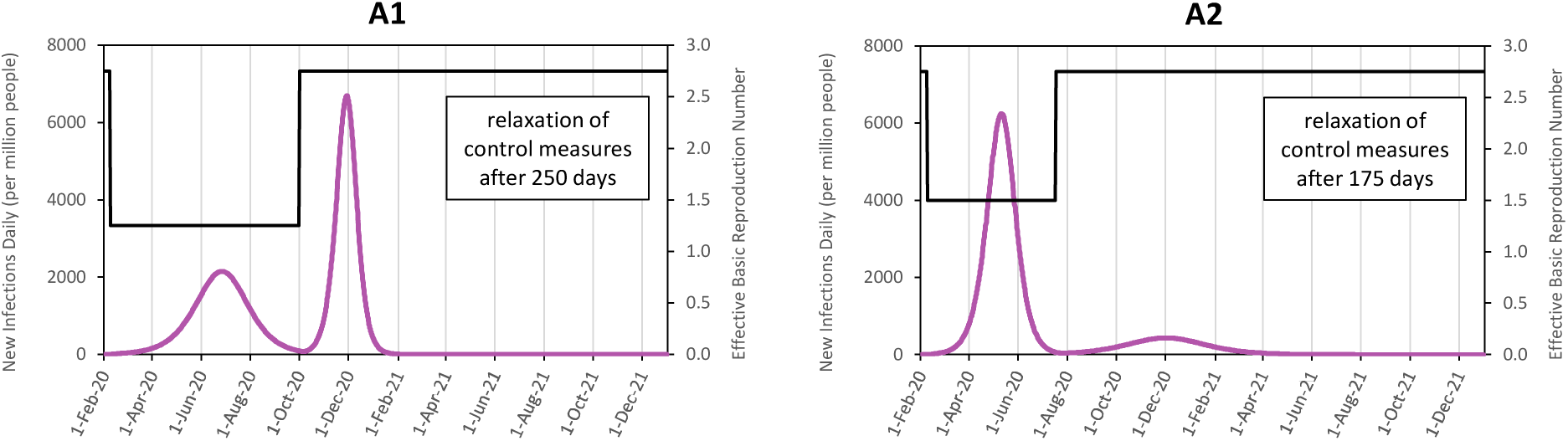
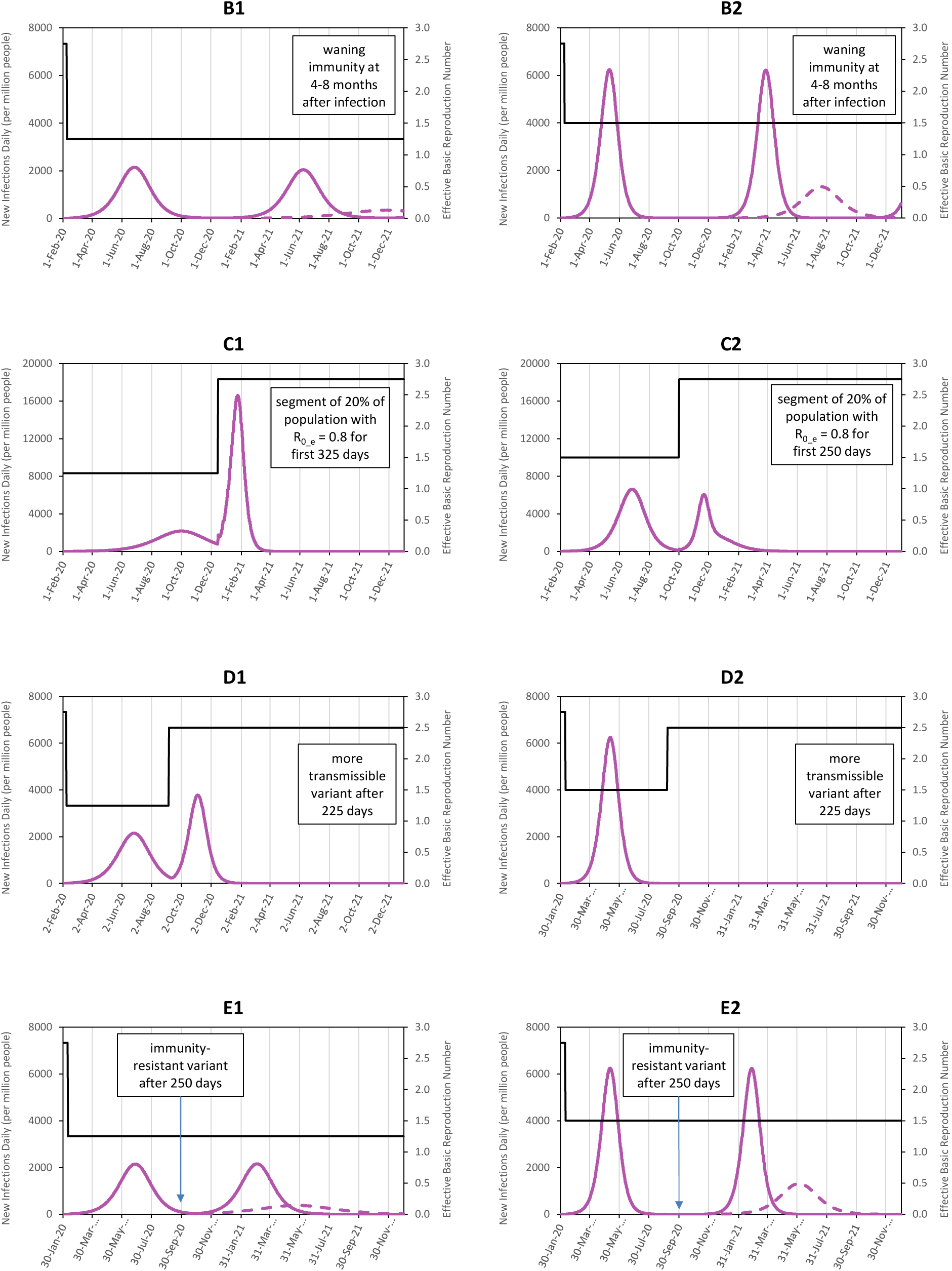
Evolution of numbers of disease cases in different scenarios following a susceptibility-driven outbreak with control measures. Calculated values for new daily cases per million people, from a simple SIR (susceptible-infected-recovered) model, under different assumptions. Each scenario starts with a complete outbreak during which control measures reduce the effective basic reproduction number to *R*_*0_e*_ = 1.25 (in the first column) or *R*_*0_e*_ = 1.5 (in the second column), resulting in infection levels of about 38% or 59%, respectively, by the end of the initial outbreak. (In all scenarios, the mean generation time is taken to be *t*_*g*_ = 5 days). Subsequent features of the scenarios are as follow: (**A**) Control measures are fully relaxed and *R*_*0_e*_ increases to *R*_*0*_ = 2.75 – after 250 days in panel A1 and after 175 days in panel A2. (**B**) Immunity wanes in the population, assuming a uniform distribution of immunity loss between 4 and 8 months after initial infection. The solid and dashed lines in these panels show the expected evolution when immunity wanes fully and wanes by 2/3, respectively. (**C**) Segment of the population, for which interactions within the segment are much more common than with the rest of the population, had a lower *R*_*0_e*_ = 0.80 during the initial outbreak but increases to *R*_*0*_ = 2.75 together with the rest of the population – after 325 days in panel C1 and after 250 days in panel C2. (**D**) Emergence of a new virus strain with higher transmissibility, increasing *R*_*0_e*_ to 2.5 after 225 days (both panels). (**E**) Emergence of a new virus strain which can evade immune responses, starting from 250 days. The solid and dashed lines in these panels show the expected evolution when the new strain is assumed to make all and 2/3, respectively, of the previously infected people susceptible to second infections. Note that the maximum value of the scale for new daily infections is greater for panels C1 and C2 than for the other panels.

The simulations indicate the circumstances which can generate larger, smaller or no resurgences ‐ specifically:

- Second peaks in cases can reach higher maximum values and be narrower than the first peaks when *R*_*0_e*_ increases due either to a relaxation of measures or emergence of a higher-transmissibility strain, and either (a) the first peak was substantially flattened or (b) a segment of the population, representing a substantial portion of the total population, experienced low infection levels during the first peak.
- Second peaks in cases which have lower maximum values and are broader than the first peaks are generated when there is waning immunity or immunity-resistant strains – unless the loss of immunity (to contracting and re-transmitting the virus) is complete (which causes second peaks that are similar to the first peaks), or there is simultaneously an increase in *R*_*0_e*_ due to relaxation of measures or virus strains which also have higher transmissibility.
- Second peaks, however, are not possible, or will only be very low and broad, if (1) the first peak was not significantly flattened, (2) there are no large population segments with low infection levels after the first peak, and (3) there is no significant reduction in population-level immunity due to waning or emergence of immunity-resistant strains.

The timing of second peaks depends, naturally, on the timing of the triggering events. In the case of waning immunity, the time difference between the first and second peak will always exceed the average time for people to lose immunity, and will increase beyond this lower bound for lower values of *R*_*0_e*_ and if population-level immunity loss is less than complete – for COVID-19, this means that waning immunity (alone) cannot generate a second peak in less than 6-8 months.

The results of the simulations suggest that some explanations are more or less likely to explain the resurgences in cases observed in each of the regions in which they have been observed.

In Kenya and Democratic Republic of the Congo, we note three features: (i) second peaks appear to have started following declines in stringency index in each country; (ii) neighbouring countries do not show rapid increases in cases around the same times; and (iii) the rates of increase of cases before the second peaks were somewhat faster, but not much faster, than before the first peaks. These features all suggest the explanation that the second outbreaks were driven by relaxation of control measures (cf. figure 9, panel A1), in which the virus spread among people who remained uninfected after the first, flattened peaks. It is possible that the second waves may have disproportionately affected upper- and middle-class population segments with lower-than-average infection levels at the end of the first peaks (cf. figure 9, panels C1 and C2).

In Pakistan, Afghanistan, Kyrgyzstan and Kazakhstan, the resurgences: (i) started around the time or soon after control measures were relaxed; and (ii) led to second peaks that were wider and lower than the first peaks. It seems like that declines in disease control measures led to the resurgences (cf. figure 9, panels A1 and A2). The second peaks are somewhat larger than might be expected from relaxation of control measures alone; however, they could be readily explained if case detection rates were higher for the second wave (which might have happened if, for examples, the second waves had a disproportionate effect on upper- and middle-class population segments).

Egypt and Sudan experienced second waves in October to January, following first peaks which appeared to be susceptibility-plus-measures-driven (i.e., Category B). The second wave in Sudan can be attributed to a large decline in stringency index, and the second peak is narrower that the first, as expected with a large increase in *R*_*0_e*_ (cf. figure 9, panel A1). The cause of Egypt’s second wave is less clear: Egypt’s stringency index declined only by a small amount before its second wave (although it is possible that social distancing may have relaxed to a greater extent in practice after the first outbreak abated) – and the second wave in Egypt had a slower doubling time than the first wave.

In Southern Africa, the main features to note are: (i) second waves happened in all countries in November or December; (ii) cases increased in countries irrespective of whether, and how much, the stringency index changed (including in Botswana and Mozambique which previously had experienced slow but steady trends in case numbers over time); and (iii) increases in cases were steep in all of the countries. These features point to the emergence of a new virus strain which has higher transmissibility – leading to rapid rates of growth in case numbers (cf. figure 9, panel D1) – and likely also resistance to immunity from prior infections – leading to sharp increases in countries with high prior infection levels as well as countries with low to moderate prior infection levels (cf. figure 9, panel E2, compared to figure 9, panel D2 which shows that higher transmissibility alone will not generate a large second peak in a country if large majority of people have previously been infected). Of course, it is known that the B.1.351 variant, first identified in South Africa, has both higher transmissibility and immunity resistance *[36,37,6]*, and that it has been responsible for most new COVID-19 cases in South Africa in the past 3 months. It is possible that relaxation of control measures and increased movements of people, especially around the holiday season in December, may have added to the pace with which the virus spread in some or all of these countries.

In West Africa, the main features have some similarities and some differences with those observed in Southern Africa: (i) second waves happened in most countries across the region, starting at different times between early November and January, but have not been observed in Guinea (and possibly not in Guinea-Bissau); (ii) cases increased in some countries after declines in the stringency index but without changes in other countries; and (iii) increases in cases after resurgences were rapid (with doubling times of less than 15 days) but similar to or slower than the increases during the first waves in some countries. The features suggest the presence of a new virus strain with higher transmissibility and possible resistance to immunity, but the evidence is somewhat less definitive than for Southern Africa. If there is a new virus variant circulating in West Africa, it is most likely to be the B.1.1.7 variant, which has been found in Ghana and Nigeria, but it is possible that the B.1.351 or P.1 or additional unidentified variants may also be present.

In South and Central America, Bolivia and Suriname experienced second waves which started in early December, and followed an upturn in cases in Brazil in early November. The second peak in Bolivia followed a significant decline in control measures, and could be explained by that alone (cf. figure 9, panel A1), because the curve was flattened substantially during the first wave. Similarly, the upturn in cases in Brazil followed a period during which the stringency index declined gradually. The second wave in Suriname started without any prior change in stringency index – and might be due to a higher-transmissibility variant (perhaps the P.1 variant that was first identified as coming from the Brazilian Amazon). Several other countries have experienced increases in cases from around the start of January – including Argentina, Chile, Peru and Colombia in South America as well as Mexico, Guatemala, Honduras and El Salvador in Central America. The simplest explanation for these increases is relaxation of social distancing behaviours over the Christmas and New Year holiday season (even if relaxations in practice are not reflected in the stringency index), although a role for virus variants cannot be discounted entirely.

## 6 Patterns in COVID-19 Status Have Emerged by Geography and Income

Figure 10 presents a map showing the COVID-19 status category for different countries and territories; see also figure 3 for information on status categories for small countries not discernible on the map. Figure 11 presents the numbers of countries and territories in different categories, by income and by region of the globe.

**Figure 10:**
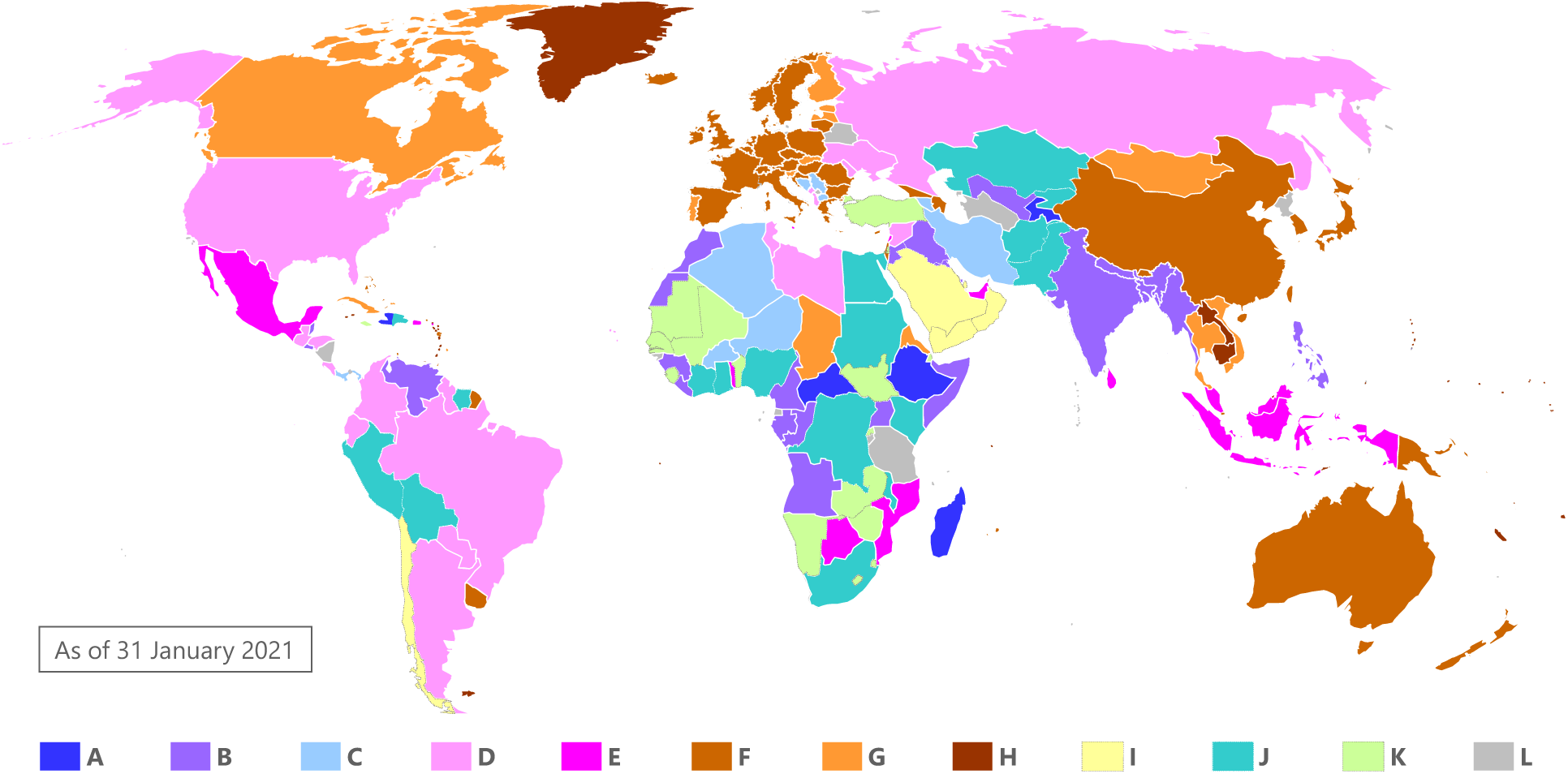
COVID-19 status of each country and territory, as of 31 January 2021. The categories are defined and described in figure 2.

**Figure 11:**
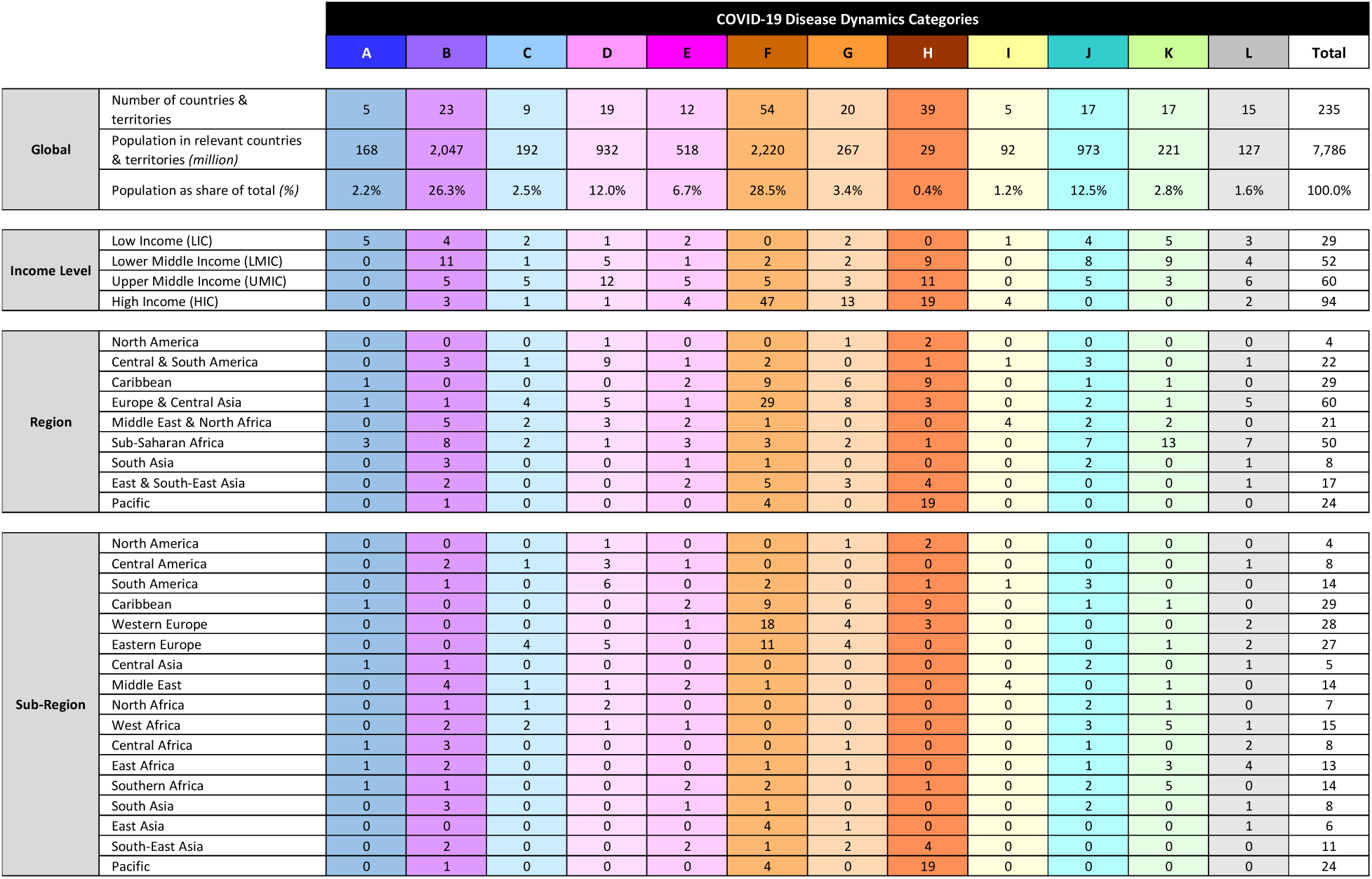
Numbers of countries and territories, from different income groups and regions, in each COVID-19 status category, as of 31 January 2021.

### COVID-19 epidemic status is correlated with income, but most categories include countries of all income levels

Countries of all income levels appear in nearly all the main disease dynamics categories. Nevertheless, there are clear correlations between income and COVID-19 status categories.

Most high-income countries have controlled the spread of SARS-CoV-2 through measures – accounting for 79 of the 113 countries in Categories F, G and H. Of the 15 HICs in other categories, the most populous is the United States of America, which had, as of 31 January 2021, “flattened the curve” but had not suppressed the virus spread, and five Arabian Peninsula countries in which the epidemic appears to have disproportionately spread in only parts of their populations.

Middle-income countries are spread across all categories. They account for 45 of the 63 countries which have slowed the disease significantly but not fully suppressed it (Categories B, C, D and E) – including populous countries such as India, Indonesia, Brazil, Bangladesh, Russia, Mexico and the Philippines. Furthermore, 25 out of 34 countries in Categories J and K are MICs – including Pakistan, Nigeria and Egypt – which have seen large resurgences in cases after susceptibility-driven peaks (or other uncertainties in categorizing the dynamics of their COVID-19 epidemics). Middle income countries also account for about half of the countries which have kept the disease out (Category H), and some others which have suppressed the disease to very low levels (including in Categories F and G) including several countries in South-East Asia, China, and many island states.

Some low-income countries have experienced largely unmitigated susceptibility-driven dynamics (7 of 29 countries, including countries in Categories A and some in Category J), while others have “flattened the curve” to varying degrees (11 countries in Categories B, C, D, E and J).

### Regional trends have emerged in COVID-19 epidemic status – but there are exceptions in each region

There are clear regional trends in the state of COVID-19 epidemics as of 31 January 2021. This is not surprising for three reasons. First, countries within a given region often have similar income levels. Second, the disease dynamics in one country is likely to influence in the dynamics in neighbouring countries, due to transmission of the disease across land borders. Third, governments and citizens look to neighbouring countries in determining disease control measures. There was more diversity in the state of the epidemic within regions earlier in the pandemic, but regional patterns have become clearer over time, especially as some countries which kept the disease out or under control during the early months of the pandemic have subsequently experienced more significant outbreaks.

In the Americas, the disease has spread slowly but has not been suppressed (Categories D and E), up to 31 January 2021, in most countries – including those with the largest populations, including the United States, Mexico, Colombia, Brazil and Argentina. Many of the Caribbean islands have kept SARS-CoV-2 out or under control (Categories F, G and H), but some have not, in particular Dominican Republic, Haïti, Jamaica and Puerto Rico. Several countries in South and Central America have showed prominent resurgences in cases, perhaps indicating the spread of a new variant with higher transmissibility and potentially also immunity resistance – including Bolivia, Peru and Suriname, in which cases had declined to low levels after susceptibility-driven peaks before increasing again (Category J).

Western and Northern European countries have, for the most part, controlled the disease through social distancing and hygiene measures, through two or three waves, and fall in Categories F and G. Some countries appear – based on estimates of excess deaths *[8]* as well as reported cases – to have reached infection levels well above 10% and perhaps beyond 20%, including Belgium, Spain and the United Kingdom, while other countries appear to have kept infection levels to low single-digit percentages, likely including Germany, Denmark, Finland and Norway.

Across Eastern Europe, the Levant, the Caucuses and Iran, all countries have constrained growth of the disease significantly, but infection levels in most have grown to moderate levels. The countries fall into three broad groups based on the COVID-19 status categories, although some countries likely lie at the boundary between categories. In many of countries, the virus was controlled or crushed during the early months of the pandemic. In the second half of 2020, case numbers grew substantially. In several countries in the Balkans, as well as Armenia and Iran, cases appear to have gone through peaks and declined due to rising immunity levels in the population (albeit likely at fairly low infection levels due to reductions in *R*_*0_e*_ caused by control measures) – and these countries fall into Category C. In other countries in Eastern Europe, as well as Georgia and Azerbaijan, increases in cases in the second half of the year were brought under control through measures – putting them in Categories F or G – although likely only after significant numbers of people were infected. For Belarus, Kosovo, Palestine and Turkey, it isn’t possible to determine whether Category C or Category F applies, and they are classified in Categories K or L. In some other countries in the region – including Moldova, Ukraine, Russia, Syria, Lebanon and Iraq – cases have shown, on average, a relatively steady increase over time, putting those countries into Categories D and E (or Category B in the case of Iraq).

In South and Central Asia, the virus has spread widely in most countries and cases have declined. In India, Bangladesh, Nepal and Uzbekistan, the case curve was flattened considerably, and current infection levels are likely moderate (Category B). In Pakistan, Afghanistan, Kyrgyzstan, and Kazakhstan (all in Category J), there have been two peaks in cases; the second peaks may have been generated purely due to relaxation of control measures, leading to spreading of the disease to previously uninfected people, but the possibility cannot be discounted that a new variant with some immunity resistance and/or higher transmissibility may have contributed to the second waves. Bhutan has contained the outbreaks of the virus to date (Category F).

Most countries in East and South-East Asia have largely kept the disease under control. Viet Nam, Laos, Cambodia, Thailand and Taiwan (all in Categories F or H) have largely kept the disease out – as has China in the time since it suppressed the initial outbreak of the disease in February and March of 2020. Malaysia, Mongolia and Myanmar also kept the virus until the second half of 2020, but have experienced widespread outbreaks since then. The disease has spread very slowly in Indonesia (in Category E) – which has one of the longest doubling times in the world – but has not been suppressed to date. The Philippines appears to be past the peak of its epidemic to date, and the curve was flattened significantly – meaning, like for other countries in Category B, that significant numbers of people have been infected but equally that sizable numbers remain susceptible to the disease (even before new variants are taken in account).

In Australia, New Zealand and most Pacific Island States, SARS-CoV-2 has been excluded through quarantines, together with testing and tracing and lockdowns when the virus has spread beyond quarantined individuals (Categories F and H).

There are significant differences in the status of COVID-19 epidemics across the African continent. Many countries appear to have experienced widespread epidemics followed by declines in case numbers. Disease spread in those countries was largely unmitigated in a few countries, but the curve was flattened in most. In some countries – Tunisia, Libya, Togo, Botswana and Mozambique – the effective reproduction number was reduced to very close to 1 and cases spread very slowly during 2020 (falling into Category D or E). A few countries – Eritrea as well as Chad (and possibly Niger and other Sahelian countries) – appear to have kept the disease out and suppressed small outbreaks, and a few, such as Algeria and Burkina Faso, may have experienced widespread outbreaks after keeping the virus under control during earlier outbreaks. As described earlier, several countries in Southern Africa, in West Africa, and in Northeast Africa experienced rapid growth in case numbers in December and January. Spikes in cases have occurred in countries which appeared to have previously experienced susceptibility-driven peaks (previously Category B, now Category J), in countries where the nature of earlier peaks was uncertain (Category K) and in countries with slowly rising numbers of cases (Category E).

## 7 Estimates of Global Infections and Deaths to Date, and of Potential Future Deaths

### Actual infections and deaths are likely much higher than previously estimated

For each country, we estimate the actual number of infections and deaths to date, based on the overlap between two plausible ranges. The first set of ranges comes from the total numbers of reported cases and deaths, and plausible ranges of case and death detection rates for the country’s income level. The second set of ranges comes from the expected range of infection levels for the country’s disease dynamics category, and the estimated infection fatality rate (IFR) for the country based on its population by age. Figure 12 shows these estimates for different groups of countries. Globally, the estimates suggest that roughly 1.3–3.0 billion people have been infected by SARS-CoV-2 to date, or about 17–39% of the population, which is between 13 and 30 times the number of confirmed cases, and perhaps twice to four times as much as previous estimates of total infection numbers *[38]*. The estimate for actual deaths to date is 4.6–10.0 million, between 2.1 and 4.5 times the number of confirmed deaths attributed to COVID-19.

**Figure 12:**
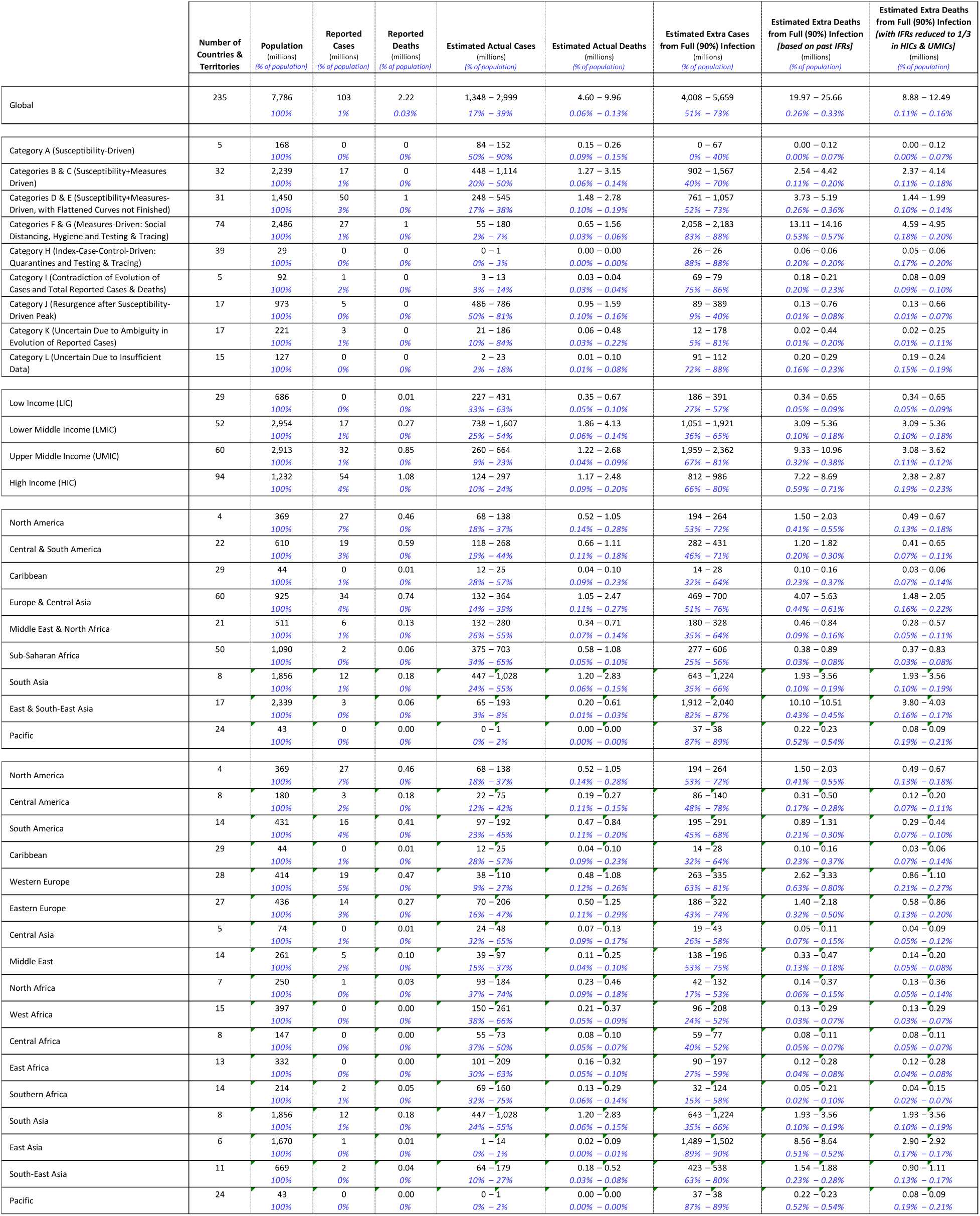
Estimates of actual and potential additional cases and deaths if 90% of the population were infected, for different groups of countries and territories. Estimates are shown for different disease dynamics categories, income levels and regions.

### High-income countries, China and India account for about two-thirds of potential extra deaths (before vaccinations or reinfections)

We next estimate the number of people who remain vulnerable to infection, assuming that up to a total of 90% of the population (somewhat higher than the proportion expected if *R*_*0*_ = 2.75 and the outbreak were completely uncontrolled) could be infected in the absence of control measures or vaccination. We also estimate the number of additional deaths, based on published estimates of age-specific IFRs *[20]* and the population by age in each country. There is some evidence that IFRs may have declined in recent months due to improvements in treatment of COVID-19 patients; it is also likely that health systems in LMICs and LICs have less capacity to handle COVID-19 cases and will be unable to provide the same standard of care. Consequently, we make a second estimate of potential additional deaths, by scaling the numbers for HICs and UMICs by 1/3. The estimates for different groups of countries also appear in figure 12. Not surprisingly, Category A/B/C countries have the lowest estimates for additional cases and deaths (as a share of population), while Category F/G/H countries have the highest estimates. The gaps between categories are even greater for estimated deaths, because Category A/B/C countries have younger populations than the global average and Category F/G/H countries have the oldest populations. Allowing for better COVID-19 treatment in richer countries reduces the effect because Category A/B/C countries are mainly LICs and LMICs while Category F/G/H countries are mainly HICs and UMICs.

Across the world, we estimate that if 90% of the population were infected, COVID-19 would cause an additional 8.9–12.5 million deaths allowing for reduced mortality in HICs and UMICs due to improvements in COVID-19 care (or 20.0–25.7 million deaths without such reductions in age-specific infection fatality ratios). HICs account for about 2.4–2.9 million of the potential additional deaths (assuming reduced mortality), China for about 2.1 million, and India for about 1.7–2.9 million (large numbers of people remain uninfected because India’s case curve was flattened significantly and reductions in infection fatality rates from improved COVID-19 care are not likely to apply). In all, HICs, China and India account for about two-thirds of the potential extra deaths that could occur if 90% of all people were infected (69% of the lower estimates and 63% of the upper estimates). Vaccinations, of course, will reduce these numbers substantially: as of 16 February 2020, HICs had administered about 40 million vaccine doses (which corresponds to 17% of the over-65 population of HICs, although some people have already received two doses, and not all vaccines have been administered to people over-65), China had administered about 41 million doses (corresponding to 3% of the over-65 population), and India had administered about 9 million doses (corresponding to 0.7% of the over-65 population). New disease variants that can evade immune responses (whether from prior infection or vaccination) could allow for infection rates of more than 90% of the population – but it remains to be seen whether the infection fatality ratios from reinfections will be comparable to those from first infections or much lower as might be expected.

## 8 Implications for Immunity Testing, Disease Control and Vaccination Strategies

Policies and actions to manage the COVID-19 emergency should be different in countries which currently have low, moderate or high infection and immunity levels. In this section, we present broad recommendations on immunity testing, disease control measures and vaccination strategies for different categories – recognizing that, even within categories, there will be variability across countries and territories which will require further tailoring of policies and programmes.

### Testing for total infection and immunity levels

It is important to get accurate information on the current state of the COVID-19 epidemic in various countries. Ideally, this would be achieved through immunity testing of representative population samples. It may be impractical for every country to implement serological testing programmes – but even studying a selection of countries across the different categories would help to test our conclusions that countries in Categories A-E and I-K (i.e., other than Categories F-H) have high to moderate infection and immunity levels, and the findings of this report could be applied to other countries which cannot deploy immunity testing quickly.

Of course, many serological studies have already been undertaken – and their estimates of infection and immunity levels are consistent with the findings of this study. Seroprevalence levels in several European countries, the United States, Canada and China all indicate low immunity levels *[18,20]*, while studies in several cities and regions in Brazil, India, Kenya, Nigeria, Pakistan, Qatar, and South Africa *[3-7,18,20]* reported finding antibodies for SARS-CoV-2 in large percentages of the studied populations.

Note that serological testing will underestimate the number of people who have been infected by SARS-CoV-2 once more than 4–6 months have passed, due to waning of SARS-CoV-2 antibodies. As discussed earlier, it is possible, therefore, that serological testing may understate the actual degree of immunity in a population, because some such people may have antibodies at levels below the detection threshold of the serology tests or may have memory B cell or T cell responses, either or both of which may reduce their vulnerability to infection and/or reduce their likelihood to pass on the virus to other people.

In some places, reliable estimates of actual deaths due to COVID-19 may be a substitute for immunity testing to determine the share of population infected to date, at least approximately. Such estimates have been made for many HICs and a few MICs – including Bolivia, Ecuador, Peru, Mexico and South Africa – by comparing total deaths in since the start of the COVID-19 pandemic with expected baselines based on deaths in recent years *[9]*. Estimates have been made using novel methods for several LICs, including for Sudan using a social media survey, a joint serological and molecular survey and modelling *[10]*, for Syria using community-uploaded obituary certificates and modelling *[11]*, for Yemen using geospatial analysis of burial surface areas in cemeteries *[12]* and for Zambia from a systematic post-mortem surveillance study *[13]*. By comparing the estimates of actual deaths with estimates of expected mortality rates, one can infer estimates for the share of population infected to date. These estimates infer low infection levels for 34 HICs, a wide range of infection levels for UMICs, and moderate to high infection levels for the LMICs and LICs studied *[8]*.

The implications of immunity levels on the disease will become complex to understand during the year ahead, for several reasons:

- Immunity from first infections will wane. More research is needed to determine to what extent people are at risk of reinfection after immunity begins to wane, how severe the symptoms from second infections will be, and whether reinfected people will transmit the virus at the same rates as people infected for the first time.
- People will gain immunity through vaccinations. More research will be needed to determine whether, and to what extent, vaccinated people are protected from contracting and re-transmitting the virus, and to determine for how long immunity from vaccines remains before waning. For countries in Categories A-E, which likely have high to moderate infection levels, the total numbers of immune people as vaccination programmes ramp up will depend on whether vaccinations can be targeted to people who have not already contracted SARS-CoV-2 or are distributed generally irrespective of prior infection.
- New virus strains will render previously immune people susceptible again. Several new strains, such as the B.1.351 variant first identified in South Africa and the P.1 and P.2 variants (descended from the B.1.1.28 variant) first identified in Brazil, appear likely to reduce immunity levels. However, it is likely that previously infected or vaccinated people will have partial immunity to these variants, reducing the severity of their symptoms and possibly reducing the degree to which they transmit the new strain to others. Further research on these variants, and any others that emerge, will be necessary.

### Disease control strategies

Countries in different categories ought to pursue different approaches to disease control. Figure 13 helps to illustrate the choices facing countries in different disease dynamics categories, by showing calculations of the expected total infections in the event of different decisions on relaxation of control measures. In both panels, the solid black squares and line show the total infections, for different values of the effective reproduction number *R*_*0_e*_ under control measures, if measures are not relaxed. In both panels, the maximum infection level is 92.4%, the share of the population infected if an epidemic with *R*_*0*_ = 2.75 is unmitigated; note that this is higher than the herd immunity threshold, which is 63.6%, because infections continue even after new case numbers start to decline. Panel A shows the total infection levels expected if control measures are fully relaxed (causing *R*_*0_e*_ to revert to *R*_*0*_), for different timings of the relaxation of control measures after the peaks in cases. Panel B shows the total infection levels expected if control measures are relaxed well after the peak in cases (200 days) but are only partially relaxed; black squares show infection levels in the scenario in which measures are not relaxed (prior to vaccination), while black ‘X’ marks show infection levels in the scenario in which measures are fully relaxed. In both panels, arrows indicate the outbreak dynamics categories (A, B/C, D/E, F/G) that roughly correspond to the initial values of *R*_*0_e*_ in the simulations – although this correspondence is not exact because *R*_*0_e*_ varies over time for most countries in most categories.

**Figure 13:**
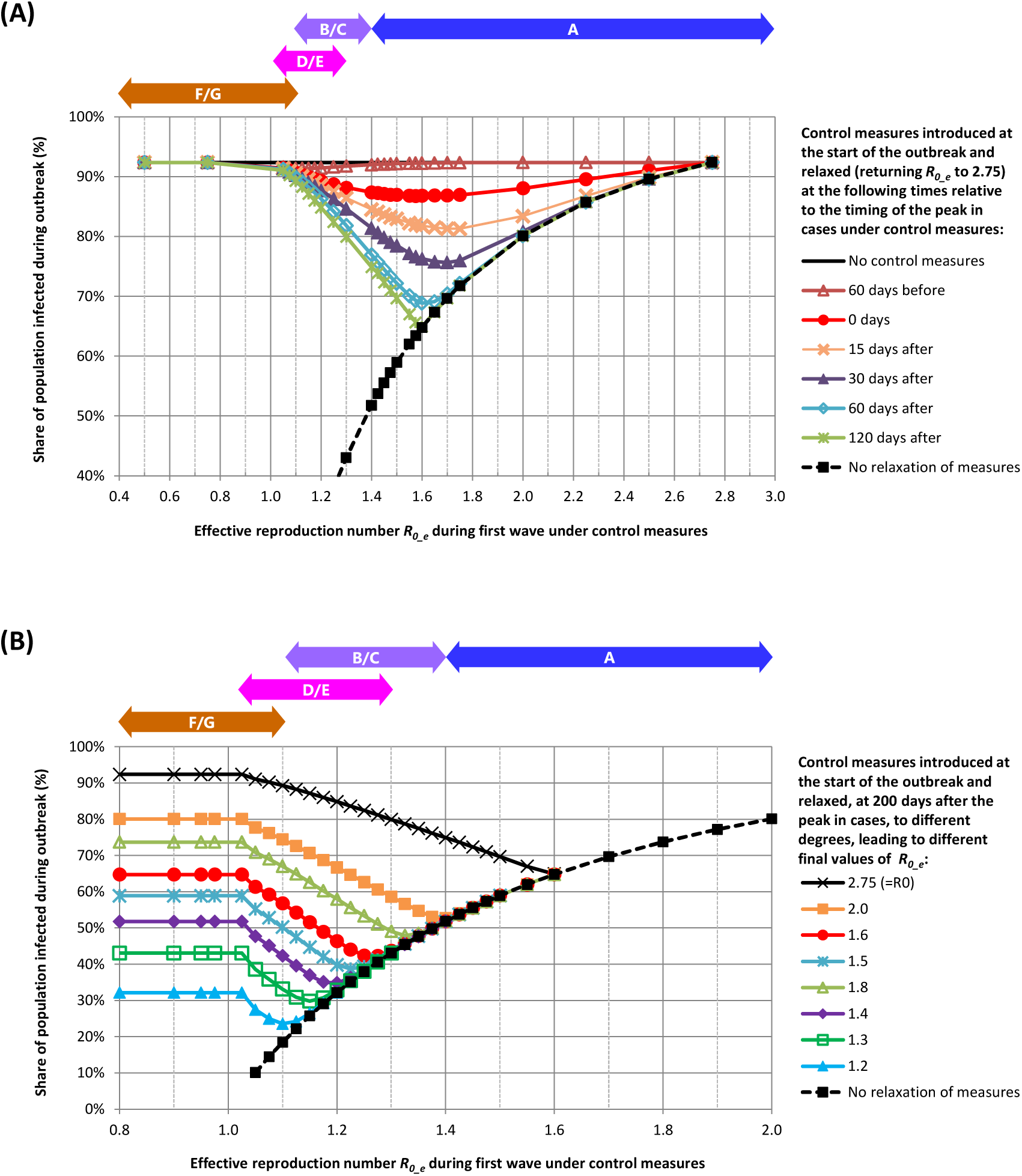
Expected total share of population infected during an epidemic for different levels of control measures and different timings and degrees of relaxation of control measures. Calculated values from a simple SIR model, with *R*_*0*_ = 2.75 and *t*_*g*_ = 5 days. (**A**) Control measures reduce *R*_*0_e*_ to between 0.5 and 2.75 at the start of the outbreak, and then are relaxed to allow *R*_*0_e*_ to return to *R*_*0*_, at different times after the peak in cases. (**B**) Control measures reduce *R*_*0_e*_ to between 0.8 and 2.0 at the start of the outbreak, and then are relaxed at 200 days after the peak in cases, to different values of *R*_*0_e*_ post-relaxation. Arrows indicate the outbreak dynamics categories (A, B/C, D/E, F/G) that roughly correspond to the initial values of *R*_*0_e*_ in the simulations – noting that this correspondence is only approximate as *R*_*0_e*_ varies over time for most countries.

As shown in both panels, for these simulations, if there is a complete first wave of the epidemic mitigated by measures, it is possible to relax measures to some degree without causing a second wave and additional infections – if the infection level at the time that measures are relaxed is less than the effective herd immunity threshold under the relaxed measures. If the first wave is complete, and if *R*_*0_e*_ *= R*_*1*_ during the first wave, the total infection level *Z*_*1*_ at the end of this wave is determined by the equation 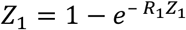 *[39]*. If *R*_*0_e*_ *= R*_*2*_ after measures are relaxed, the effective herd immunity threshold is 1 − 1/*R*_2_. Consequently, there will no second wave if

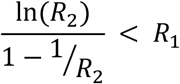

Figure 14 shows the maximum values of *R*_*2*_ for which there is no second wave after relaxation of measures, from this inequality. However, if measures are relaxed before the first wave is complete, the infection level will be lower than *Z*_*1*_, and the maximum value of *R*_*2*_ for which there is no second wave will be lower than that determined by the above inequality. If *R*_*2*_ *= R*_*0*_ satisfies the inequality, then it is possible to relax measures fully.

Virus variants with higher transmissibility increase *R_0_e_* even if control measures stay constant, and thus can generate second waves of sizes determined by figure 13. Virus variants that evade immune responses, or waning of immunity from infection, would allow for re-infections, and thus for greater numbers of additional infections after first waves than those suggested by figure 13.

**Figure 14:**
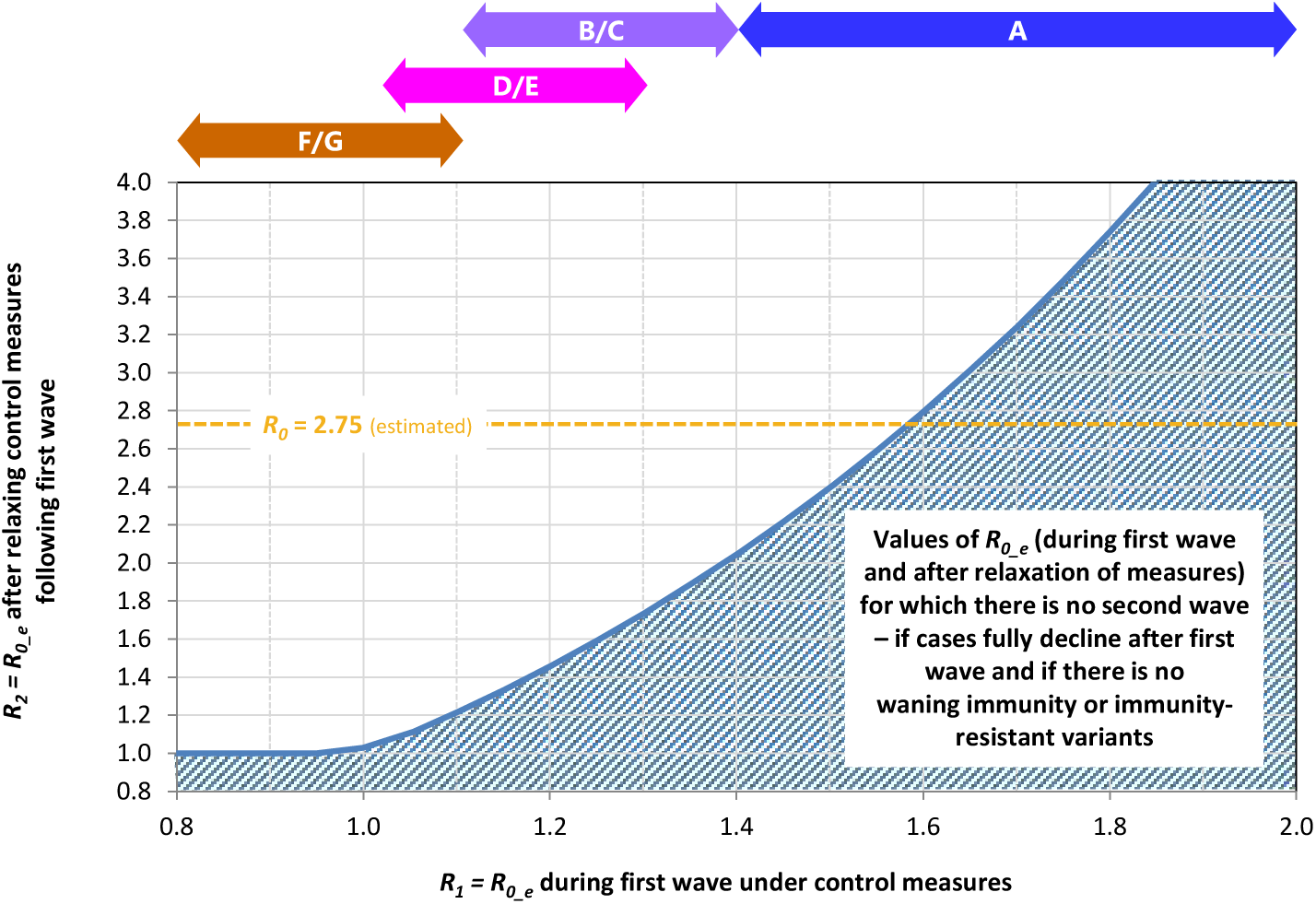
Maximum values of *R*_*0_e*_ after relaxation of measures, for different values of *R*_*0_e*_ during the first wave under control measures, for which there is no second wave after relaxation of measures. Values determined from the inequality presented in the text, which assumes relaxation of measures only after the first wave if fully complete, that there are no significant homogeneities in the population and that there is no waning of immunity or emergence of immunity-resistant variants. Arrows indicate the outbreak dynamics categories (A, B/C, D/E, F/G) that roughly correspond to the values of *R*_*0_e*_ during the first wave – noting that this correspondence is only approximate as *R*_*0_e*_ varies over time for most countries.

#### For countries in which the disease has declined mainly due to low susceptibility levels, and in which most people have been infected (Category A)

There may be room to relax control measures – subject to some important cautions – without putting additional lives at risk from the disease. Per figure 13, relaxing control measures is not expected to lead to many additional infections (unless relaxation occurs very soon after the peak in cases or values of *R*_*0_e*_ lie between 1.4 and 1.6). Most countries in Category A have already relaxed control measures after case numbers declined, and they may be able to relax measures further – re-opening schools, re-starting other health programmes, letting people go fully back to their livelihoods, permitting international travel. However, there are two important points of caution.

First, many Category A countries have upper- and middle-class population segments which are likely to have implemented greater degrees of social distancing during the epidemic to date, and hence likely to have much lower infection levels than in the overall population; these segments could experience substantial outbreaks if they relax their behaviours or if new higher-transmissibility variants emerge (which has occurred already in several regions). Health authorities should consider approaches to encourage people in such population segments to maintain disease control measures.

Second, waning immunity or new immunity-resistant strains of SARS-CoV-2 might lead to new outbreaks. If immunity loss is complete or new variants are fully able to evade immune responses, or if new variants have higher basic reproduction numbers, then it would be unlikely that control measures would be any more effective at constraining new outbreaks than they were in 2020. If immunity loss or resistance to immunity in new variants is only partial, and if the reproduction numbers are not much higher than in the original variant, then it may be possible to suppress new outbreaks, or at least to “flatten the curve” significantly, with control measures similar to those used in 2020 – and governments would need to determine whether the economic and social costs were justified given the potential to protect lives (and the risk to lives would depend on the infection fatality rates for reinfections after waning immunity or with new variants of the virus).

#### For countries in which control measures slowed the disease spread significantly but did not suppress it, after moderate numbers of people have been infected (Categories B and C)

For countries in Categories B and C, in which reported case numbers have fully declined from their peaks, it should be possible to relax those control measures that have the greatest negative health, economic and social consequences. As indicated in panel B of figure 13, for Category B and C countries, the value of *R*_*0_e*_ can increase somewhat without generating additional cases. However, full returns to normal (until enough people have been vaccinated) should be avoided, because large new outbreaks will happen if *R*_*0_e*_ is decreased by too much – as appears to have happened in Kenya and Bolivia.

Countries in Categories B and C, like those in Category A, need to be careful about of risks of outbreaks in population segments that may have maintained lower infection levels during the outbreak to date.

If, and most likely when, they experience waning immunity or immunity-resistant strains, the calculus for Category B and C countries is likely to be different from that for Category A countries. Having lower current infection levels, these countries face greater potential for loss of life if new outbreaks spread through the whole population. Even if immunity loss or resistance to immunity in new variants is total, and certainly if they are less than total, Category B and C countries may well be able to suppress new outbreaks with control measures similar in nature to those which resulted in a flattened curve during the first wave of the disease. However, higher-transmissibility strains pose a risk of large outbreaks, with or without the added danger of immunity resistance.

#### For countries in which control measures slowed the disease spread significantly but did not suppress it, and in which case numbers are still declining from the peak or are still increasing (Categories D and E)

For countries in Category D, in which reported case numbers have not fully declined from their peaks, control measures should not be relaxed until only after cases have fully declined. Even though some countries in Category D may have similar values of *R*_*0_e*_ under control measures as those in Category B, panel A of figure 13 shows that relaxing control measures too soon after a peak in cases can result in significantly greater numbers of infected people – as appears to have happened in Brazil, Colombia and Paraguay, for example, in which control measures were relaxed soon after case numbers peaked, and case numbers have since increased rather than continuing to decline.

For countries in Category E, where cases have been rising especially slowly, it may be possible to push *R*_*0_e*_ below 1 and hence “suppress the curve” by introducing some additional control measures or improving compliance with existing measures, or by starting vaccinations with people who are playing the greatest roles in spreading the disease.

Countries in Categories D and E, like those in Categories A-C, need to be careful about risks of outbreaks in population segments that may have maintained lower infection levels during the outbreak to date.

If, and most likely when, they experience higher-transmissibility or immunity-resistant strains, or waning of immunity from first infections, Category D countries could experience resurgences in cases – which may have happened in Argentina and Brazil – and Category E countries could experience accelerations in the growth of case numbers – as appears to have happened in Mozambique and Togo. Additional control measures would be required to keep case numbers declining or to keep the case curve as flat as it has been to date.

#### For countries in which disease control measures have suppressed the spread of SARS-CoV-2 (Categories F and G) or largely kept it out (Category H)

COVID-19 control measures, put in place by governments and implemented by citizens, have saved perhaps 13.1–14.2 million lives in Category F-H countries. To continue to protect these lives, control measures need to be maintained until vaccines become widely available. Measures will need to be strengthened to compensate when higher-transmissibility variants emerge – as has already taken place in Europe in recent weeks as the B.1.1.7 variant spread. Note that immunity-resistant strains do not necessarily add to the risk in countries in Categories F, G and H – except to the extent that such strains may reduce the effectiveness of vaccines and require the development and distribution of new vaccines.

### Vaccination strategies

Multiple vaccines have been developed, and are being deployed, which have high levels of effectiveness in preventing COVID-19 *[40]*. The effectiveness of these vaccines may be reduced for some new variants of the SARS-CoV-2 virus. It is not yet clear to what extent the vaccines reduce transmission of the virus, either of the early variants or the more recent variants with higher transmissibility and/or ability to evade immune responses.

Announcements made by various companies suggest that perhaps 12.5 billion doses of the major vaccines could be produced in 2021, enough for 6.25 billion people (as the vaccines require two doses to be effective), or most of the world’s population. However, supplies are limited at present and will continue to be limited globally at least until the middle of 2021. And supplies will be limited in most of the world well beyond that, with most countries only expected to achieve widespread vaccination coverage by 2022 or 2023, according to the Economist Intelligence Unit *[41]*. The limitations on supply of vaccines raises several questions. First, to which segments of the population should vaccines be deployed first? Second, how should vaccines be allocated across countries? Answers to these questions should be driven principally by ethics and equity, among countries and individuals. Making these choices requires information on the potential benefits, in terms of deaths averted or life-years saved, of different choices. Key analytical questions include: With a limited supply of vaccine, is it better to vaccinate directly those who are most at risk of death or severe illness, or is it better to vaccinate the general population, or key transmitters within the population, to constrain disease spread and hence protect (indirectly) greater numbers of vulnerable people? What allocation of vaccine doses across countries will protect the greatest numbers of people – taking into account both different numbers of vulnerable people and the potential benefits of possible vaccination strategies in different countries?

A.B. Hogan et al. have used mathematical models to study some of these questions and reach several important findings *[42]*. The number of cases and deaths averted through vaccination is, not surprisingly, greater if the proportion of people already infected is smaller at the time vaccination starts and/or if the immunity conferred by past infection has a shorter duration. More deaths per thousand people are averted in HICs and UMICs than in LMICs and LICs, because HICs and UMICs have older populations, if the ability of health systems to save lives of COVID-19 patients is ignored, but the numbers are roughly similar across country income groups if the constraints on health systems are factored into the estimates – and the numbers of life-years gained is higher in LICs and LMICs. They find that, “within a country, for a limited supply (doses for <20% of the population) the optimal strategy is to target the elderly and other high-risk groups” but that, if a larger supply is available within a short time, “the optimal strategy switches to targeting key transmitters (i.e., the working age population and potentially children) to indirectly protect the elderly and vulnerable” – for simulations which assume that small numbers of people have previously been infected.

Current policies in several countries call for deployment of vaccines first to healthcare workers and then by age cohort, starting with the oldest *[43]*.

For countries in Category A, our findings imply that early vaccination should concentrate on elderly and vulnerable people. There is no alternative strategy to consider in the short term, because vaccination is not required to create herd immunity in the population to the initial variants of the virus – although vaccination to create herd immunity could become relevant for future variants if they evade immune responses and if vaccines are available to protect against such new variants. The greatest danger, both now and into the future, lies in the threat to elderly and vulnerable people from endemic SARS-CoV-2 over time, as the disease is unlikely to be eliminated completely.

For countries in Categories F and G and in Category H, which have low current infection levels, the simulations of Hogan et al. are most applicable, and vaccinations should be given first to frontline healthcare and other essential workers and to elderly and vulnerable groups (starting with the oldest and most vulnerable).

For countries in Categories D and E, there may be situations in which the optimal strategy for early vaccination switches to targeting key transmitters (mainly working-age people) and indirectly protecting the elderly and vulnerable, even if the supply of vaccine doses is enough for only a small share of the population. In some of these countries, if key transmitters are vaccinated (with vaccines that are capable of limiting transmission and not just reducing severity of disease and mortality), and if current disease control measures are maintained and higher-transmissibility virus variants don’t become common, then it may be possible to halt transmission of the disease completely, while waiting for further vaccine supplies to arrive (after which disease control measures could be released). The strategy to start with key transmitters to suppress the disease has already been adopted by Indonesia *[44]*, and a recent paper proposed that mass vaccination of adults aged 20-49 should be considered in the United States to bring the disease under control because the reproduction number for this group has remained above 1 and sustained resurgences in cases *[45]*. For each country in Categories D and E, careful modelling would be necessary to determine if the key transmitter strategy is optimal, taking into account the likely supplies of vaccines and when they will be available. Vaccination of elderly and vulnerable groups would likely be the most prudent choice in the absence of good country-specific modelling and an effective plan for identifying and vaccinating key transmitters. In countries, such as Indonesia, that opt for a strategy to vaccinate working-age people first, it will be important to keep control measures in place and to keep high-transmissibility variants of the virus out; otherwise, the benefits of the key transmitter vaccination strategy will be lost, and the COVID-19 cases will continue to grow in number.

For countries in Category B, vaccinations should probably be given first to frontline healthcare and other essential workers and to elderly and vulnerable groups (starting with the oldest and most vulnerable), similar to countries in Categories F, G and H. However, if there are resurgences in cases across the population (and not just in limited segments of the population), due to higher-transmissibility or immunity-resistant variants or otherwise, then the optimal strategy may switch to targeting key transmitters, similar to some countries in Categories D and E.

In allocating vaccine doses across countries, Hogan et al. found that, “the strategy that maximises the total deaths averted is one in which the available vaccine doses are allocated preferentially to higher-income countries who have the highest at-risk elderly populations,” but noted that “the optimality of this allocation is sensitive to many assumptions and will vary depending both on the vaccine characteristics and the stage of the epidemic in each country at vaccine introduction,” and concluded that, “[g]iven this uncertainty, allocating vaccine doses according to population size appears to be the next most efficient approach.”

Our findings reinforce the uncertainty strongly: it is very likely that that stage of the epidemic varies greatly across countries. For most countries, the optimal allocation of vaccines doses is likely still to be according to population size (or according to population over 60, scaled up by factors of perhaps 3-4 for LICs and LMICs with less effective healthcare) – and then for those countries to give doses first to elderly and vulnerable people. However, the global optimal allocation strategy might include providing somewhat greater supplies, during the next few months, to Category D and E countries where using the vaccine to halt spread of the disease might be possible (provided that disease control measures are maintained in those countries). It is clear, in any case, that further modelling of vaccine allocation strategies is essential, taking into account the actual vaccine efficacies (in limiting both illness and virus transmission) and projected available doses by month, as well as allowing for disease stage categories in different countries.

Higher-transmissibility variants of the SARS-CoV-2 virus will not affect early vaccination strategies for most countries – other than to increase the urgency of distributing vaccines, especially in some Category F and G countries which may not be able to keep the disease suppressed once the higher-transmissibility variant begins to dominate new infections. The exceptions are any Category D or E countries that consider starting with vaccinations of key transmitters to suppress the disease spread; such countries will need to be sure that they can keep the disease suppressed even if higher-transmissibility variants become common.

Immunity-resistant variants of the virus may reduce the effectiveness of current vaccines. However, the immune responses generated by current vaccines are likely to be at least partially effective against new variants, even if the variant can avoid being fully neutralized by antibody responses.

In the longer term, vaccination strategies may need to plan for multiple cycles of vaccinations. Immunity-resistant virus variants will necessitate development and deployment of new vaccines that are fully effective against the new variants. Immunity acquired through vaccination is likely to wane over time – like immunity acquired through infection. In making long-term plans, countries will face a wide range of options for who to vaccinate (elderly and vulnerable populations, key transmitters or entire populations) and for frequency of vaccination (every 6 months, annual, or once if residual benefits are sufficient). Optimal strategies for each country will be complicated to determine, as the choice will depend on many factors, including vaccine effectiveness in reducing mortality and in reducing transmission, how effectiveness wanes over time, mortality rates and transmissibility of new variants (in general and in previously infected or vaccinated people), and, once the risks to life and health from endemic COVID decrease to the point where COVID-19 is not an overriding issue, comparison with other health and budgetary priorities.

## Data Availability

All data used for this research presented in this report can be freely downloaded from the cited sources. Much of the relevant data used is presented in Annexes A and B.

https://covid19.who.int

https://ourworldindata.org/coronavirus-data-explorer

https://www.bsg.ox.ac.uk/covidtracker

https://www.economist.com/graphic-detail/2020/07/15/tracking-covid-19-excess-deaths-across-countries

https://doi.org/10.1101/2020.07.23.20160895

## Authors, Acknowledgements and Declarations

### Authors

J. Paul Callan is a strategy consultant in the field of global economic and social development. He works on strategic planning, designing new initiatives and organizational development, for multilateral agencies, development banks, governments, foundations and NGOs. Paul holds a BA in theoretical physics from Trinity College Dublin and a PhD in physics from Harvard University.

*Contact.*

Carlijn J.A. Nouwen is a strategy consultant who works on economic and social development in sub-Sahara Africa. Her project portfolio focuses on healthcare, sustainable economic development, and investment strategies. Carlijn holds a MSc in industrial engineering and management science from Eindhoven University of Technology.

*Contact.*

Axel S. Lexmond is an academic researcher in the field of computational modelling of multi- phase fluid dynamics. He has worked at universities in the Netherlands and South Africa and for applied research companies. Axel holds a MSc in chemical engineering from Delft University of Technology and a PhD in engineering from Eindhoven University of Technology.

Othmane Fourtassi is an analyst on global economic and social development, who works with clients in the social and public sectors. He holds a BA in economics from Yale University.

### Author contributions

J.P.C. conceived the study and devised the categorization scheme and approach. J.P.C. and C.J.A.N. conducted the categorization of countries and territories based on the defining characteristics for each category. O.F. and J.P.C. created the Excel model used to implement the approach, analyze data and make estimates. A.S.L. and C.J.A.N. developed the SIR model to simulate different scenarios for disease control measures, and J.P.C. helped to define the scenarios. J.P.C. prepared the manuscript, following discussions with and incorporating input from all authors.

## Acknowledgements

The authors wish to thank Muhannad Alramlawi, Yuliya Meskela, Debora Mulokozi and Christelle Nayandi for help with parts of the research. We also acknowledge many useful discussions including with Nynke Dekker, Turlough Downes, Edwin Macharia, James Mwangi, Tobias Rinke de Wit and Raymond Russell. However, the authors are solely responsible for the content of this report.

## Annex A: Evolution of Reported Cases and Stringency Index for Each Country and Territory

In this Annex, we present the data on reported daily new cases, as recorded in the WHO Coronavirus Disease (COVID-19) Dashboard, and on the the “stringency index” of control measures compiled by the Oxford COVID-19 Government Response Tracker. The data was downloaded on 2 February 2021.

The reported new cases are averaged over 7 days for most countries and over 14 days for a few countries in which case reporting was often less frequent than once a week.

The stringency index is shown on an inverted scale. This makes comparison easier with the figures showing model disease dynamics in response to changes in the effective basic reproduction number (in figures 1 and 9). However, although increases in stringency index are expected to decrease *R*_*0_e*_, changes in stringency index will not necessarily produce proportional changes in *R*_*0_e*_ (as some measures are likely to more impact on *R*_*0_e*_ than others), and how much change in stringency index is required to produce a given change in *R*_*0_e*_ is likely to vary strongly across countries.

Countries and territories are grouped into the main COVID-19 status categories determined through our analysis, and into further sub-categories as described in the headings below.

At the start of each category, a box provides a short description of the disease dynamics characteristic of the category and a typical example of the expected evolution of actual cases the category. The typical examples are identical to the corresponding panels in figure 1, except that the scales on the axes are adjusted: the timeframe (on the horizontal axis) is limited to end on 31 January 2021 (like the country data presented) and the maximum value on the actual daily new cases (vertical) axis is adjusted for some categories.

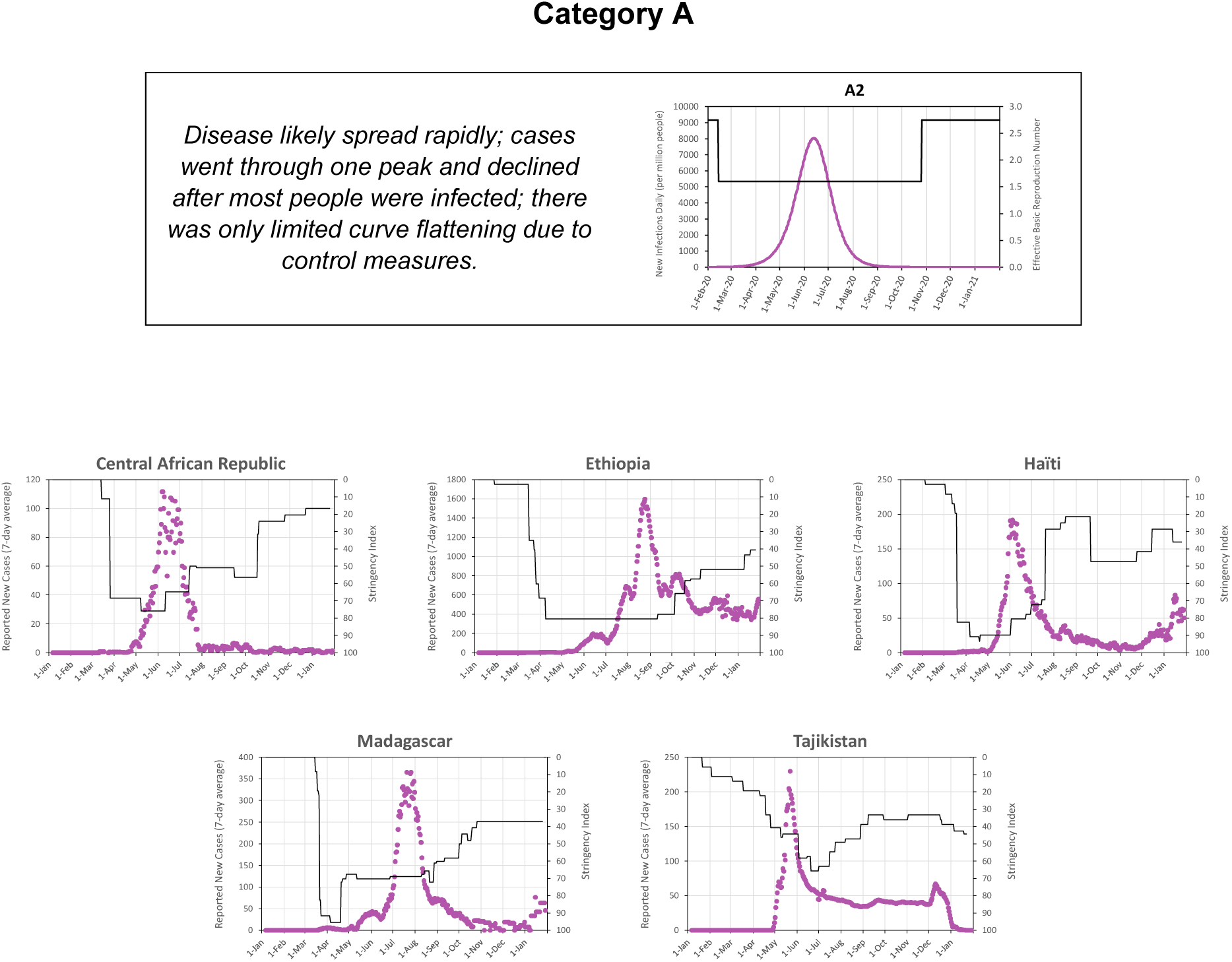

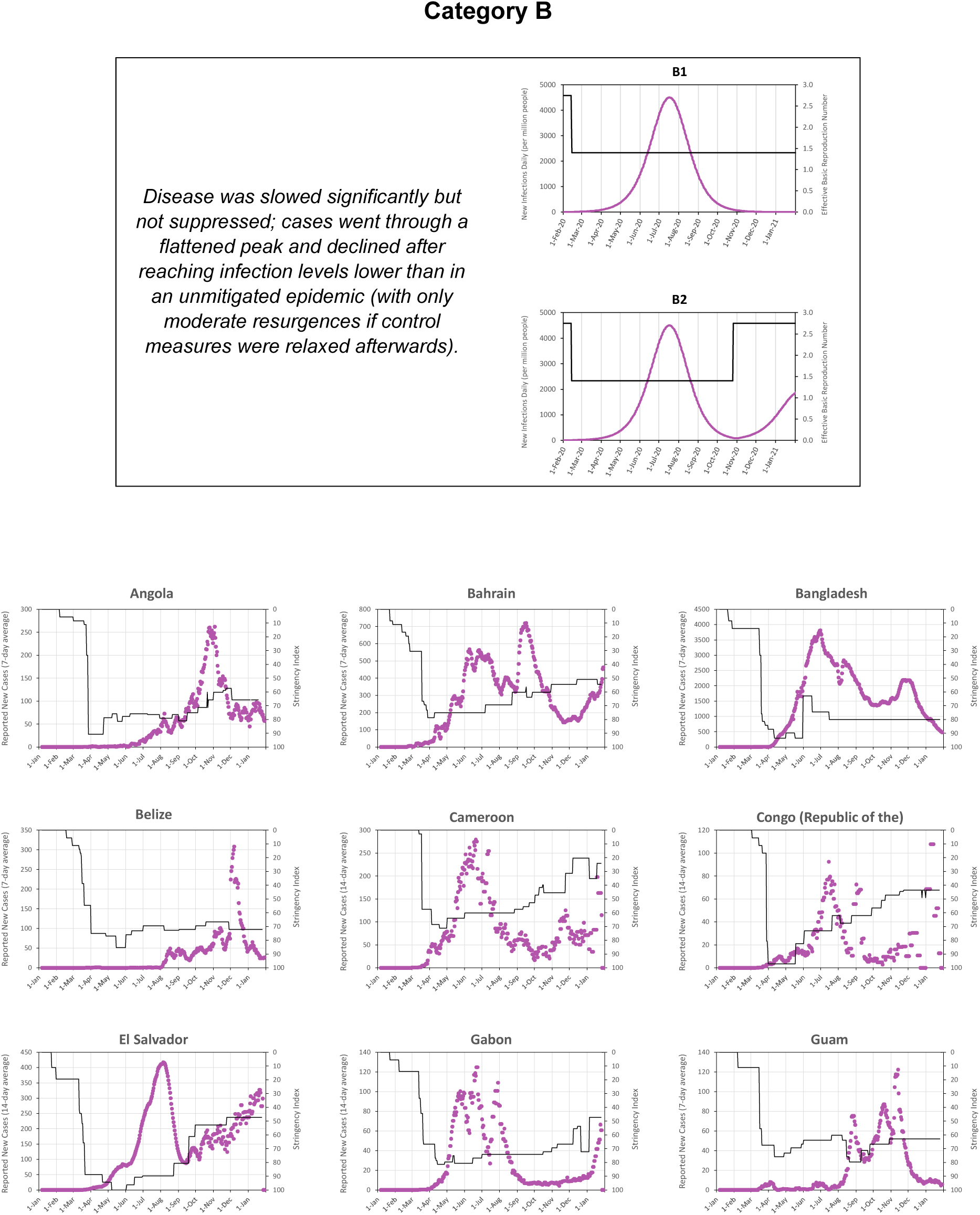

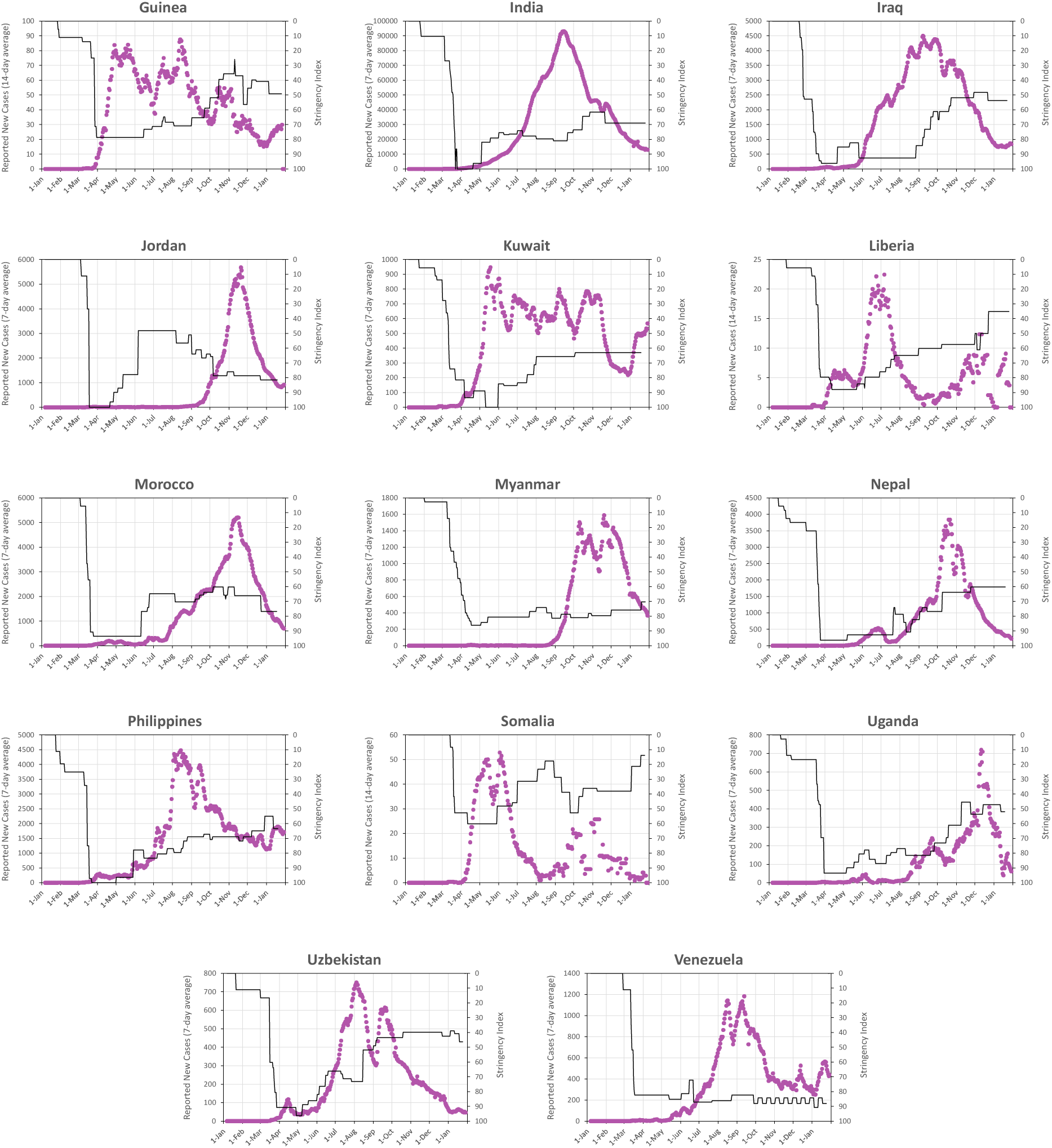

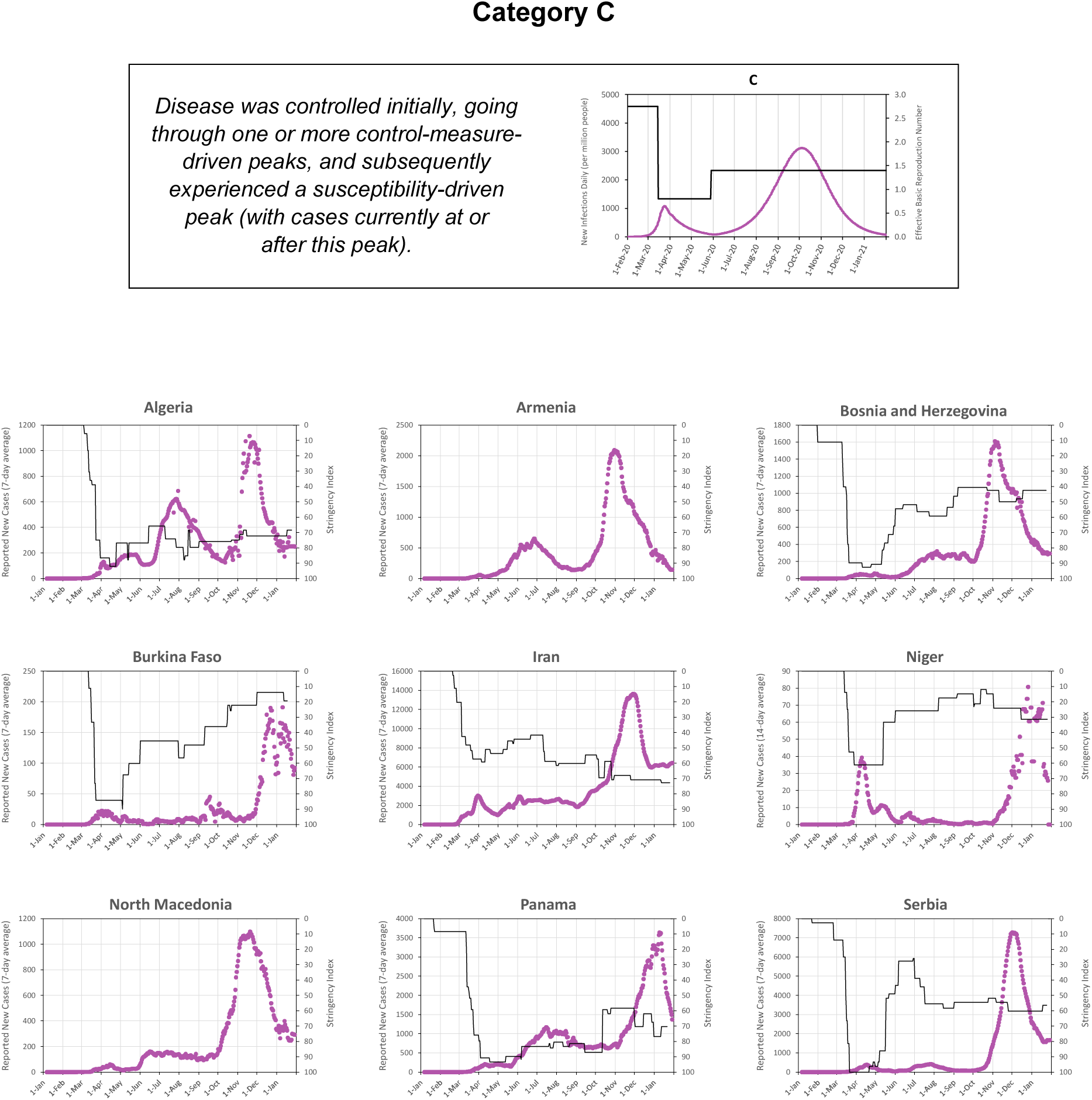

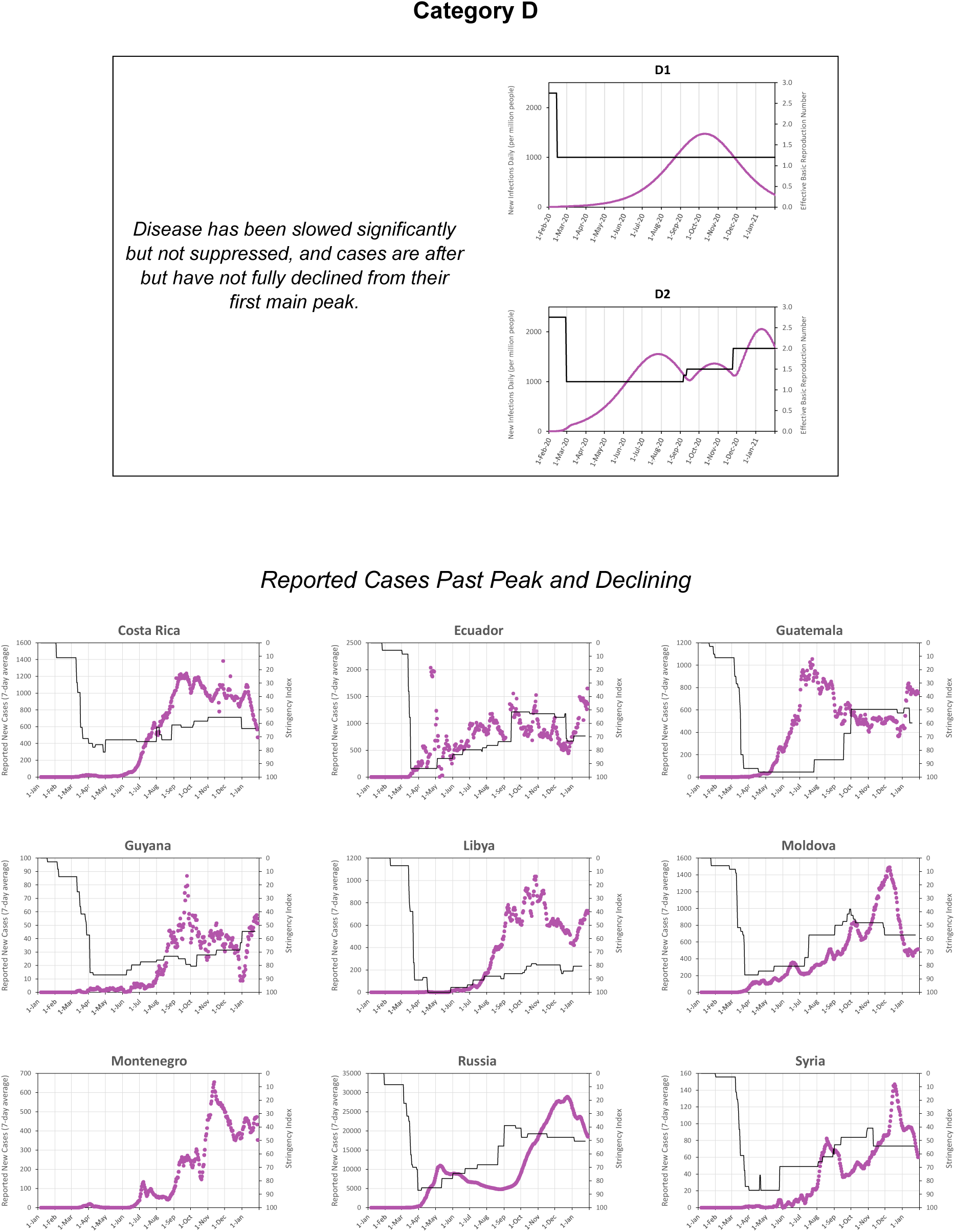

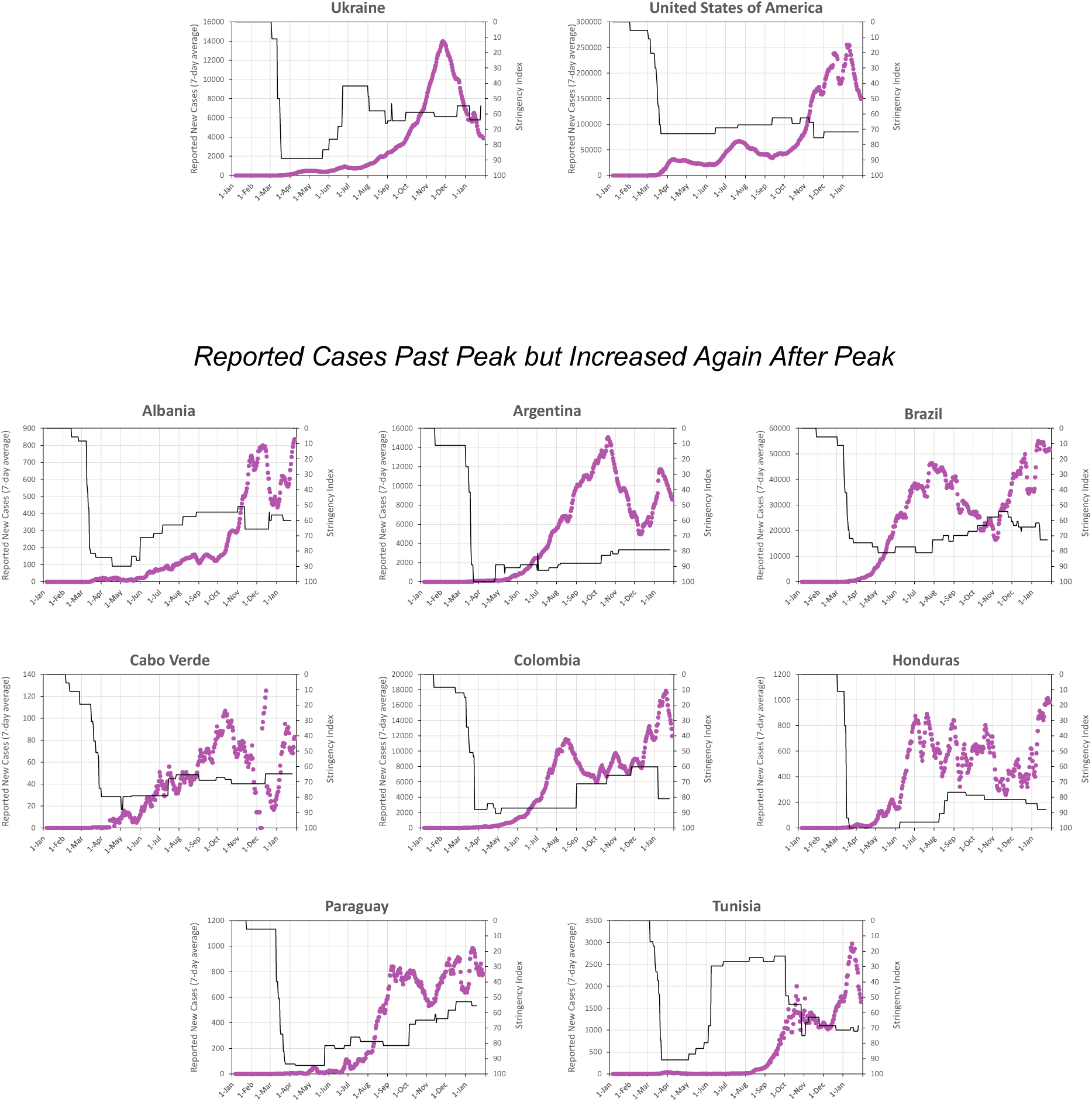

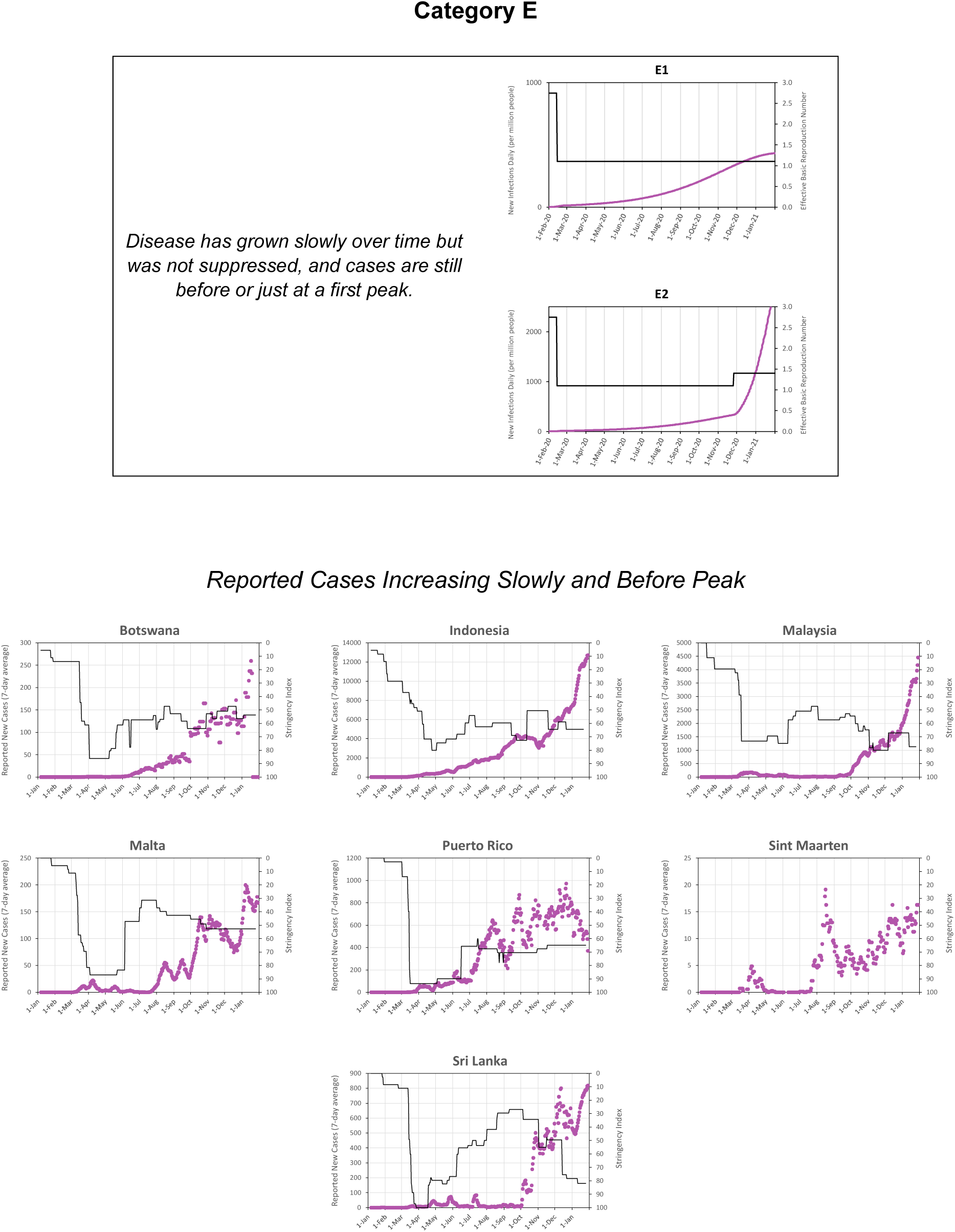

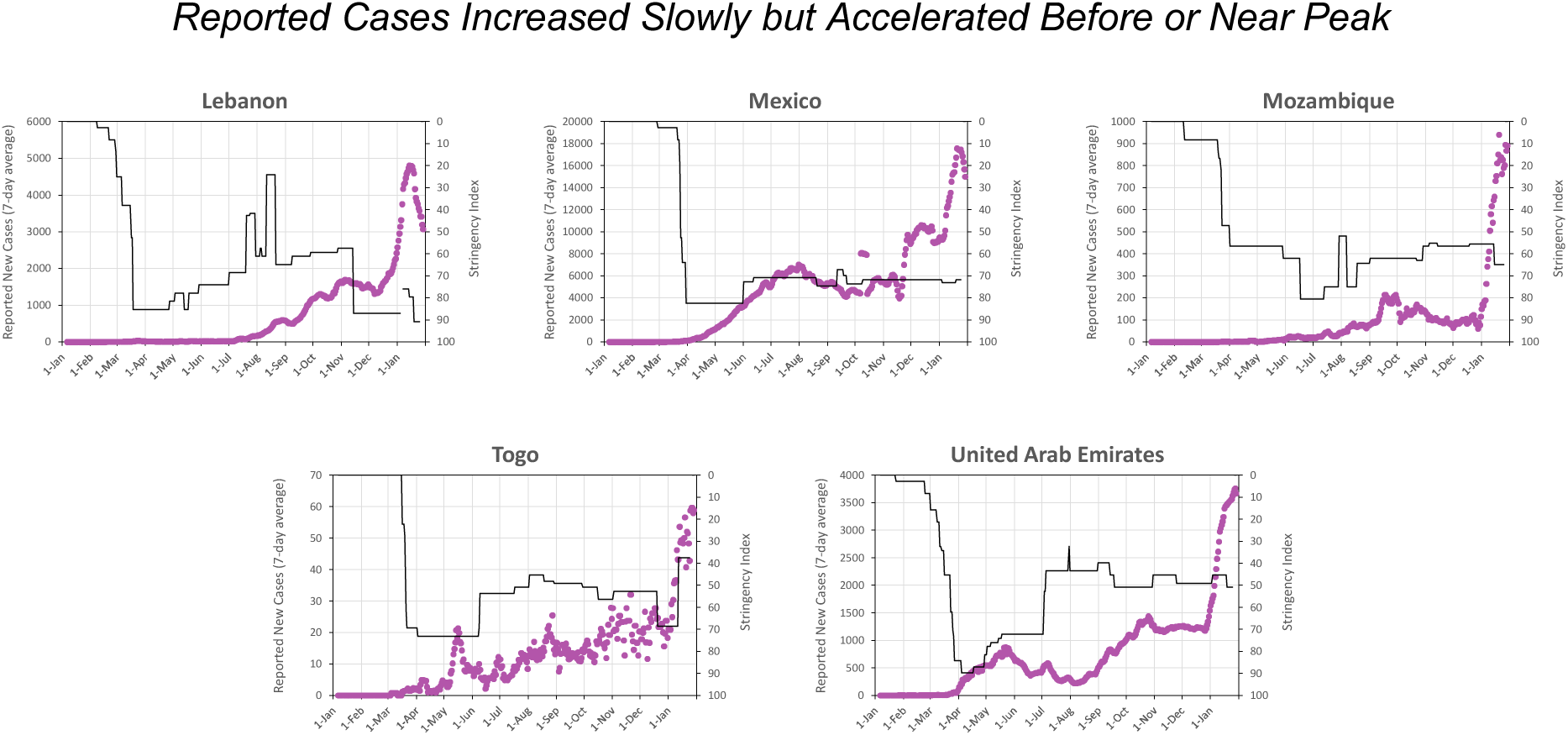

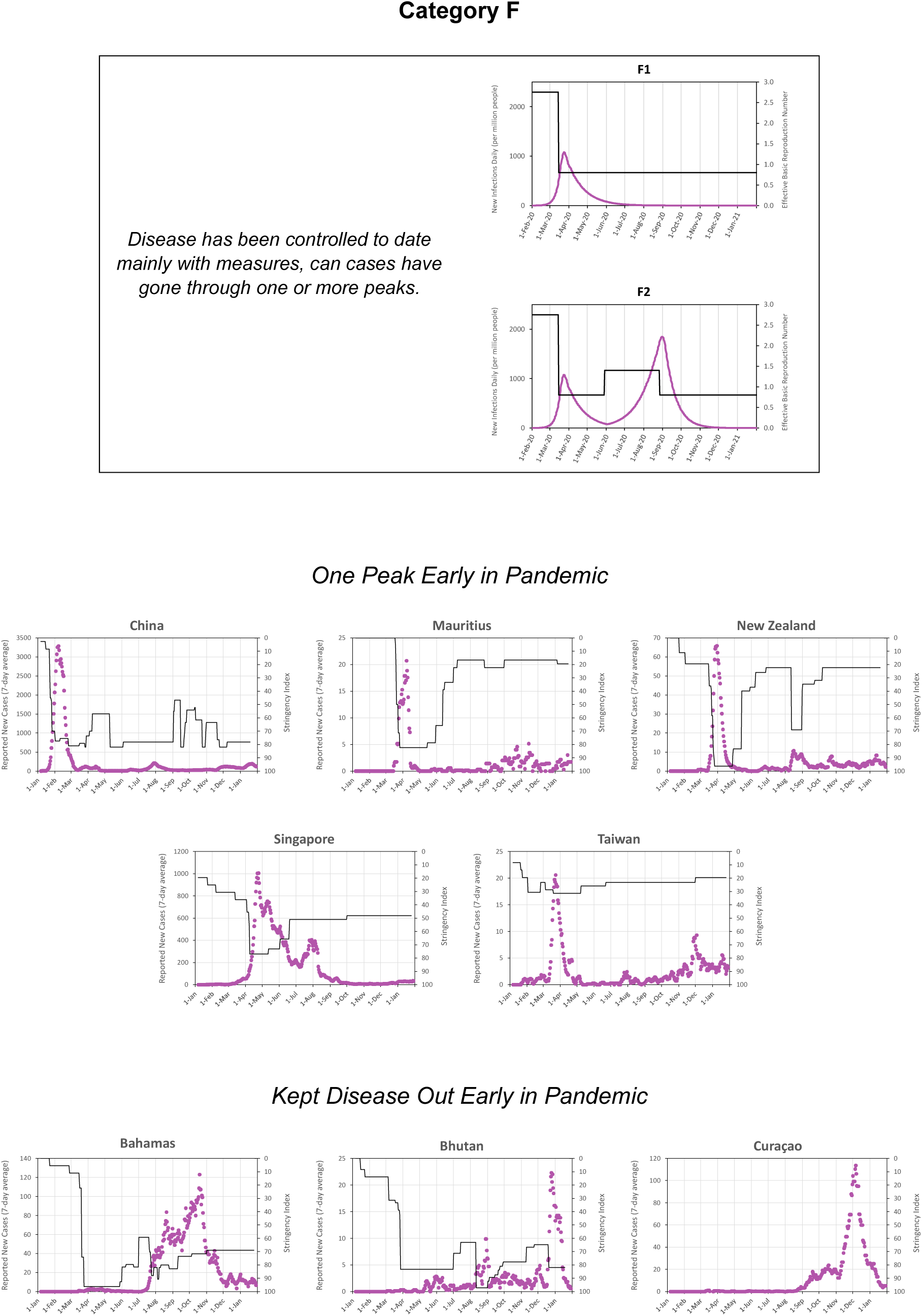

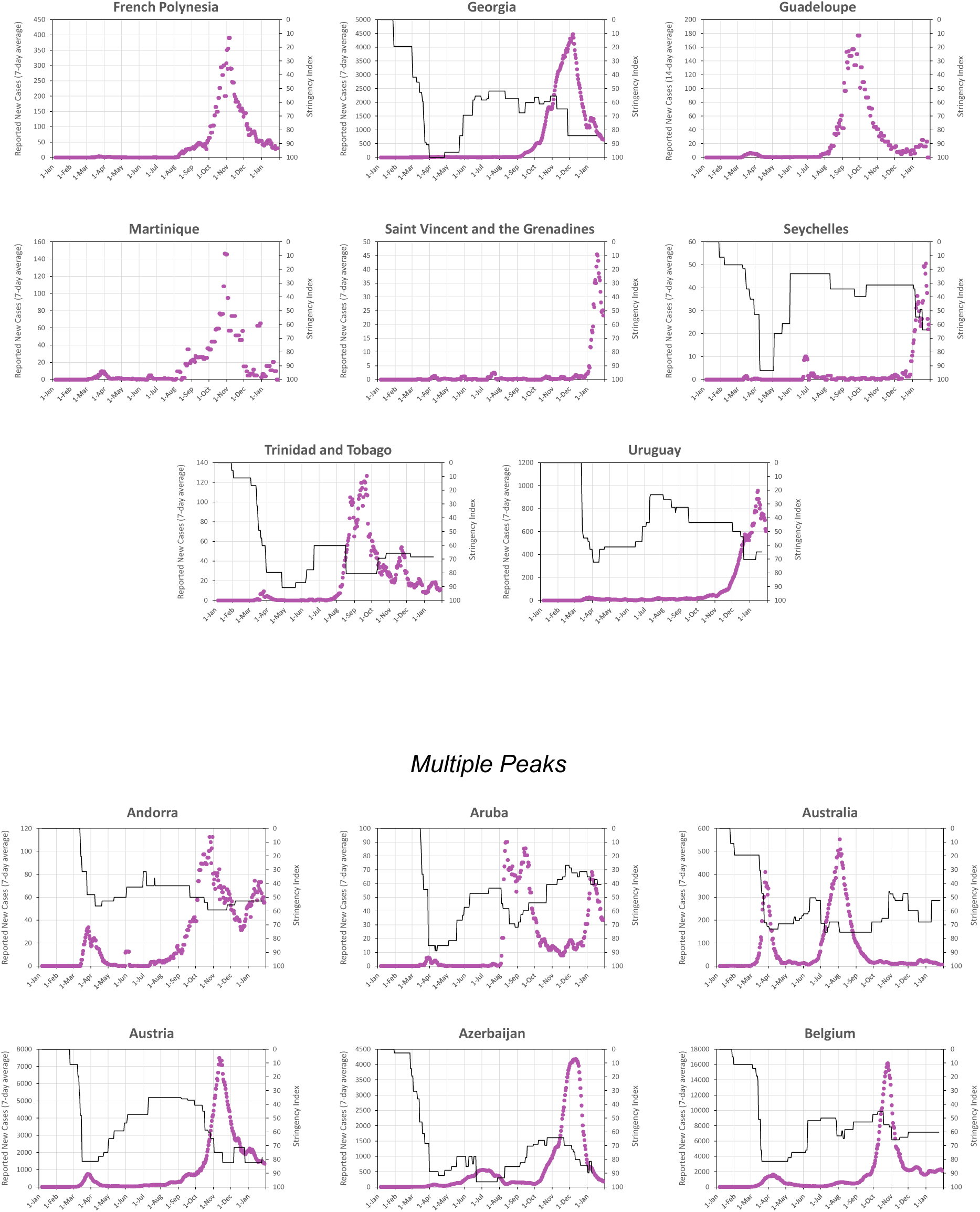

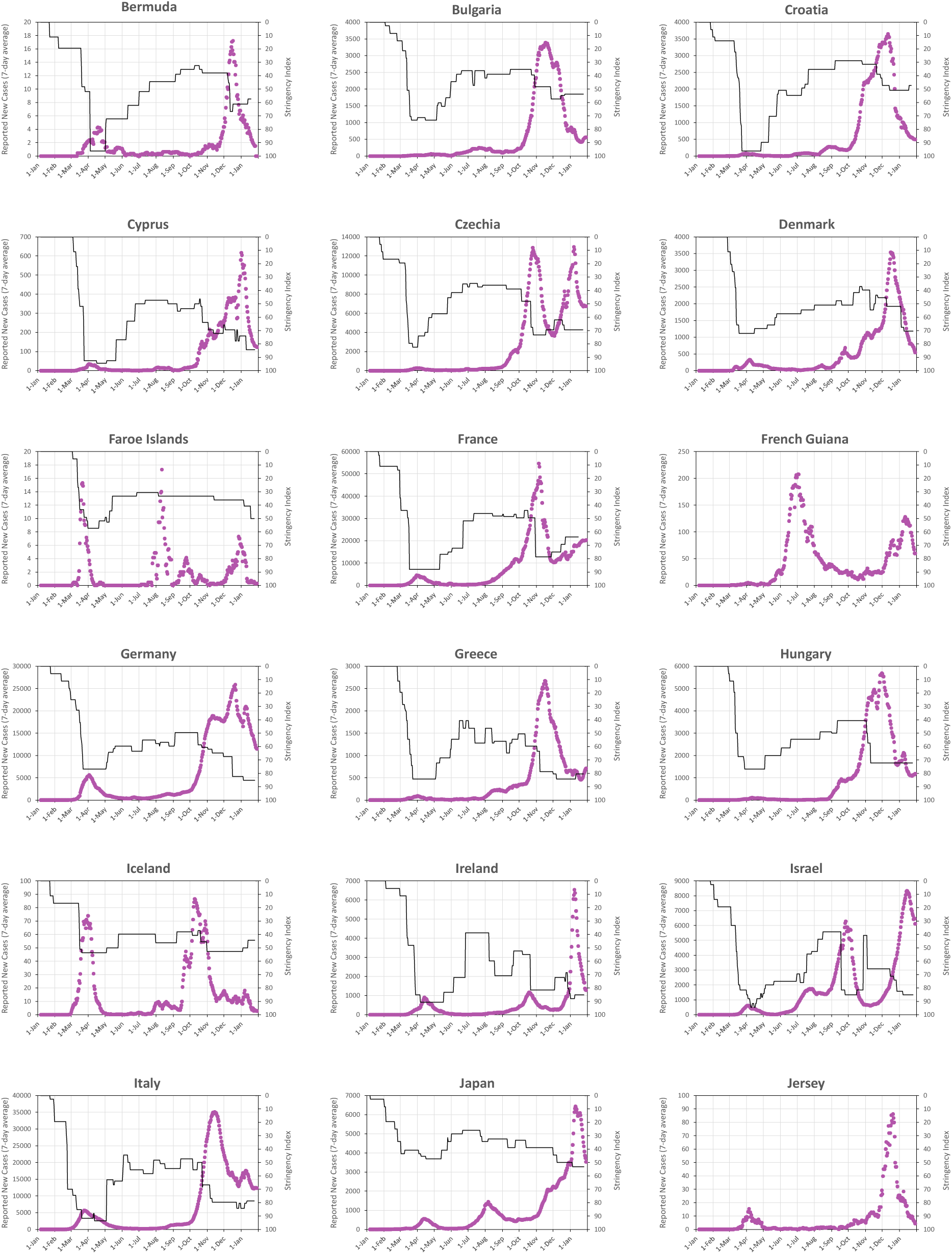

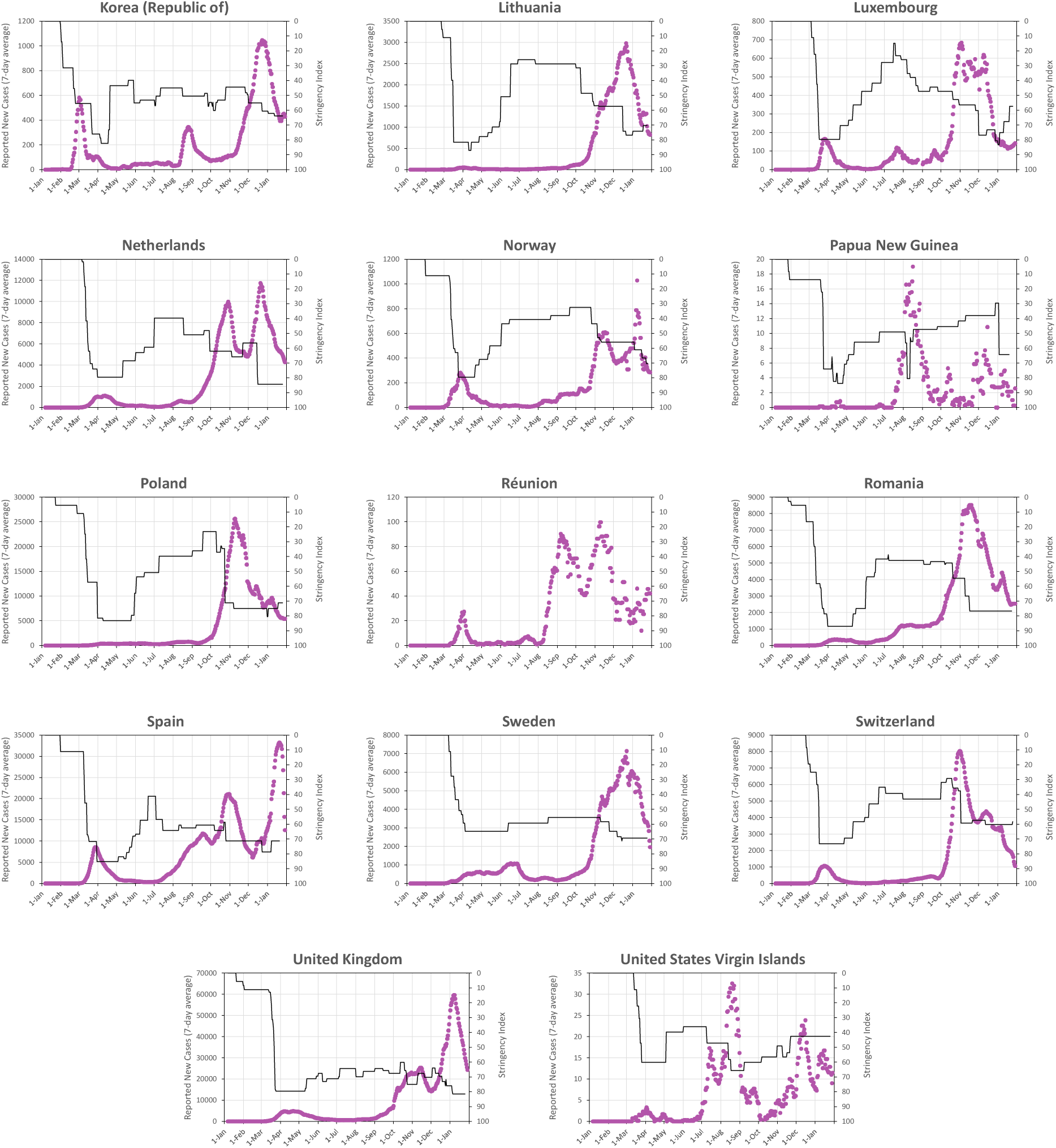

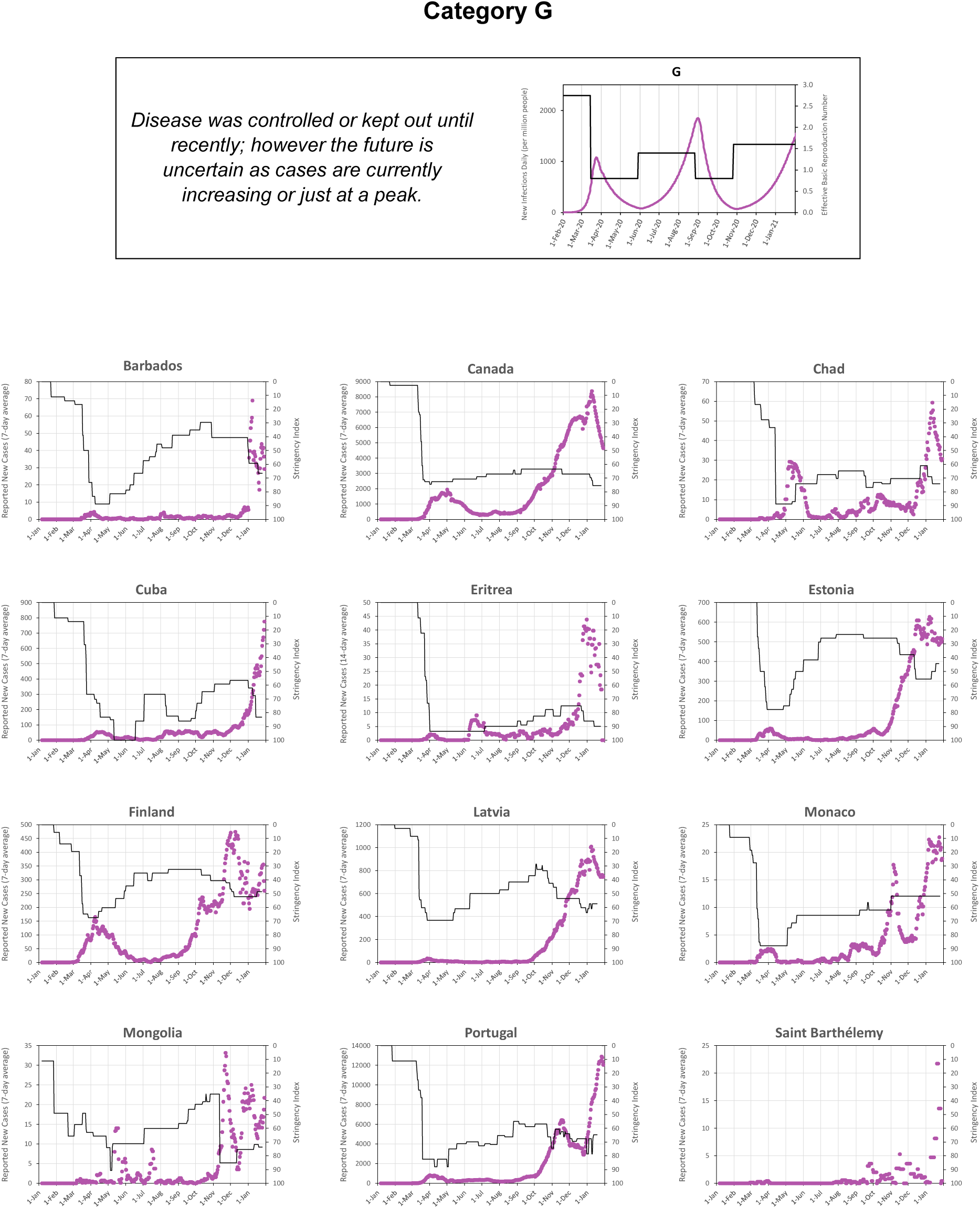

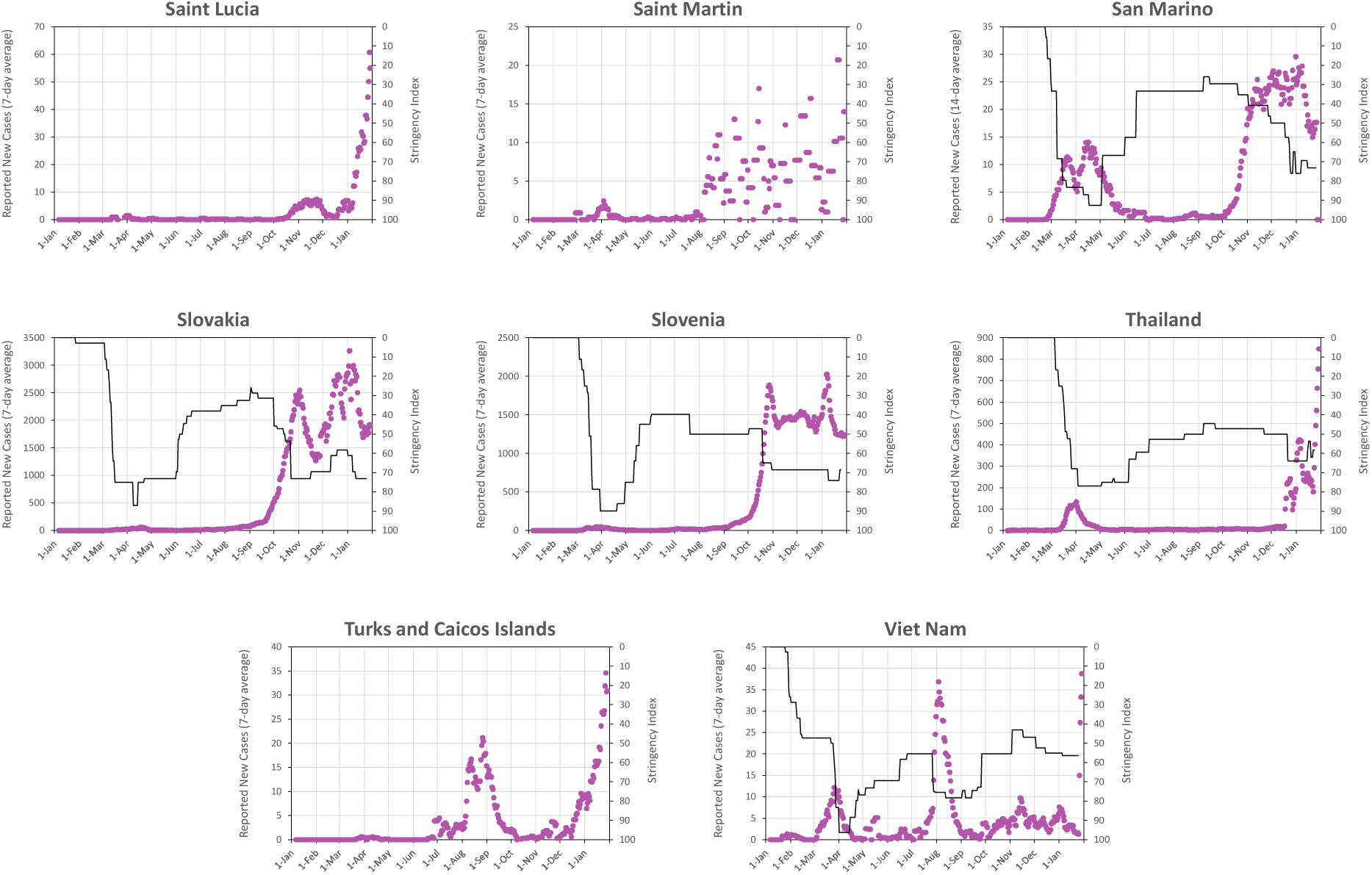

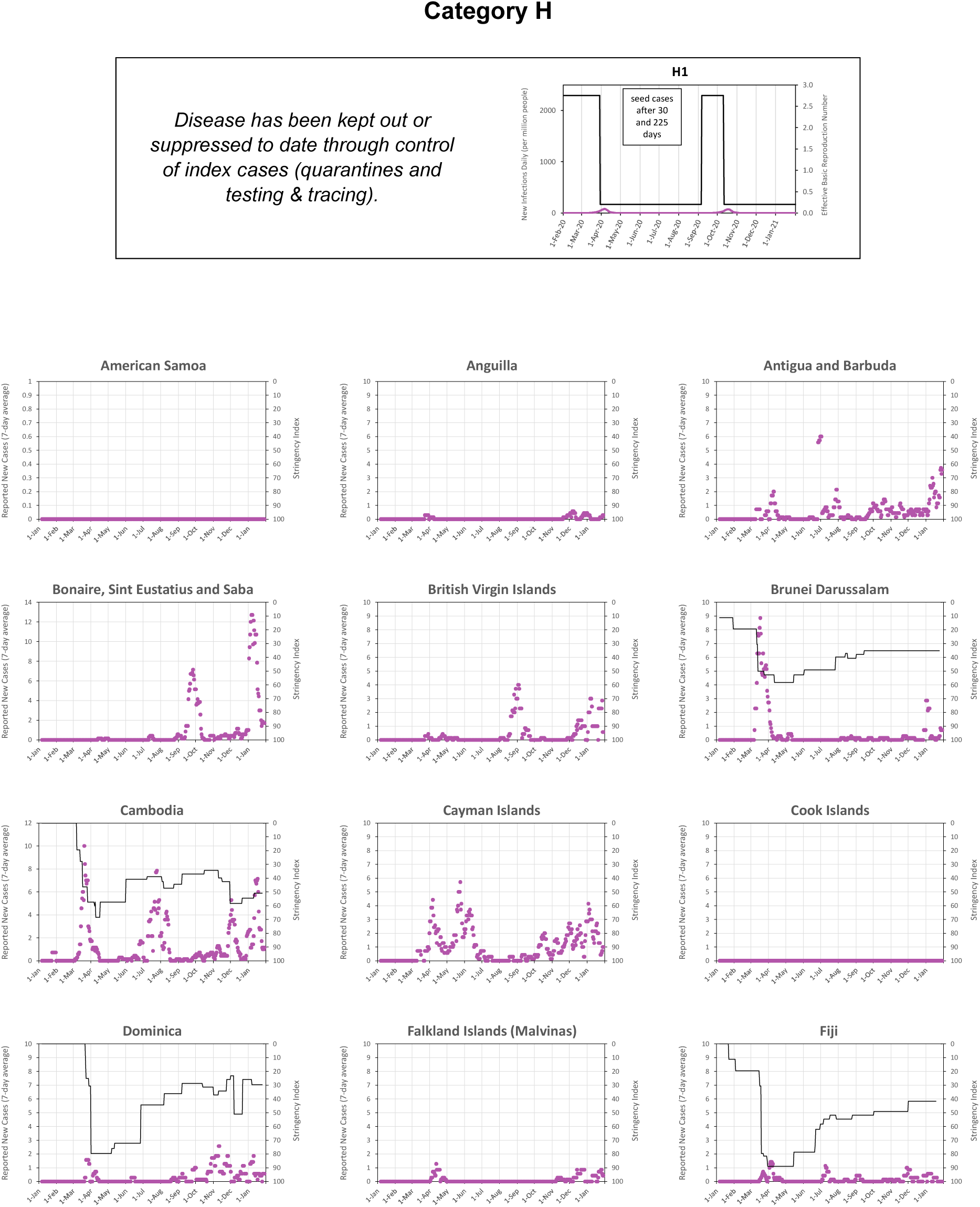

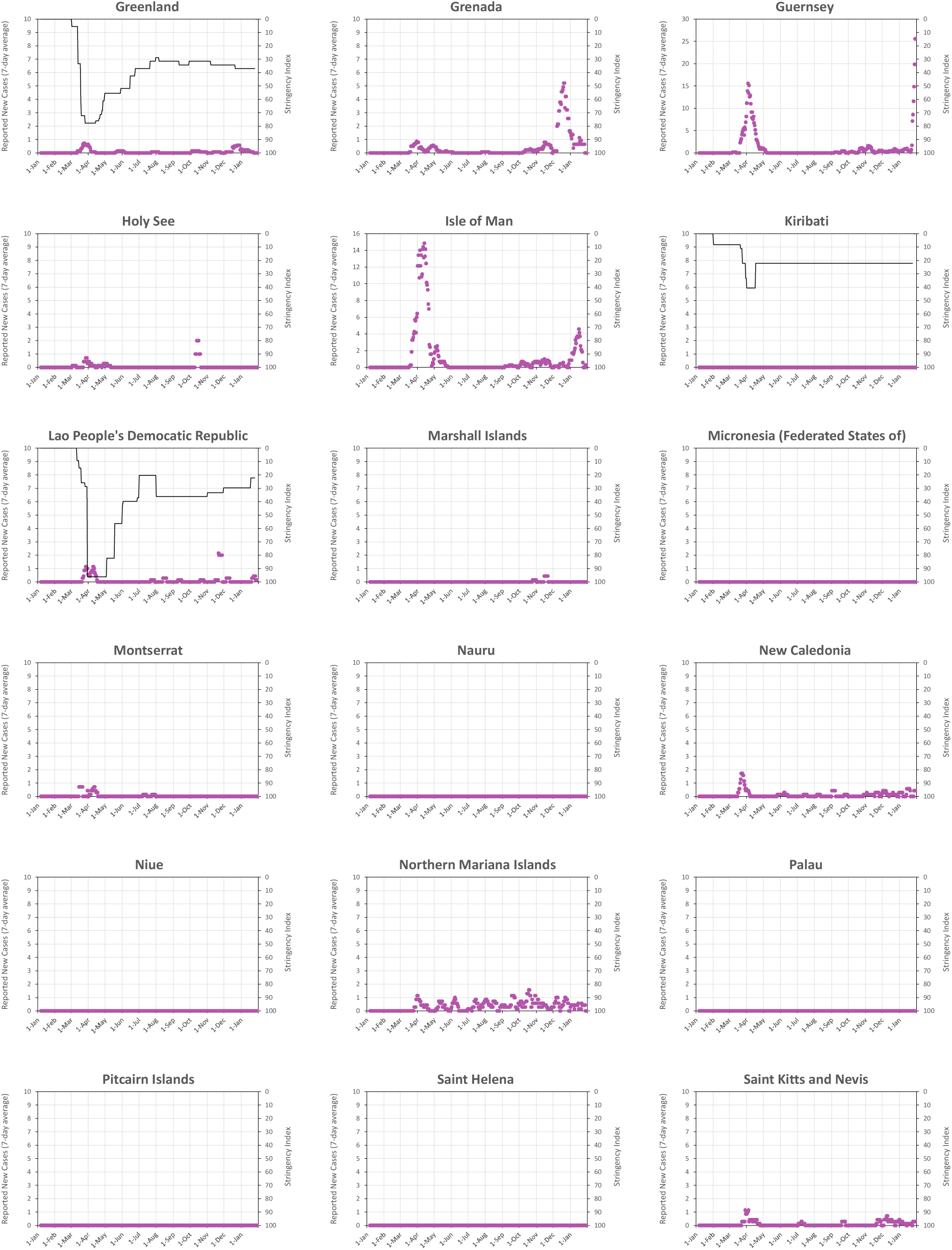

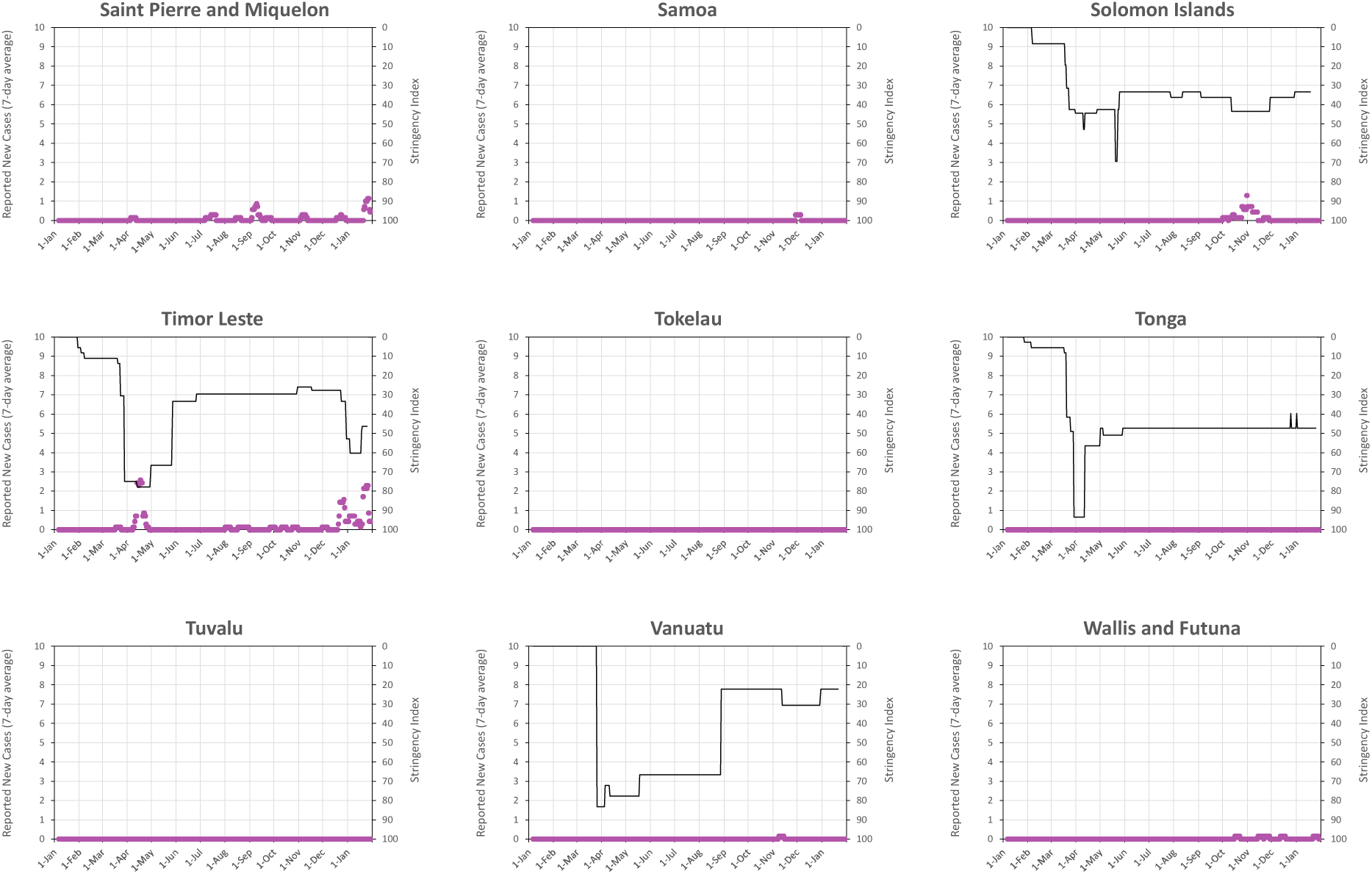

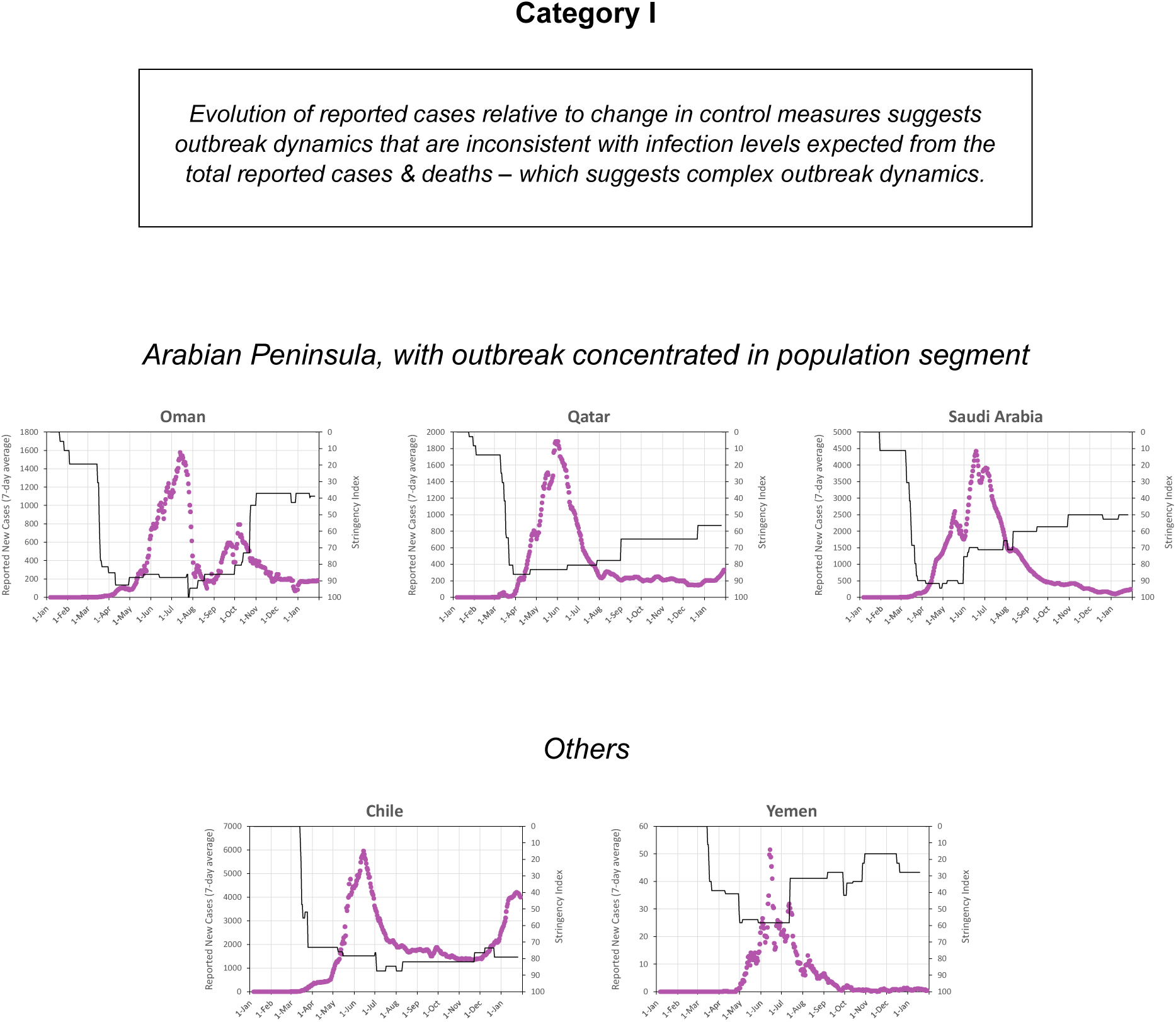

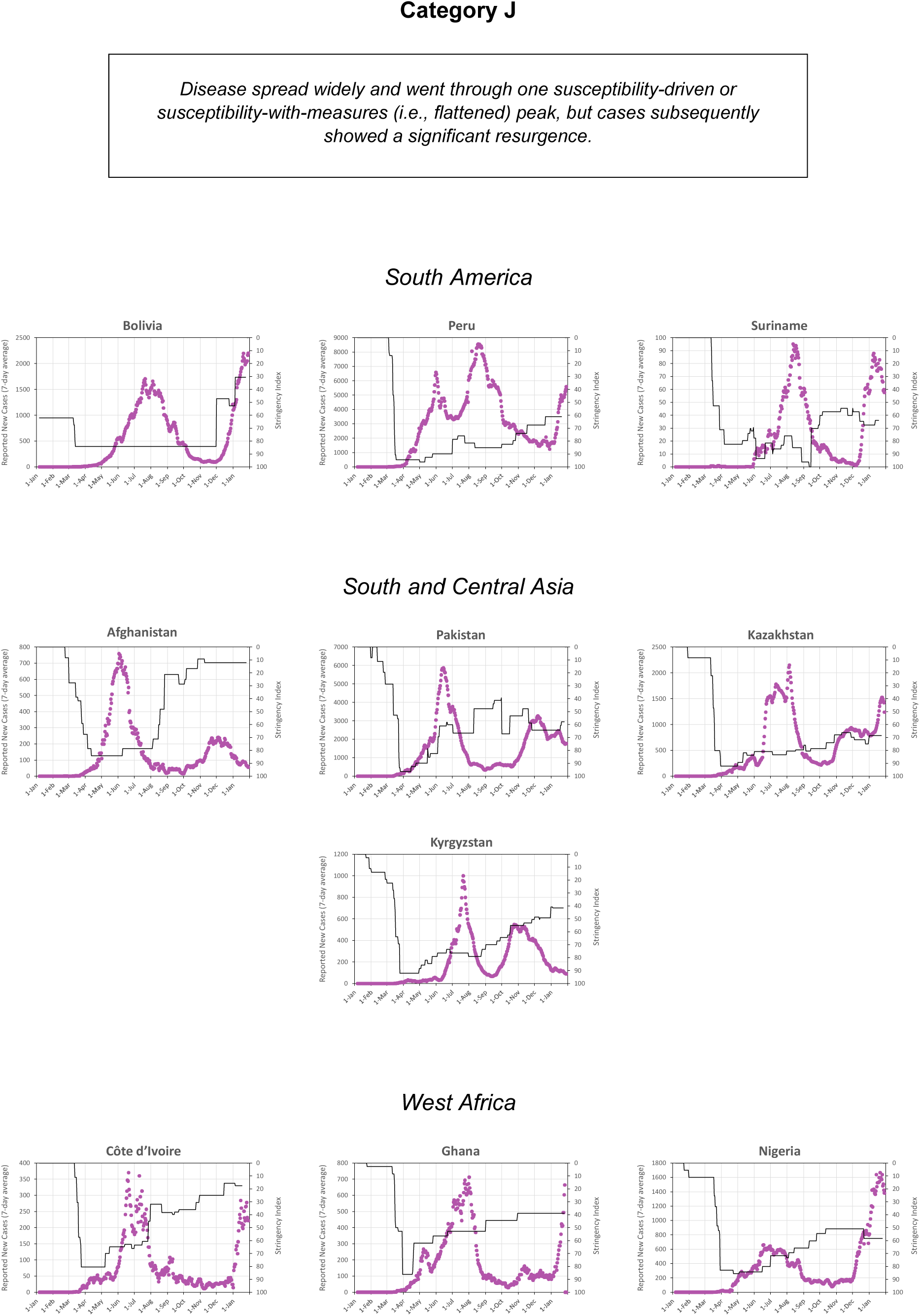

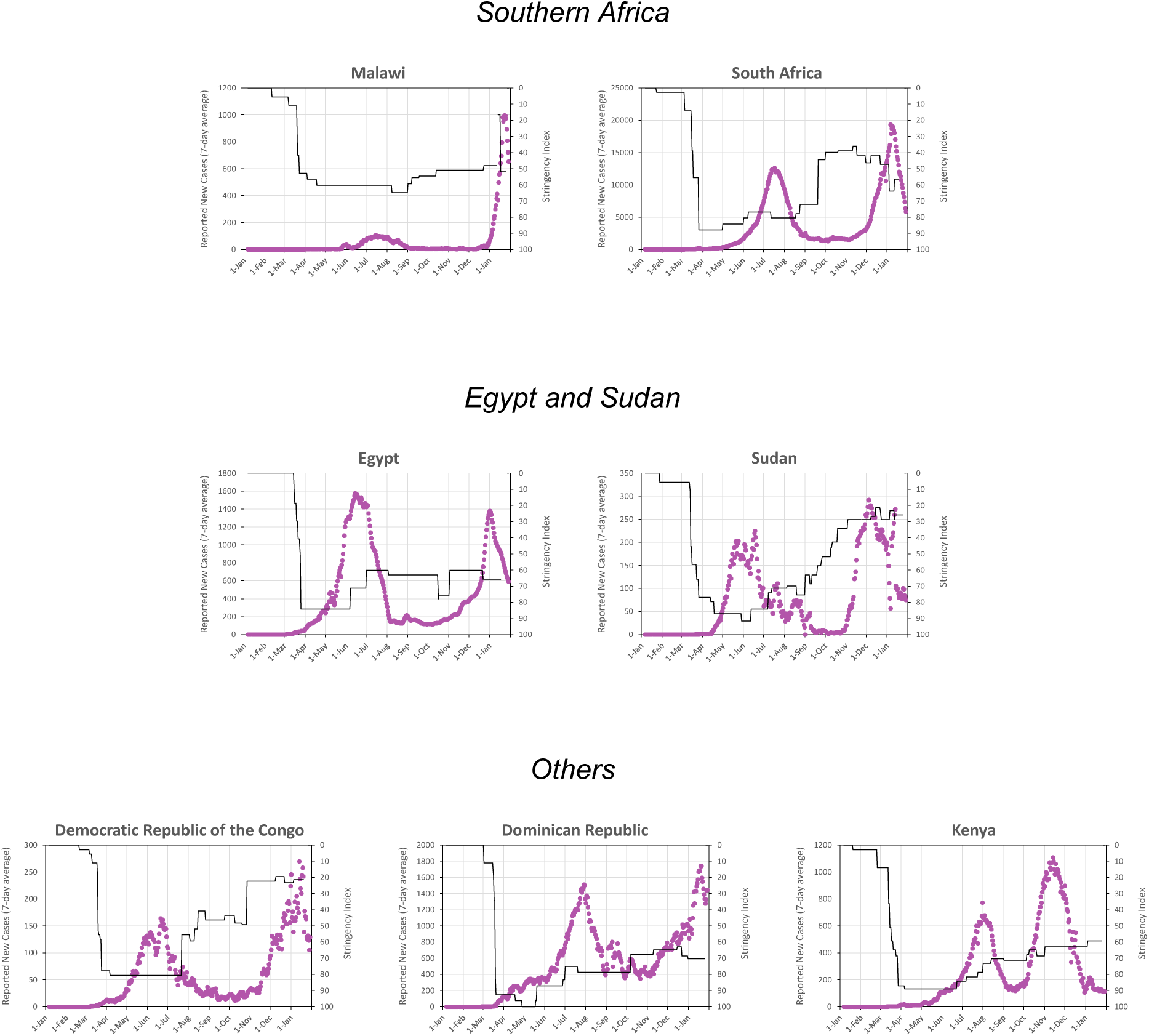

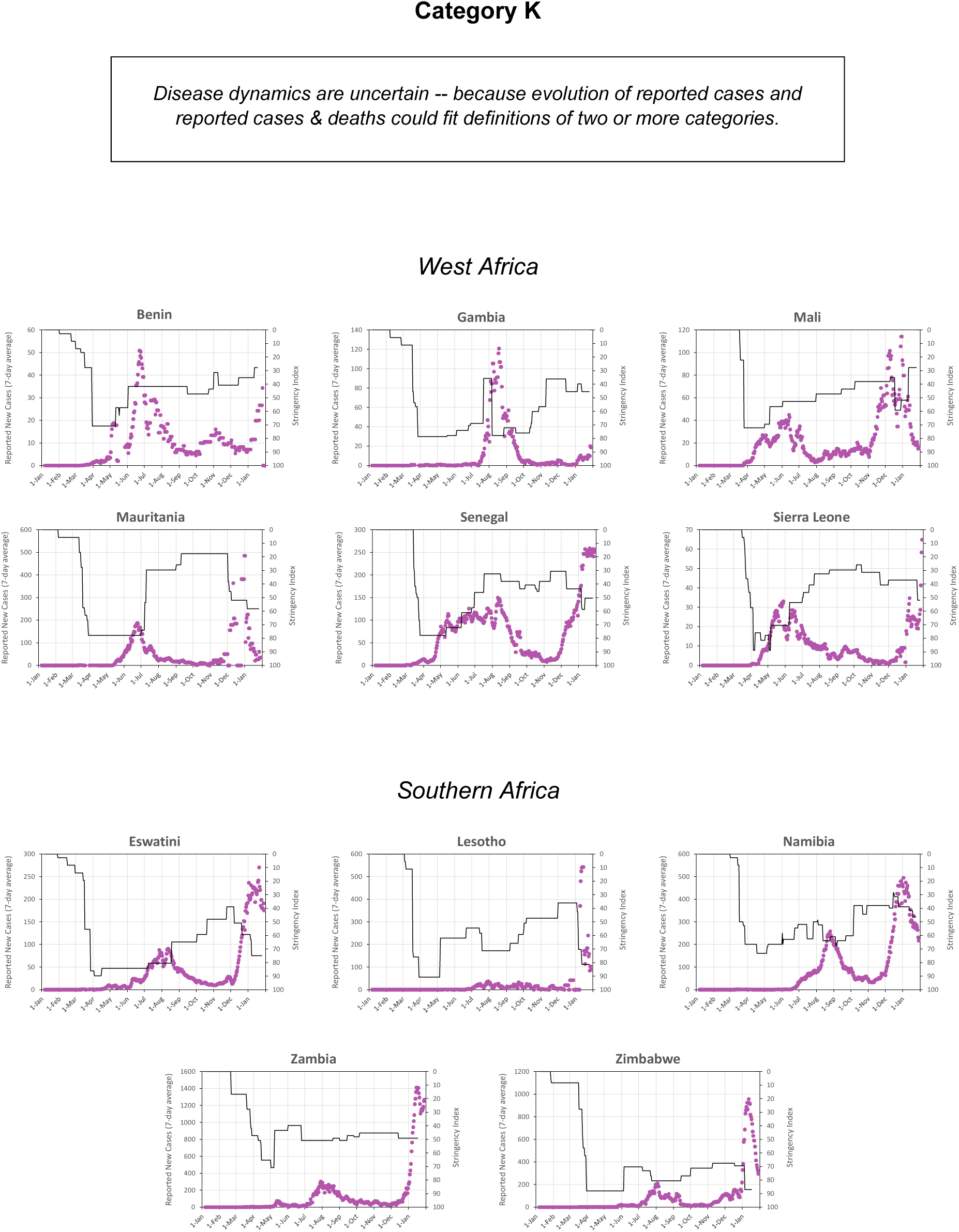

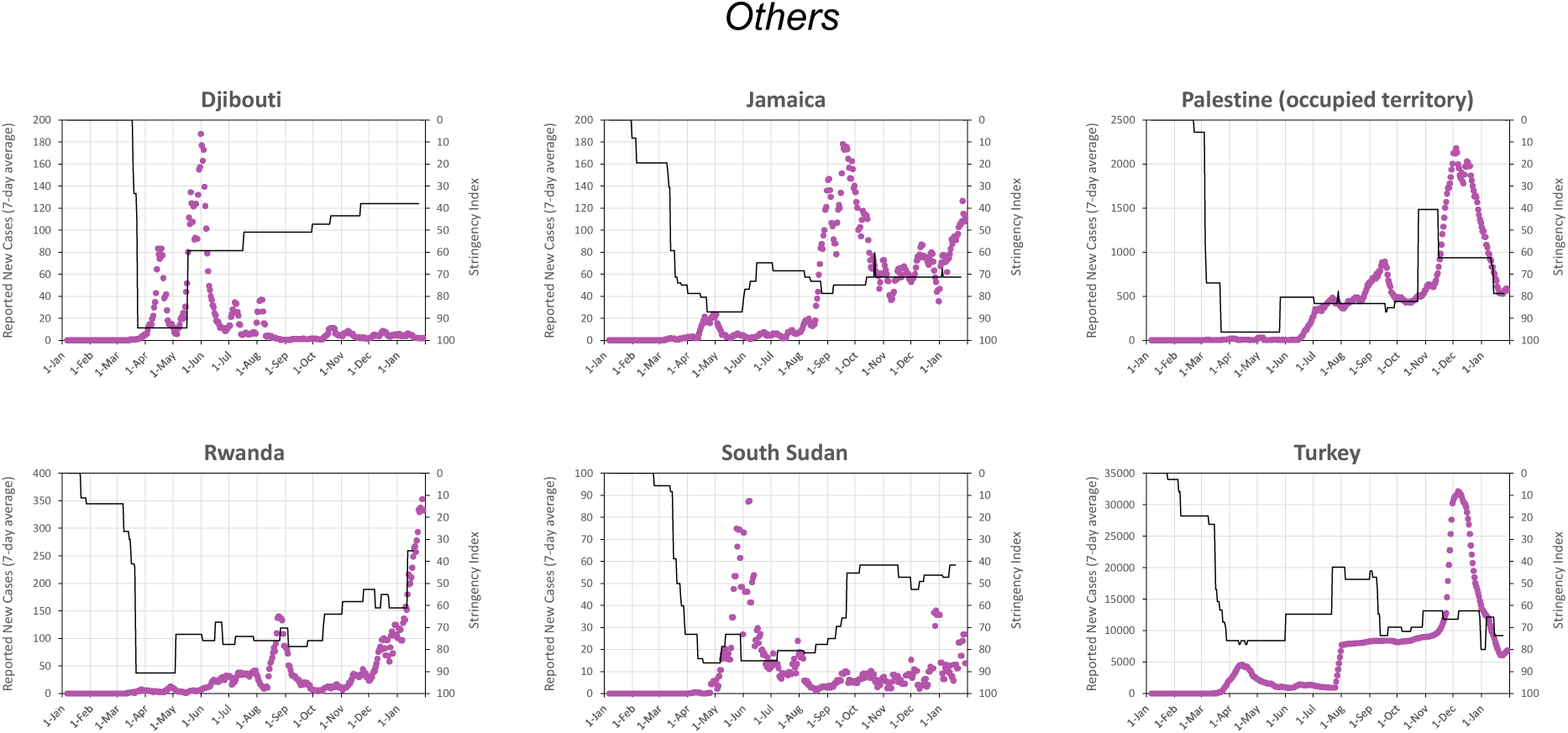

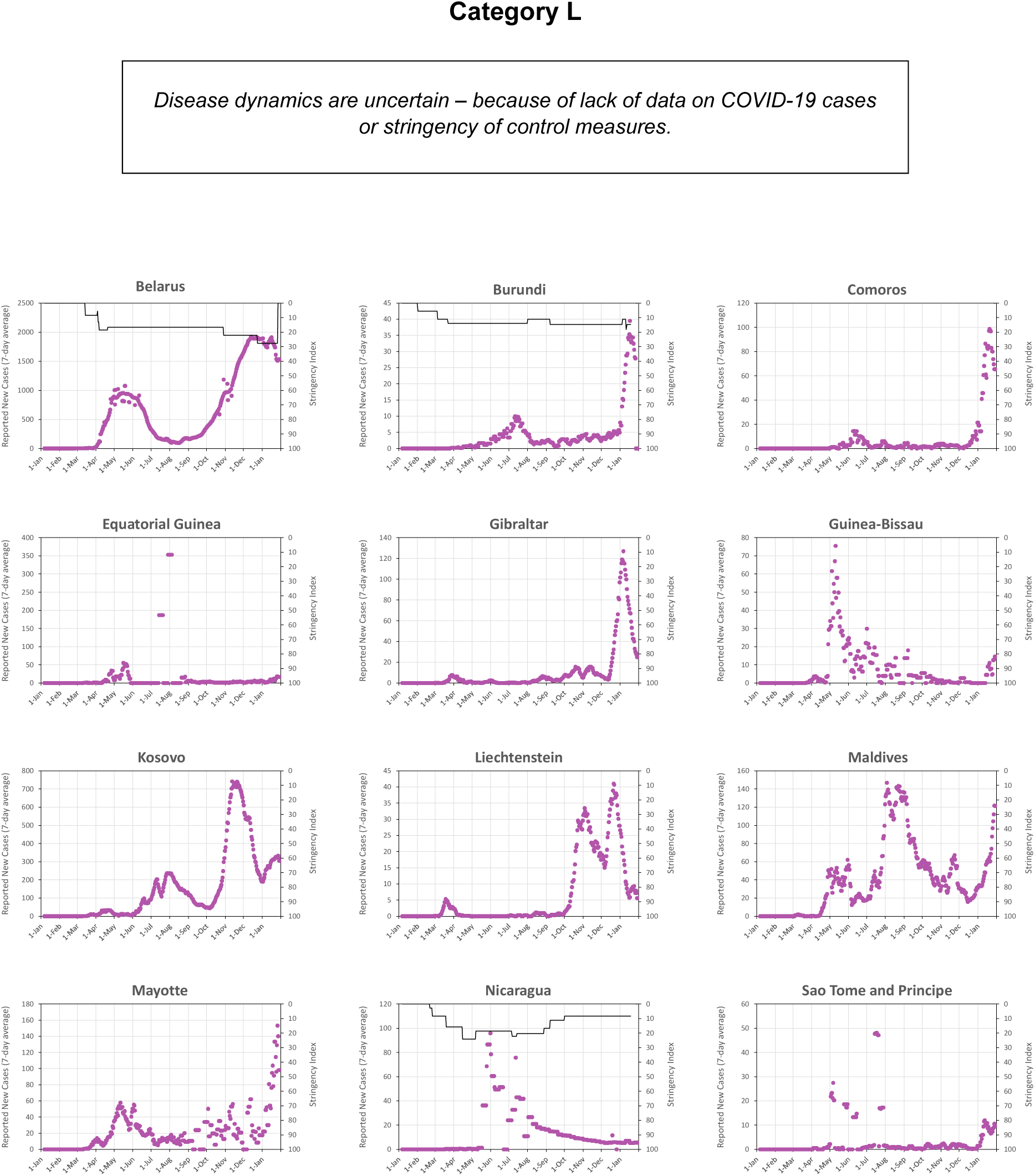

## Annex B: Total Reported Cases and Deaths and Fatality Ratios for Each Country

In this Annex, we present the data on total reported cases and deaths for each country. We also present the case fatality ratio (CFR), the ratio of reported deaths to reported cases, and the expected infection fatality ration (IFR), estimated from age-specific IFRs *[12]* and each country’s population by age. The data was downloaded on 2 February 2021.

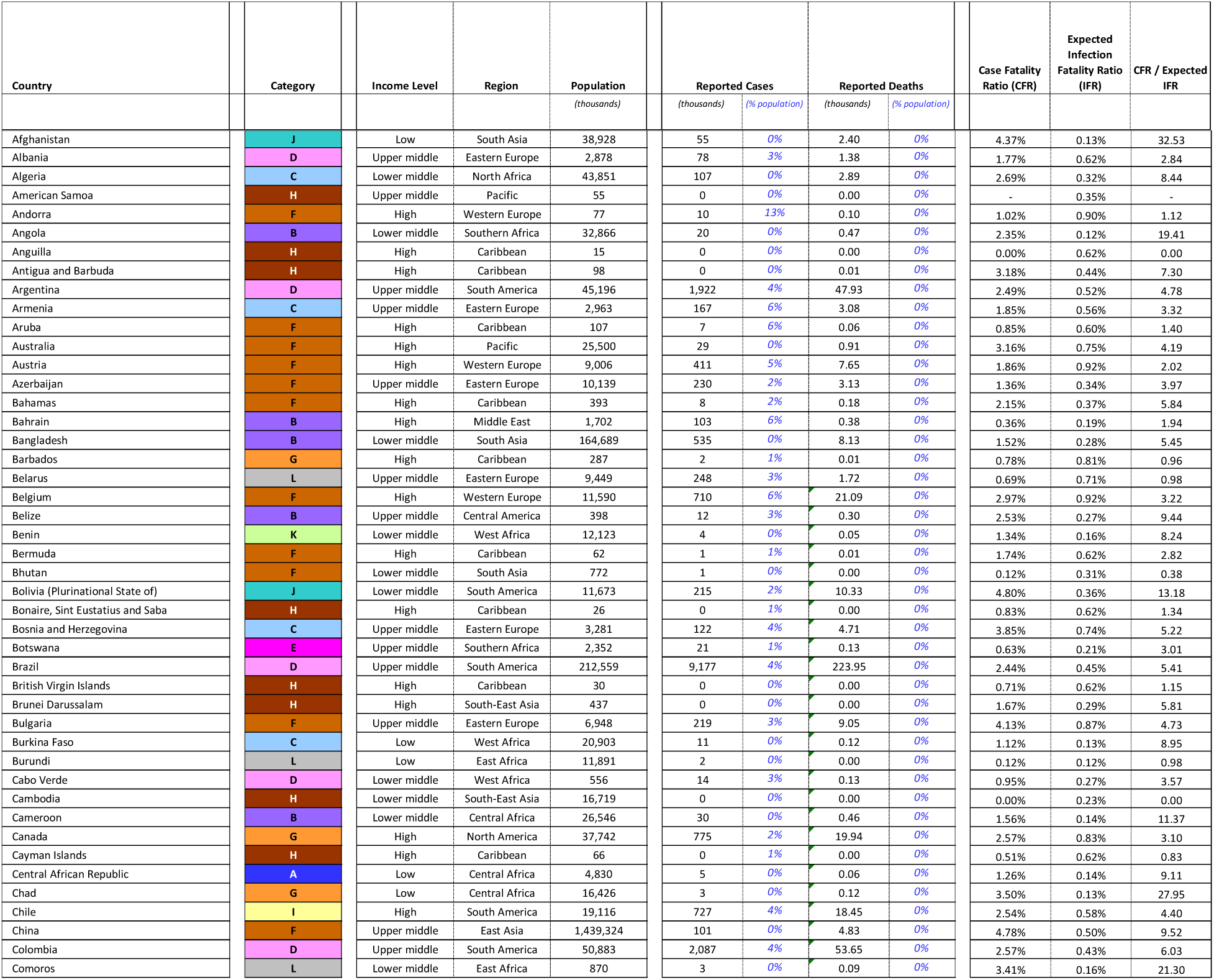
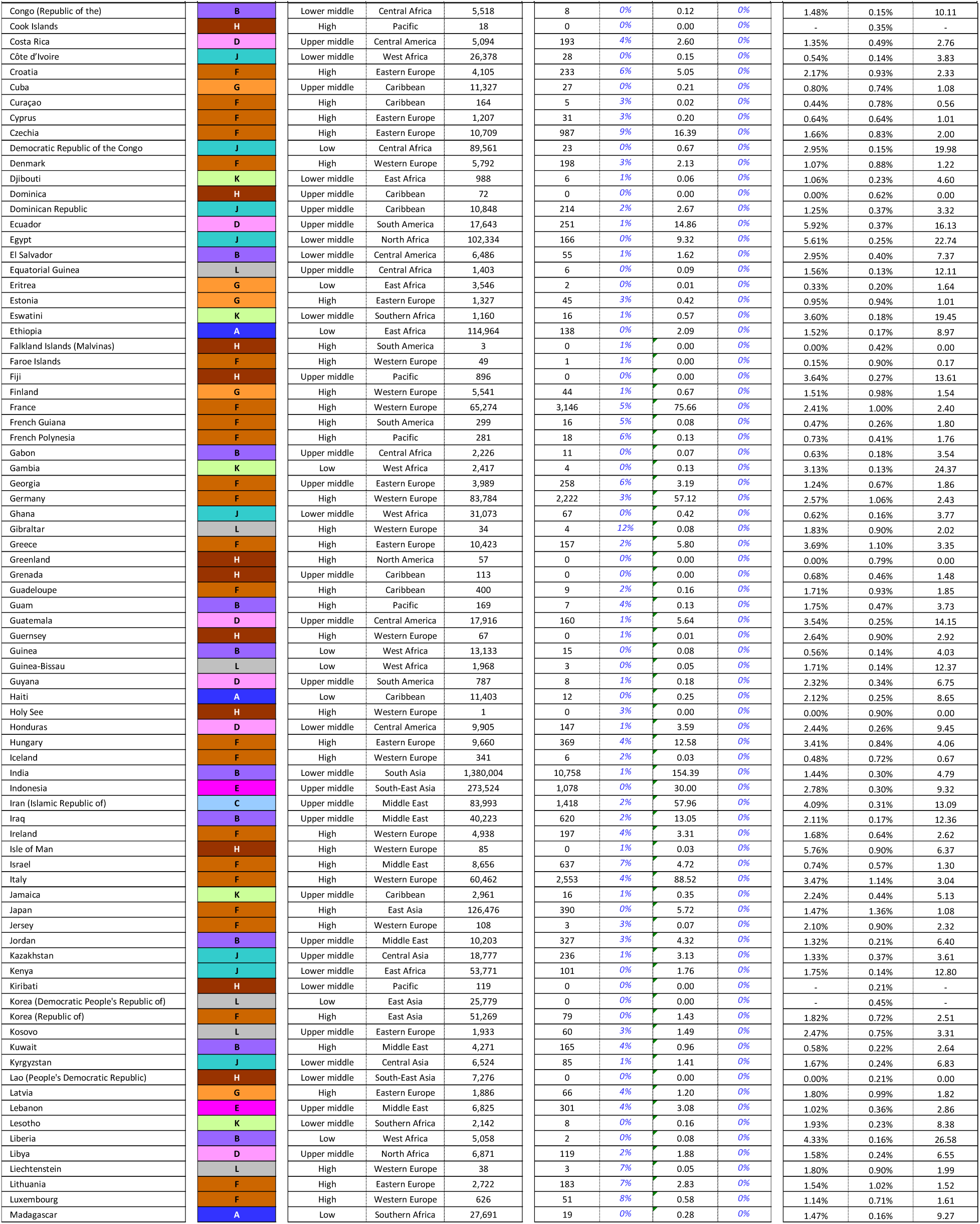
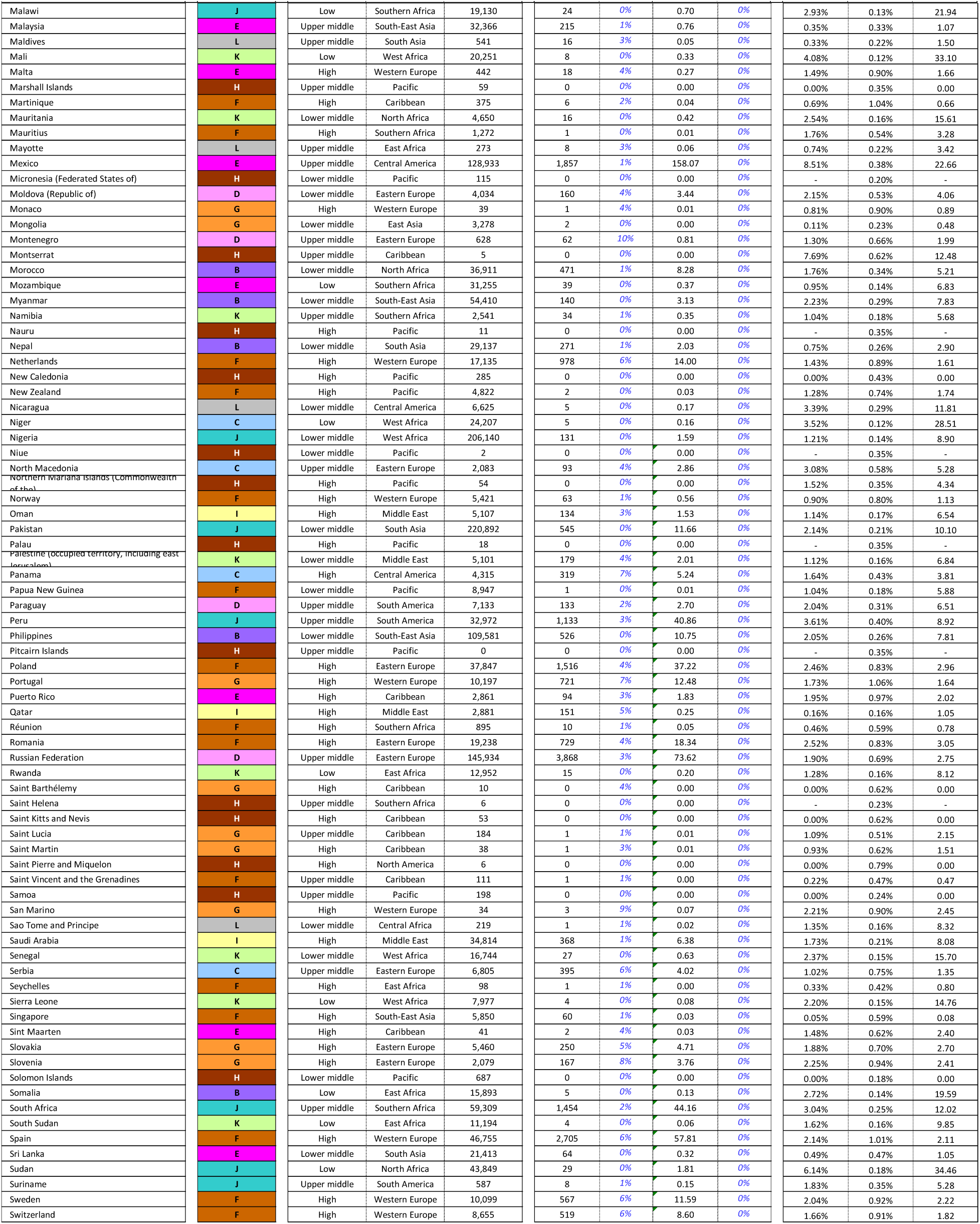
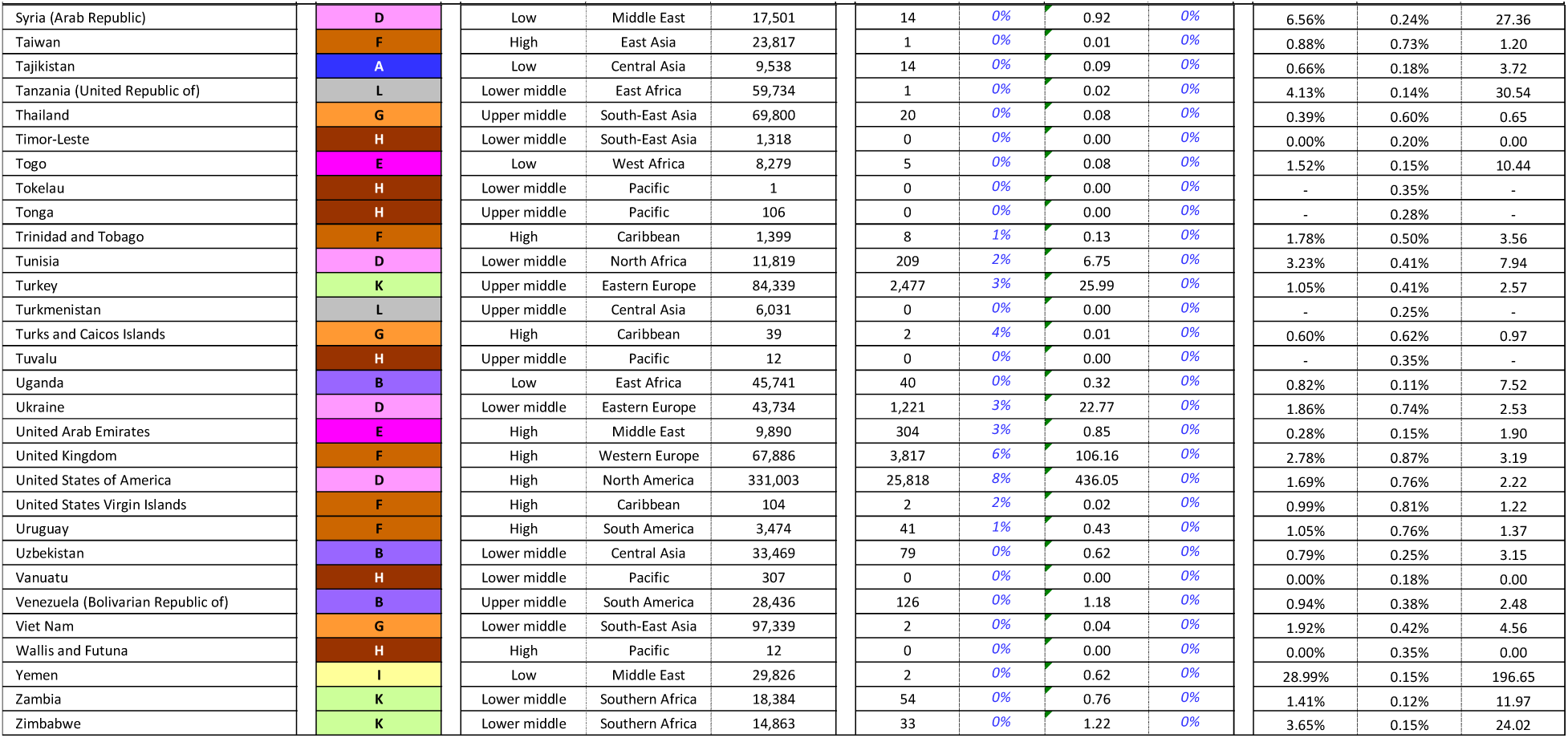

| Country | Category | Income Level | Region | Population<br>(thousands) | Reported Cases |  | Reported Deaths |  | Case Fatality<br>Ratio (CFR) | Expected<br>Infection<br>Fatality Ratio<br>(IFR) | CFR / Expected<br>IFR |
| --- | --- | --- | --- | --- | --- | --- | --- | --- | --- | --- | --- |
|  |  |  |  |  | (thousands) | (% population) | (thousands) | (% population) |  |  |  |
| Afghanistan | J | Low | South Asia | 38,928 | 55 | 0% | 2.40 | 0% | 4.37% | 0.13% | 32.53 |
| Albania | D | Upper middle | Eastern Europe | 2,878 | 78 | 3% | 1.38 | 0% | 1.77% | 0.62% | 2.84 |
| Algeria | C | Lower middle | North Africa | 43,851 | 107 | 0% | 2.89 | 0% | 2.69% | 0.32% | 8.44 |
| American Samoa | H | Upper middle | Pacific | 55 | 0 | 0% | 0.00 | 0% | - | 0.35% | - |
| Andorra | F | High | Western Europe | 77 | 10 | 13% | 0.10 | 0% | 1.02% | 0.90% | 1.12 |
| Angola | B | Lower middle | Southern Africa | 32,866 | 20 | 0% | 0.47 | 0% | 2.35% | 0.12% | 19.41 |
| Anguilla | H | High | Caribbean | 15 | 0 | 0% | 0.00 | 0% | 0.00% | 0.62% | 0.00 |
| Antigua and Barbuda | H | High | Caribbean | 98 | 0 | 0% | 0.01 | 0% | 3.18% | 0.44% | 7.30 |
| Argentina | D | Upper middle | South America | 45,196 | 1,922 | 4% | 47.93 | 0% | 2.49% | 0.52% | 4.78 |
| Armenia | C | Upper middle | Eastern Europe | 2,963 | 167 | 6% | 3.08 | 0% | 1.85% | 0.56% | 3.32 |
| Aruba | F | High | Caribbean | 107 | 7 | 6% | 0.06 | 0% | 0.85% | 0.60% | 1.40 |
| Australia | F | High | Pacific | 25,500 | 29 | 0% | 0.91 | 0% | 3.16% | 0.75% | 4.19 |
| Austria | F | High | Western Europe | 9,006 | 411 | 5% | 7.65 | 0% | 1.86% | 0.92% | 2.02 |
| Azerbaijan | F | Upper middle | Eastern Europe | 10,139 | 230 | 2% | 3.13 | 0% | 1.36% | 0.34% | 3.97 |
| Bahamas | F | High | Caribbean | 393 | 8 | 2% | 0.18 | 0% | 2.15% | 0.37% | 5.84 |
| Bahrain | B | High | Middle East | 1,702 | 103 | 6% | 0.38 | 0% | 0.36% | 0.19% | 1.94 |
| Bangladesh | B | Lower middle | South Asia | 164,689 | 535 | 0% | 8.13 | 0% | 1.52% | 0.28% | 5.45 |
| Barbados | G | High | Caribbean | 287 | 2 | 1% | 0.01 | 0% | 0.78% | 0.81% | 0.96 |
| Belarus | L | Upper middle | Eastern Europe | 9,449 | 248 | 3% | 1.72 | 0% | 0.69% | 0.71% | 0.98 |
| Belgium | F | High | Western Europe | 11,590 | 710 | 6% | 21.09 | 0% | 2.97% | 0.92% | 3.22 |
| Belize | B | Upper middle | Central America | 398 | 12 | 3% | 0.30 | 0% | 2.53% | 0.27% | 9.44 |
| Benin | K | Lower middle | West Africa | 12,123 | 4 | 0% | 0.05 | 0% | 1.34% | 0.16% | 8.24 |
| Bermuda | F | High | Caribbean | 62 | 1 | 1% | 0.01 | 0% | 1.74% | 0.62% | 2.82 |
| Bhutan | F | Lower middle | South Asia | 772 | 1 | 0% | 0.00 | 0% | 0.12% | 0.31% | 0.38 |
| Bolivia (Plurinational State of) | J | Lower middle | South America | 11,673 | 215 | 2% | 10.33 | 0% | 4.80% | 0.36% | 13.18 |
| Bonaire, Sint Eustatius and Saba | H | High | Caribbean | 26 | 0 | 1% | 0.00 | 0% | 0.83% | 0.62% | 1.34 |
| Bosnia and Herzegovina | C | Upper middle | Eastern Europe | 3,281 | 122 | 4% | 4.71 | 0% | 3.85% | 0.74% | 5.22 |
| Botswana | E | Upper middle | Southern Africa | 2,352 | 21 | 1% | 0.13 | 0% | 0.63% | 0.21% | 3.01 |
| Brazil | D | Upper middle | South America | 212,559 | 9,177 | 4% | 223.95 | 0% | 2.44% | 0.45% | 5.41 |
| British Virgin Islands | H | High | Caribbean | 30 | 0 | 0% | 0.00 | 0% | 0.71% | 0.62% | 1.15 |
| Brunei Darussalam | H | High | South-East Asia | 437 | 0 | 0% | 0.00 | 0% | 1.67% | 0.29% | 5.81 |
| Bulgaria | F | Upper middle | Eastern Europe | 6,948 | 219 | 3% | 9.05 | 0% | 4.13% | 0.87% | 4.73 |
| Burkina Faso | C | Low | West Africa | 20,903 | 11 | 0% | 0.12 | 0% | 1.12% | 0.13% | 8.95 |
| Burundi | L | Low | East Africa | 11,891 | 2 | 0% | 0.00 | 0% | 0.12% | 0.12% | 0.98 |
| Cabo Verde | D | Lower middle | West Africa | 556 | 14 | 3% | 0.13 | 0% | 0.95% | 0.27% | 3.57 |
| Cambodia | H | Lower middle | South-East Asia | 16,719 | 0 | 0% | 0.00 | 0% | 0.00% | 0.23% | 0.00 |
| Cameroon | B | Lower middle | Central Africa | 26,546 | 30 | 0% | 0.46 | 0% | 1.56% | 0.14% | 11.37 |
| Canada | G | High | North America | 37,742 | 775 | 2% | 19.94 | 0% | 2.57% | 0.83% | 3.10 |
| Cayman Islands | H | High | Caribbean | 66 | 0 | 1% | 0.00 | 0% | 0.51% | 0.62% | 0.83 |
| Central African Republic | A | Low | Central Africa | 4,830 | 5 | 0% | 0.06 | 0% | 1.26% | 0.14% | 9.11 |
| Chad | G | Low | Central Africa | 16,426 | 3 | 0% | 0.12 | 0% | 3.50% | 0.13% | 27.95 |
| Chile | I | High | South America | 19,116 | 727 | 4% | 18.45 | 0% | 2.54% | 0.58% | 4.40 |
| China | F | Upper middle | East Asia | 1,439,324 | 101 | 0% | 4.83 | 0% | 4.78% | 0.50% | 9.52 |
| Colombia | D | Upper middle | South America | 50,883 | 2,087 | 4% | 53.65 | 0% | 2.57% | 0.43% | 6.03 |
| Comoros | L | Lower middle | East Africa | 870 | 3 | 0% | 0.09 | 0% | 3.41% | 0.16% | 21.30 |
|  |  |  |  |  | (thousands) | (% population) | (thousands) | (% population) |  |  |  |
| Congo (Republic of the) | B | Lower middle | Central Africa | 5,518 | 8 | 0% | 0.12 | 0% | 1.48% | 0.15% | 10.11 |
| Cook Islands | H | High | Pacific | 18 | 0 | 0% | 0.00 | 0% | - | 0.35% | - |
| Costa Rica | D | Upper middle | Central America | 5,094 | 193 | 4% | 2.60 | 0% | 1.35% | 0.49% | 2.76 |
| Côte d'Ivoire | J | Lower middle | West Africa | 26,378 | 28 | 0% | 0.15 | 0% | 0.54% | 0.14% | 3.83 |
| Croatia | F | High | Eastern Europe | 4,105 | 233 | 6% | 5.05 | 0% | 2.17% | 0.93% | 2.33 |
| Cuba | G | Upper middle | Caribbean | 11,327 | 27 | 0% | 0.21 | 0% | 0.80% | 0.74% | 1.08 |
| Curaçao | F | High | Caribbean | 164 | 5 | 3% | 0.02 | 0% | 0.44% | 0.78% | 0.56 |
| Cyprus | F | High | Eastern Europe | 1,207 | 31 | 3% | 0.20 | 0% | 0.64% | 0.64% | 1.01 |
| Czechia | F | High | Eastern Europe | 10,709 | 987 | 9% | 16.39 | 0% | 1.66% | 0.83% | 2.00 |
| Democratic Republic of the Congo | J | Low | Central Africa | 89,561 | 23 | 0% | 0.67 | 0% | 2.95% | 0.15% | 19.98 |
| Denmark | F | High | Western Europe | 5,792 | 198 | 3% | 2.13 | 0% | 1.07% | 0.88% | 1.22 |
| Djibouti | K | Lower middle | East Africa | 988 | 6 | 1% | 0.06 | 0% | 1.06% | 0.23% | 4.60 |
| Dominica | H | Upper middle | Caribbean | 72 | 0 | 0% | 0.00 | 0% | 0.00% | 0.62% | 0.00 |
| Dominican Republic | J | Upper middle | Caribbean | 10,848 | 214 | 2% | 2.67 | 0% | 1.25% | 0.37% | 3.32 |
| Ecuador | D | Upper middle | South America | 17,643 | 251 | 1% | 14.86 | 0% | 5.92% | 0.37% | 16.13 |
| Egypt | J | Lower middle | North Africa | 102,334 | 166 | 0% | 9.32 | 0% | 5.61% | 0.25% | 22.74 |
| El Salvador | B | Lower middle | Central America | 6,486 | 55 | 1% | 1.62 | 0% | 2.95% | 0.40% | 7.37 |
| Equatorial Guinea | L | Upper middle | Central Africa | 1,403 | 6 | 0% | 0.09 | 0% | 1.56% | 0.13% | 12.11 |
| Eritrea | G | Low | East Africa | 3,546 | 2 | 0% | 0.01 | 0% | 0.33% | 0.20% | 1.64 |
| Estonia | G | High | Eastern Europe | 1,327 | 45 | 3% | 0.42 | 0% | 0.95% | 0.94% | 1.01 |
| Eswatini | K | Lower middle | Southern Africa | 1,160 | 16 | 1% | 0.57 | 0% | 3.60% | 0.18% | 19.45 |
| Ethiopia | A | Low | East Africa | 114,964 | 138 | 0% | 2.09 | 0% | 1.52% | 0.17% | 8.97 |
| Falkland Islands (Malvinas) | H | High | South America | 3 | 0 | 1% | 0.00 | 0% | 0.00% | 0.42% | 0.00 |
| Faroe Islands | F | High | Western Europe | 49 | 1 | 1% | 0.00 | 0% | 0.15% | 0.90% | 0.17 |
| Fiji | H | Upper middle | Pacific | 896 | 0 | 0% | 0.00 | 0% | 3.64% | 0.27% | 13.61 |
| Finland | G | High | Western Europe | 5,541 | 44 | 1% | 0.67 | 0% | 1.51% | 0.98% | 1.54 |
| France | F | High | Western Europe | 65,274 | 3,146 | 5% | 75.66 | 0% | 2.41% | 1.00% | 2.40 |
| French Guiana | F | High | South America | 299 | 16 | 5% | 0.08 | 0% | 0.47% | 0.26% | 1.80 |
| French Polynesia | F | High | Pacific | 281 | 18 | 6% | 0.13 | 0% | 0.73% | 0.41% | 1.76 |
| Gabon | B | Upper middle | Central Africa | 2,226 | 11 | 0% | 0.07 | 0% | 0.63% | 0.18% | 3.54 |
| Gambia | K | Low | West Africa | 2,417 | 4 | 0% | 0.13 | 0% | 3.13% | 0.13% | 24.37 |
| Georgia | F | Upper middle | Eastern Europe | 3,989 | 258 | 6% | 3.19 | 0% | 1.24% | 0.67% | 1.86 |
| Germany | F | High | Western Europe | 83,784 | 2,222 | 3% | 57.12 | 0% | 2.57% | 1.06% | 2.43 |
| Ghana | J | Lower middle | West Africa | 31,073 | 67 | 0% | 0.42 | 0% | 0.62% | 0.16% | 3.77 |
| Gibraltar | L | High | Western Europe | 34 | 4 | 12% | 0.08 | 0% | 1.83% | 0.90% | 2.02 |
| Greece | F | High | Eastern Europe | 10,423 | 157 | 2% | 5.80 | 0% | 3.69% | 1.10% | 3.35 |
| Greenland | H | High | North America | 57 | 0 | 0% | 0.00 | 0% | 0.00% | 0.79% | 0.00 |
| Grenada | H | Upper middle | Caribbean | 113 | 0 | 0% | 0.00 | 0% | 0.68% | 0.46% | 1.48 |
| Guadeloupe | F | High | Caribbean | 400 | 9 | 2% | 0.16 | 0% | 1.71% | 0.93% | 1.85 |
| Guam | B | High | Pacific | 169 | 7 | 4% | 0.13 | 0% | 1.75% | 0.47% | 3.73 |
| Guatemala | D | Upper middle | Central America | 17,916 | 160 | 1% | 5.64 | 0% | 3.54% | 0.25% | 14.15 |
| Guernsey | H | High | Western Europe | 67 | 0 | 1% | 0.01 | 0% | 2.64% | 0.90% | 2.92 |
| Guinea | B | Low | West Africa | 13,133 | 15 | 0% | 0.08 | 0% | 0.56% | 0.14% | 4.03 |
| Guinea-Bissau | L | Low | West Africa | 1,968 | 3 | 0% | 0.05 | 0% | 1.71% | 0.14% | 12.37 |
| Guyana | D | Upper middle | South America | 787 | 8 | 1% | 0.18 | 0% | 2.32% | 0.34% | 6.75 |
| Haiti | A | Low | Caribbean | 11,403 | 12 | 0% | 0.25 | 0% | 2.12% | 0.25% | 8.65 |
| Holy See | H | High | Western Europe | 1 | 0 | 3% | 0.00 | 0% | 0.00% | 0.90% | 0.00 |
| Honduras | D | Lower middle | Central America | 9,905 | 147 | 1% | 3.59 | 0% | 2.44% | 0.26% | 9.45 |
| Hungary | F | High | Eastern Europe | 9,660 | 369 | 4% | 12.58 | 0% | 3.41% | 0.84% | 4.06 |
| Iceland | F | High | Western Europe | 341 | 6 | 2% | 0.03 | 0% | 0.48% | 0.72% | 0.67 |
| India | B | Lower middle | South Asia | 1,380,004 | 10,758 | 1% | 154.39 | 0% | 1.44% | 0.30% | 4.79 |
| Indonesia | E | Upper middle | South-East Asia | 273,524 | 1,078 | 0% | 30.00 | 0% | 2.78% | 0.30% | 9.32 |
| Iran (Islamic Republic of) | C | Upper middle | Middle East | 83,993 | 1,418 | 2% | 57.96 | 0% | 4.09% | 0.31% | 13.09 |
| Iraq | B | Upper middle | Middle East | 40,223 | 620 | 2% | 13.05 | 0% | 2.11% | 0.17% | 12.36 |
| Ireland | F | High | Western Europe | 4,938 | 197 | 4% | 3.31 | 0% | 1.68% | 0.64% | 2.62 |
| Isle of Man | H | High | Western Europe | 85 | 0 | 1% | 0.03 | 0% | 5.76% | 0.90% | 6.37 |
| Israel | F | High | Middle East | 8,656 | 637 | 7% | 4.72 | 0% | 0.74% | 0.57% | 1.30 |
| Italy | F | High | Western Europe | 60,462 | 2,553 | 4% | 88.52 | 0% | 3.47% | 1.14% | 3.04 |
| Jamaica | K | Upper middle | Caribbean | 2,961 | 16 | 1% | 0.35 | 0% | 2.24% | 0.44% | 5.13 |
| Japan | F | High | East Asia | 126,476 | 390 | 0% | 5.72 | 0% | 1.47% | 1.36% | 1.08 |
| Jersey | F | High | Western Europe | 108 | 3 | 3% | 0.07 | 0% | 2.10% | 0.90% | 2.32 |
| Jordan | B | Upper middle | Middle East | 10,203 | 327 | 3% | 4.32 | 0% | 1.32% | 0.21% | 6.40 |
| Kazakhstan | J | Upper middle | Central Asia | 18,777 | 236 | 1% | 3.13 | 0% | 1.33% | 0.37% | 3.61 |
| Kenya | J | Lower middle | East Africa | 53,771 | 101 | 0% | 1.76 | 0% | 1.75% | 0.14% | 12.80 |
| Kiribati | H | Lower middle | Pacific | 119 | 0 | 0% | 0.00 | 0% | - | 0.21% | - |
| Korea (Democratic People's Republic of) | L | Low | East Asia | 25,779 | 0 | 0% | 0.00 | 0% | - | 0.45% | - |
| Korea (Republic of) | F | High | East Asia | 51,269 | 79 | 0% | 1.43 | 0% | 1.82% | 0.72% | 2.51 |
| Kosovo | L | Upper middle | Eastern Europe | 1,933 | 60 | 3% | 1.49 | 0% | 2.47% | 0.75% | 3.31 |
| Kuwait | B | High | Middle East | 4,271 | 165 | 4% | 0.96 | 0% | 0.58% | 0.22% | 2.64 |
| Kyrgyzstan | J | Lower middle | Central Asia | 6,524 | 85 | 1% | 1.41 | 0% | 1.67% | 0.24% | 6.83 |
| Lao (People's Democratic Republic) | H | Lower middle | South-East Asia | 7,276 | 0 | 0% | 0.00 | 0% | 0.00% | 0.21% | 0.00 |
| Latvia | G | High | Eastern Europe | 1,886 | 66 | 4% | 1.20 | 0% | 1.80% | 0.99% | 1.82 |
| Lebanon | E | Upper middle | Middle East | 6,825 | 301 | 4% | 3.08 | 0% | 1.02% | 0.36% | 2.86 |
| Lesotho | K | Lower middle | Southern Africa | 2,142 | 8 | 0% | 0.16 | 0% | 1.93% | 0.23% | 8.38 |
| Liberia | B | Low | West Africa | 5,058 | 2 | 0% | 0.08 | 0% | 4.33% | 0.16% | 26.58 |
| Libya | D | Upper middle | North Africa | 6,871 | 119 | 2% | 1.88 | 0% | 1.58% | 0.24% | 6.55 |
| Liechtenstein | L | High | Western Europe | 38 | 3 | 7% | 0.05 | 0% | 1.80% | 0.90% | 1.99 |
| Lithuania | F | High | Eastern Europe | 2,722 | 183 | 7% | 2.83 | 0% | 1.54% | 1.02% | 1.52 |
| Luxembourg | F | High | Western Europe | 626 | 51 | 8% | 0.58 | 0% | 1.14% | 0.71% | 1.61 |
| Madagascar | A | Low | Southern Africa | 27,691 | 19 | 0% | 0.28 | 0% | 1.47% | 0.16% | 9.27 |

| Country | Category | Income Level | Region | Population<br>(thousands) | Reported Cases<br>(thousands) (% population) |  | Reported Deaths<br>(thousands) (% population) |  | Case Fatality<br>Ratio (CFR) | Expected<br>Infection<br>Fatality Ratio (IFR) | CFR / Expected<br>IFR |
| --- | --- | --- | --- | --- | --- | --- | --- | --- | --- | --- | --- |
| Malawi | J | Low | Southern Africa | 19,130 | 24 | 0% | 0.70 | 0% | 2.93% | 0.13% | 21.94 |
| Malaysia | E | Upper middle | South-East Asia | 32,366 | 215 | 1% | 0.76 | 0% | 0.35% | 0.33% | 1.07 |
| Maldives | L | Upper middle | South Asia | 541 | 16 | 3% | 0.05 | 0% | 0.33% | 0.22% | 1.50 |
| Mali | K | Low | West Africa | 20,251 | 8 | 0% | 0.33 | 0% | 4.08% | 0.12% | 33.10 |
| Malta | E | High | Western Europe | 442 | 18 | 4% | 0.27 | 0% | 1.49% | 0.90% | 1.66 |
| Marshall Islands | H | Upper middle | Pacific | 59 | 0 | 0% | 0.00 | 0% | 0.00% | 0.35% | 0.00 |
| Martinique | F | High | Caribbean | 375 | 6 | 2% | 0.04 | 0% | 0.69% | 1.04% | 0.66 |
| Mauritania | K | Lower middle | North Africa | 4,650 | 16 | 0% | 0.42 | 0% | 2.54% | 0.16% | 15.61 |
| Mauritius | F | High | Southern Africa | 1,272 | 1 | 0% | 0.01 | 0% | 1.76% | 0.54% | 3.28 |
| Mayotte | L | Upper middle | East Africa | 273 | 8 | 3% | 0.06 | 0% | 0.74% | 0.22% | 3.42 |
| Mexico | E | Upper middle | Central America | 128,933 | 1,857 | 1% | 158.07 | 0% | 8.51% | 0.38% | 22.66 |
| Micronesia (Federated States of) | H | Lower middle | Pacific | 115 | 0 | 0% | 0.00 | 0% | - | 0.20% | - |
| Moldova (Republic of) | D | Lower middle | Eastern Europe | 4,034 | 160 | 4% | 3.44 | 0% | 2.15% | 0.53% | 4.06 |
| Monaco | G | High | Western Europe | 39 | 1 | 4% | 0.01 | 0% | 0.81% | 0.90% | 0.89 |
| Mongolia | G | Lower middle | East Asia | 3,278 | 2 | 0% | 0.00 | 0% | 0.11% | 0.23% | 0.48 |
| Montenegro | D | Upper middle | Eastern Europe | 628 | 62 | 10% | 0.81 | 0% | 1.30% | 0.66% | 1.99 |
| Montserrat | H | Upper middle | Caribbean | 5 | 0 | 0% | 0.00 | 0% | 7.69% | 0.62% | 12.48 |
| Morocco | B | Lower middle | North Africa | 36,911 | 471 | 1% | 8.28 | 0% | 1.76% | 0.34% | 5.21 |
| Mozambique | E | Low | Southern Africa | 31,255 | 39 | 0% | 0.37 | 0% | 0.95% | 0.14% | 6.83 |
| Myanmar | B | Lower middle | South-East Asia | 54,410 | 140 | 0% | 3.13 | 0% | 2.23% | 0.29% | 7.83 |
| Namibia | K | Upper middle | Southern Africa | 2,541 | 34 | 1% | 0.35 | 0% | 1.04% | 0.18% | 5.68 |
| Nauru | H | High | Pacific | 11 | 0 | 0% | 0.00 | 0% | - | 0.35% | - |
| Nepal | B | Lower middle | South Asia | 29,137 | 271 | 1% | 2.03 | 0% | 0.75% | 0.26% | 2.90 |
| Netherlands | F | High | Western Europe | 17,135 | 978 | 6% | 14.00 | 0% | 1.43% | 0.89% | 1.61 |
| New Caledonia | H | High | Pacific | 285 | 0 | 0% | 0.00 | 0% | 0.00% | 0.43% | 0.00 |
| New Zealand | F | High | Pacific | 4,822 | 2 | 0% | 0.03 | 0% | 1.28% | 0.74% | 1.74 |
| Nicaragua | L | Lower middle | Central America | 6,625 | 5 | 0% | 0.17 | 0% | 3.39% | 0.29% | 11.81 |
| Niger | C | Low | West Africa | 24,207 | 5 | 0% | 0.16 | 0% | 3.52% | 0.12% | 28.51 |
| Nigeria | J | Lower middle | West Africa | 206,140 | 131 | 0% | 1.59 | 0% | 1.21% | 0.14% | 8.90 |
| Niue | H | Lower middle | Pacific | 2 | 0 | 0% | 0.00 | 0% | - | 0.35% | - |
| North Macedonia | C | Upper middle | Eastern Europe | 2,083 | 93 | 4% | 2.86 | 0% | 3.08% | 0.58% | 5.28 |
| Northern Mariana Islands (commonwealth of the U.S.) | H | High | Pacific | 54 | 0 | 0% | 0.00 | 0% | 1.52% | 0.35% | 4.34 |
| Norway | F | High | Western Europe | 5,421 | 63 | 1% | 0.56 | 0% | 0.90% | 0.80% | 1.13 |
| Oman | I | High | Middle East | 5,107 | 134 | 3% | 1.53 | 0% | 1.14% | 0.17% | 6.54 |
| Pakistan | J | Lower middle | South Asia | 220,892 | 545 | 0% | 11.66 | 0% | 2.14% | 0.21% | 10.10 |
| Palau | H | High | Pacific | 18 | 0 | 0% | 0.00 | 0% | - | 0.35% | - |
| Palestine (occupied territory, including east Jerusalem) | K | Lower middle | Middle East | 5,101 | 179 | 4% | 2.01 | 0% | 1.12% | 0.16% | 6.84 |
| Panama | C | High | Central America | 4,315 | 319 | 7% | 5.24 | 0% | 1.64% | 0.43% | 3.81 |
| Papua New Guinea | F | Lower middle | Pacific | 8,947 | 1 | 0% | 0.01 | 0% | 1.04% | 0.18% | 5.88 |
| Paraguay | D | Upper middle | South America | 7,133 | 133 | 2% | 2.70 | 0% | 2.04% | 0.31% | 6.51 |
| Peru | J | Upper middle | South America | 32,972 | 1,133 | 3% | 40.86 | 0% | 3.61% | 0.40% | 8.92 |
| Philippines | B | Lower middle | South-East Asia | 109,581 | 526 | 0% | 10.75 | 0% | 2.05% | 0.26% | 7.81 |
| Pitcairn Islands | H | Upper middle | Pacific | 0 | 0 | 0% | 0.00 | 0% | - | 0.35% | - |
| Poland | F | High | Eastern Europe | 37,847 | 1,516 | 4% | 37.22 | 0% | 2.46% | 0.83% | 2.96 |
| Portugal | G | High | Western Europe | 10,197 | 721 | 7% | 12.48 | 0% | 1.73% | 1.06% | 1.64 |
| Puerto Rico | E | High | Caribbean | 2,861 | 94 | 3% | 1.83 | 0% | 1.95% | 0.97% | 2.02 |
| Qatar | I | High | Middle East | 2,881 | 151 | 5% | 0.25 | 0% | 0.16% | 0.16% | 1.05 |
| Réunion | F | High | Southern Africa | 895 | 10 | 1% | 0.05 | 0% | 0.46% | 0.59% | 0.78 |
| Romania | F | High | Eastern Europe | 19,238 | 729 | 4% | 18.34 | 0% | 2.52% | 0.83% | 3.05 |
| Russian Federation | D | Upper middle | Eastern Europe | 145,934 | 3,868 | 3% | 73.62 | 0% | 1.90% | 0.69% | 2.75 |
| Rwanda | K | Low | East Africa | 12,952 | 15 | 0% | 0.20 | 0% | 1.28% | 0.16% | 8.12 |
| Saint Barthélemy | G | High | Caribbean | 10 | 0 | 4% | 0.00 | 0% | 0.00% | 0.62% | 0.00 |
| Saint Helena | H | Upper middle | Southern Africa | 6 | 0 | 0% | 0.00 | 0% | - | 0.23% | - |
| Saint Kitts and Nevis | H | High | Caribbean | 53 | 0 | 0% | 0.00 | 0% | 0.00% | 0.62% | 0.00 |
| Saint Lucia | G | Upper middle | Caribbean | 184 | 1 | 1% | 0.01 | 0% | 1.09% | 0.51% | 2.15 |
| Saint Martin | G | High | Caribbean | 38 | 1 | 3% | 0.01 | 0% | 0.93% | 0.62% | 1.51 |
| Saint Pierre and Miquelon | H | High | North America | 6 | 0 | 0% | 0.00 | 0% | 0.00% | 0.79% | 0.00 |
| Saint Vincent and the Grenadines | F | Upper middle | Caribbean | 111 | 1 | 1% | 0.00 | 0% | 0.22% | 0.47% | 0.47 |
| Samoa | H | Upper middle | Pacific | 198 | 0 | 0% | 0.00 | 0% | 0.00% | 0.24% | 0.00 |
| San Marino | G | High | Western Europe | 34 | 3 | 9% | 0.07 | 0% | 2.21% | 0.90% | 2.45 |
| Sao Tome and Principe | L | Lower middle | Central Africa | 219 | 1 | 1% | 0.02 | 0% | 1.35% | 0.16% | 8.32 |
| Saudi Arabia | I | High | Middle East | 34,814 | 368 | 1% | 6.38 | 0% | 1.73% | 0.21% | 8.08 |
| Senegal | K | Lower middle | West Africa | 16,744 | 27 | 0% | 0.63 | 0% | 2.37% | 0.15% | 15.70 |
| Serbia | C | Upper middle | Eastern Europe | 6,805 | 395 | 6% | 4.02 | 0% | 1.02% | 0.75% | 1.35 |
| Seychelles | F | High | East Africa | 98 | 1 | 1% | 0.00 | 0% | 0.33% | 0.42% | 0.80 |
| Sierra Leone | K | Low | West Africa | 7,977 | 4 | 0% | 0.08 | 0% | 2.20% | 0.15% | 14.76 |
| Singapore | F | High | South-East Asia | 5,850 | 60 | 1% | 0.03 | 0% | 0.05% | 0.59% | 0.08 |
| Sint Maarten | E | High | Caribbean | 41 | 2 | 4% | 0.03 | 0% | 1.48% | 0.62% | 2.40 |
| Slovakia | G | High | Eastern Europe | 5,460 | 250 | 5% | 4.71 | 0% | 1.88% | 0.70% | 2.70 |
| Slovenia | G | High | Eastern Europe | 2,079 | 167 | 8% | 3.76 | 0% | 2.25% | 0.94% | 2.41 |
| Solomon Islands | H | Lower middle | Pacific | 687 | 0 | 0% | 0.00 | 0% | 0.00% | 0.18% | 0.00 |
| Somalia | B | Low | East Africa | 15,893 | 5 | 0% | 0.13 | 0% | 2.72% | 0.14% | 19.59 |
| South Africa | J | Upper middle | Southern Africa | 59,309 | 1,454 | 2% | 44.16 | 0% | 3.04% | 0.25% | 12.02 |
| South Sudan | K | Low | East Africa | 11,194 | 4 | 0% | 0.06 | 0% | 1.62% | 0.16% | 9.85 |
| Spain | F | High | Western Europe | 46,755 | 2,705 | 6% | 57.81 | 0% | 2.14% | 1.01% | 2.11 |
| Sri Lanka | E | Lower middle | South Asia | 21,413 | 64 | 0% | 0.32 | 0% | 0.49% | 0.47% | 1.05 |
| Sudan | J | Low | North Africa | 43,849 | 29 | 0% | 1.81 | 0% | 6.14% | 0.18% | 34.46 |
| Suriname | J | Upper middle | South America | 587 | 8 | 1% | 0.15 | 0% | 1.83% | 0.35% | 5.28 |
| Sweden | F | High | Western Europe | 10,099 | 567 | 6% | 11.59 | 0% | 2.04% | 0.92% | 2.22 |
| Switzerland | F | High | Western Europe | 8,655 | 519 | 6% | 8.60 | 0% | 1.66% | 0.91% | 1.82 |
|  |  |  |  |  | (thousands) | (% population) | (thousands) | (% population) |  |  |  |
| Syria (Arab Republic) | D | Low | Middle East | 17,501 | 14 | 0% | 0.92 | 0% | 6.56% | 0.24% | 27.36 |
| Taiwan | F | High | East Asia | 23,817 | 1 | 0% | 0.01 | 0% | 0.88% | 0.73% | 1.20 |
| Tajikistan | A | Low | Central Asia | 9,538 | 14 | 0% | 0.09 | 0% | 0.66% | 0.18% | 3.72 |
| Tanzania (United Republic of) | L | Lower middle | East Africa | 59,734 | 1 | 0% | 0.02 | 0% | 4.13% | 0.14% | 30.54 |
| Thailand | G | Upper middle | South-East Asia | 69,800 | 20 | 0% | 0.08 | 0% | 0.39% | 0.60% | 0.65 |
| Timor-Leste | H | Lower middle | South-East Asia | 1,318 | 0 | 0% | 0.00 | 0% | 0.00% | 0.20% | 0.00 |
| Togo | E | Low | West Africa | 8,279 | 5 | 0% | 0.08 | 0% | 1.52% | 0.15% | 10.44 |
| Tokelau | H | Lower middle | Pacific | 1 | 0 | 0% | 0.00 | 0% | - | 0.35% | - |
| Tonga | H | Upper middle | Pacific | 106 | 0 | 0% | 0.00 | 0% | - | 0.28% | - |
| Trinidad and Tobago | F | High | Caribbean | 1,399 | 8 | 1% | 0.13 | 0% | 1.78% | 0.50% | 3.56 |
| Tunisia | D | Lower middle | North Africa | 11,819 | 209 | 2% | 6.75 | 0% | 3.23% | 0.41% | 7.94 |
| Turkey | K | Upper middle | Eastern Europe | 84,339 | 2,477 | 3% | 25.99 | 0% | 1.05% | 0.41% | 2.57 |
| Turkmenistan | L | Upper middle | Central Asia | 6,031 | 0 | 0% | 0.00 | 0% | - | 0.25% | - |
| Turks and Caicos Islands | G | High | Caribbean | 39 | 2 | 4% | 0.01 | 0% | 0.60% | 0.62% | 0.97 |
| Tuvalu | H | Upper middle | Pacific | 12 | 0 | 0% | 0.00 | 0% | - | 0.35% | - |
| Uganda | B | Low | East Africa | 45,741 | 40 | 0% | 0.32 | 0% | 0.82% | 0.11% | 7.52 |
| Ukraine | D | Lower middle | Eastern Europe | 43,734 | 1,221 | 3% | 22.77 | 0% | 1.86% | 0.74% | 2.53 |
| United Arab Emirates | E | High | Middle East | 9,890 | 304 | 3% | 0.85 | 0% | 0.28% | 0.15% | 1.90 |
| United Kingdom | F | High | Western Europe | 67,886 | 3,817 | 6% | 106.16 | 0% | 2.78% | 0.87% | 3.19 |
| United States of America | D | High | North America | 331,003 | 25,818 | 8% | 436.05 | 0% | 1.69% | 0.76% | 2.22 |
| United States Virgin Islands | F | High | Caribbean | 104 | 2 | 2% | 0.02 | 0% | 0.99% | 0.81% | 1.22 |
| Uruguay | F | High | South America | 3,474 | 41 | 1% | 0.43 | 0% | 1.05% | 0.76% | 1.37 |
| Uzbekistan | B | Lower middle | Central Asia | 33,469 | 79 | 0% | 0.62 | 0% | 0.79% | 0.25% | 3.15 |
| Vanuatu | H | Lower middle | Pacific | 307 | 0 | 0% | 0.00 | 0% | 0.00% | 0.18% | 0.00 |
| Venezuela (Bolivarian Republic of) | B | Upper middle | South America | 28,436 | 126 | 0% | 1.18 | 0% | 0.94% | 0.38% | 2.48 |
| Viet Nam | G | Lower middle | South-East Asia | 97,339 | 2 | 0% | 0.04 | 0% | 1.92% | 0.42% | 4.56 |
| Wallis and Futuna | H | High | Pacific | 12 | 0 | 0% | 0.00 | 0% | 0.00% | 0.35% | 0.00 |
| Yemen | I | Low | Middle East | 29,826 | 2 | 0% | 0.62 | 0% | 28.99% | 0.15% | 196.65 |
| Zambia | K | Lower middle | Southern Africa | 18,384 | 54 | 0% | 0.76 | 0% | 1.41% | 0.12% | 11.97 |
| Zimbabwe | K | Lower middle | Southern Africa | 14,863 | 33 | 0% | 1.22 | 0% | 3.65% | 0.15% | 24.02 |

